# Impact of mid-life cardiovascular health on cognitive change in a bi-ethnic cohort

**DOI:** 10.1101/2025.10.23.25338553

**Authors:** Janette Vazquez, Rebecca Bernal, Luis A. Aguirre, Jesus D. Melgarejo, Gladys Maestre, Sudha Seshadri, Helen P. Hazuda, Chen-Pin Wang, Claudia L. Satizabal

## Abstract

**INTRODUCTION:** Vascular risk factors contribute to cognitive decline and dementia; however, research across populations remains scarce.

**METHODS:** We included 402 participants (mean age 57.9±3.8, 58.5% female, 52% Mexican American). Midlife cardiovascular health was assessed using the AHA Life’s Simple 7 (LS7). General cognition was measured four times over a period of 9.5 years. We used generalized estimating equations to assess the association of LS7 scores and its factors with cognitive change, overall and by ethnicity.

**RESULTS:** Meeting intermediate or ideal physical activity goals mitigated cognitive decline, particularly in Mexican Americans, and maintaining blood glucose below 125 mg/dL was associated with slower cognitive decline among non-Hispanic Whites. Additionally, Mexican Americans with a BMI <30 kg/m^2^ showed an accelerated cognitive decline. No associations were observed between total LS7 scores and cognitive decline.

**DISCUSSION:** Physical activity, blood glucose, and BMI have distinct contributions to cognitive change across different ethnicities.

## 1. Background

An increase in global life expectancy combined with a higher proportion of older adults is leading to a higher prevalence of age-related conditions such as dementia, making prevention a global health priority ^1^. Currently, 6.7 million individuals in the United States are affected by Alzheimer’s dementia (AD), and despite declining trends in dementia incidence ^2,3^, projections estimate up to 13.8 million AD cases by 2060 ^1^. Evidence has established the significant contribution of modifiable risk factors to cognitive decline and dementia, including AD. Recent research suggests that 40% of AD cases may be attributable to modifiable risk factors, notably physical inactivity, smoking, hypertension, and obesity ^4^. However, given the tendency of risk factors to cluster and interact, it is imperative to examine their collective impact on dementia ^5^.

Considering the principal role of cardiovascular health in brain aging, the American Heart Association’s Life’s Simple 7 (LS7) has been recommended to promote optimal brain health ^6^. The American Heart Association has established national goals to improve cardiovascular health by 20%, which includes three behavioral factors (healthy diet, non-smoking, and physical activity) and four health factors (blood pressure, body mass index, total cholesterol, and fasting blood glucose). These cardiovascular health indices represent an improvement over the assessment of single cardiovascular risk factors, as they often coexist and affect the brain through common pathways ^7^.

Previous studies have examined the associations between adherence to LS7 and cognitive outcomes, with findings suggesting that a better cardiovascular health score is associated with better cognitive function^5,6,8-25^, a lower risk of dementia, and improved brain health^6,9,13-17^. Individual cardiovascular risk factors also play an essential role in the development of cognitive impairment and dementia. For example, diabetes, obesity, hypertension, high cholesterol, smoking, and low physical activity have all been associated with a faster rate of cognitive decline and an increased dementia risk ^5,13,14,19,21,22^. Conversely, elements that mitigate cardiovascular risk, such as regular physical activity and a heart-healthy diet, are associated with a reduced risk of dementia ^1^.

Hispanic older adults exhibit a disproportionate risk of Alzheimer’s and other dementias compared to non-Hispanic White counterparts, with data showing that 14% of Hispanic adults aged 65 or older have Alzheimer’s dementia, compared to 10% of non-Hispanic White older adults ^26^. Mexican Americans, the largest Hispanic population in the United States, also tend to experience a higher burden of cardiovascular risk factors, including diabetes, obesity, and dyslipidemia, which can lead to elevated rates of dementia ^27^. Mexican American men have also been shown to have a higher prevalence of cognitive impairment than non-Hispanic White men in older adults (age over 85) ^28,29^. Although recent studies have focused on looking at cardiovascular health metrics across populations, AD research still lacks diversity, with Hispanics remaining underrepresented in research ^1,16^. Furthermore, past research often used a single time point of measurement for cognitive functioning, overlooking the cumulative impact of risk factors on cognitive trajectories, particularly in differing ethnic and socioeconomic populations. To fill these gaps, our study aimed at investigating how mid-life cardiovascular health, evaluated through the LS7 scores, influences cognitive decline among Mexican American and non-Hispanic White older adults who participated in the San Antonio Heart Study (SAHS) and the San Antonio Longitudinal Study of Aging (SALSA).

## 2. Methods

### 2.1 Study Sample

SAHS was initiated in 1979 to study ethnic disparities in diabetes and cardiovascular disease ^30^. SAHS enrolled 5,158 Mexican American and non-Hispanic White adults randomly selected from socioculturally diverse neighborhoods (i.e., low-, middle-, and high-income) in San Antonio, Texas. Although Mexican American and non-Hispanic White adults were recruited from middle-income transitional neighborhoods and high-income suburban neighborhoods, only Mexican Americans were recruited from the low-income “barrio” neighborhoods, given the low number of non-Hispanic inhabitants in those areas. In 1992, 749 SAHS participants aged ≥65 years were recruited to SALSA, which aimed to study ethnic differences in the disablement process model linking chronic disease to disability ^31^. SALSA performed a baseline and three follow-up examinations over a mean of 9.5 years, during which assessments of general cognitive function were conducted using the Mini-Mental State Examination (MMSE) ^32^. This study combines mid-life information gathered during SAHS and late-life cognitive function from SALSA. Additional details about these studies can be found elsewhere ^30,33^.

### 2.2 Overview of Life’s Simple 7 Scores and Covariates

Cardiovascular health indicators were measured using the American Heart Association’s Life’s Simple 7 (LS7) criteria and coded categorically as ideal, intermediate, or poor ^7^. These included diet, physical activity, smoking, body mass index, blood pressure, cholesterol, and fasting blood glucose levels. These categories were assigned numeric scores (ideal = 2 points, intermediate = 1 point, poor = 0 points) based on their definition, which were then summed to create a composite LS7 score. Details and definitions are presented in Supplementary Table 1.

Smoking status (current, quit, or never) was assessed during the interview using the National Clearinghouse for Smoking and Health Questionnaire. Dietary habits were assessed using a food frequency questionnaire. Diet categories were constructed to reflect the available data in the study, using daily intake of vegetables, fish, and sweetened beverages to create a composite score. Physical activity was self-reported as the amount of time (hours per week) an individual spent engaging in exercise. Body mass index (BMI) was calculated during the participant’s study visit and was taken as the weight (kg) divided by the height (m) squared. Blood pressure was derived from the average of the second and third readings from three measurements. Blood samples were collected after a 12-hour fast and used to measure fasting plasma glucose and total cholesterol. Information was also collected on medications used for hyperlipidemia, diabetes, and hypertension^30^.

After categorizing the seven cardiovascular health metrics into poor, intermediate, and ideal levels, we calculated the sum for each participant’s LS7 total score, which ranged from 1 to 13.

Additional covariates used in this analysis consisted of demographic and socioeconomic indicators, including age, sex, years of formal education, monthly household income (categorized as 1=$0–49, 2=$50–99, 3=$100–149, 4=$150–199, 5=$200–299, 6=$300–399, 7=$400–499, 8=$500–749, 9=$750–999, 10=$1,000–1,249, 11=$1,250–1,499, 12=$1,500– 1,999, 13=$2,000–2,499, 14=$2,500–2,999, 15=$3,000+), recruitment neighborhood, and ethnic group^30^.

### 2.3 Statistical Analysis

Participants who had missing values for any of the LS7 categories or did not have a baseline and at least one follow-up MMSE score were excluded from the analysis (n == 347). This led to an analytical sample of 402 participants.

We conducted our analysis in various steps. First, we generated descriptive statistics for the variables of interest for the total study sample, as well as for ethnic and neighborhood subgroups. Continuous variables were described using means with standard deviations, and categorical variables by frequency and percentage. We computed a new variable characterizing the distribution of our LS7 total score by ethnicity. Tertile cutoffs were determined separately within each ethnic group to account for potential distributional differences. Participants were then categorized into three groups based on these ethnicity-specific tertiles: poor (total score 1– 6), intermediate (total score 7–8), or ideal (total score 9–13), consistent with previous studies^14,24^. Although tertiles were calculated within each ethnic group, the resulting cutoff values were consistent across groups and aligned with established thresholds from prior research^14,24^. Given the small sample sizes in some of these categories and for better interpretability of our results, we combined our intermediate and ideal categories, leaving dichotomous variables (i.e., intermediate and ideal vs. poor, Table 2).

**Table 1.** Participant characteristics at mid-life and late-life.

| **Demographics** | **Total Cohort**<br>**(n=402)** | **Mexican**<br>**American**<br>**(n=209)** | **Non-Hispanic**<br>**White**<br>**(n=193)** |
| --- | --- | --- | --- |
| **Mid-Life Measurements** |  |  |  |
| **Age (years, mean, SD)** | 57.9 (3.8) | 57.8 (3.8) | 58.0 (3.8) |
| **Female (n, %)** | 235 (58.5) | 125 (59.8) | 110 (57.0) |
| **Income (mean, SD)** | 11.9 (2.9) | 10.9 (3.1) | 13.1 (2.1) |
| **Neighborhood (n, %)** |  |  |  |
| Low-income barrio | 107 (26.6) | 107 (51.2) | 0 |
| Middle income Transitional | 45 (36.1) | 45 (21.5) | 100 (51.8) |
| High income Suburban | 150 (37.3) | 57 (27.3) | 93 (48.2) |
| **Education (years, mean, SD)** | 11.46 (4.05) | 9.6 (4.4) | 13.4 (2.5) |
| **Late-Life Measurements** |  |  |  |
| **Age (years, mean, SD)** | 69.2 (3.2) | 68.7 (2.9) | 69.8 (3.4) |
| **MMSE\* Baseline (median, IQR)** | 28.0 (25.0-29.0) | 26.0 (23.0-28.0) | 28.0 (27.0-29.0) |
| **MMSE Follow Up 1 (median, IQR)** | 27.0 (24.0-29.0) | 25.0 (22.0-28.0) | 28.0 (26.0-29.0) |
| **MMSE Follow-Up 2 (median, IQR)** | 27.0 (24.0-29.0) | 26.0 (22.0-28.0) | 28.0 (27.0-29.0) |
| **MMSE Follow Up 3 (median, IQR)** | 26.0 (24.0-29.0) | 25.0 (23.0-27.0) | 28.0 (26.0-29.0) |

\*MMSE – Mini-Mental State Examination

**Table 2.** American Heart Association’s Life’s Simple 7 (LS7) participant characteristics.

| **Health Factors** | **Total Cohort**<br>**(n=402)** | **Mexican**<br>**American**<br>**(n=209)** | **Non-Hispanic**<br>**White**<br>**(n=193)** |
| --- | --- | --- | --- |
| **BMI\* (kg/m<sup>2</sup>, median IQR)** | 26.9(24.4-30.4) | 28.2(25.7-31.7) | 25.6(23.8-29.4) |
| **BMI LS7 (n, %)** |  |  |  |
| Poor | 113(28.1) | 76(36.4) | 37(19.2) |
| Intermediate + Ideal | 289(71.9) | 133(63.6) | 156(80.8) |
| **Total Fasting Cholesterol**<br>**(mg/dL, mean, SD)** | 223(39.3) | 220.6(40.9) | 225.6(37.5) |
| **Total Fasting Cholesterol LS7 (n, %)** |  |  |  |
| Poor | 119(29.6) | 61(29.2) | 58(30.1) |
| Intermediate + Ideal | 283(70.4) | 148(70.8) | 135(69.9) |
| **Systolic Blood Pressure**<br>**(mm Hg, mean, SD)** | 123.0(15.8) | 124.8(16.4) | 121.1(15.0) |
| **Diastolic Blood Pressure**<br>**(mm Hg, mean, SD)** | 73.8(7.6) | 74.7(8.1) | 72.9(6.9) |
| **Blood Pressure LS7 (n, %)** |  |  |  |
| Poor | 61(15.2) | 39(18.7) | 22(11.4) |
| Intermediate + Ideal | 341(84.8) | 170(81.3) | 171(88.6) |
| **Fasting Blood Glucose**<br>**(mg/dL, mean, SD)** | 100.4(30.1) | 104.7(37.5) | 95.7(18.0) |
| **Fasting Blood Glucose LS7 (n, %)** |  |  |  |
| Poor | 38(10.7) | 29(13.9) | 9(4.7) |
| Intermediate + Ideal | 364(89.3) | 180(86.1) | 184(95.3) |
| **Health Behaviors** |  |  |  |
| **Smoker (n, %)** | 84(20.9) | 42(20.1) | 42(21.8) |
| **Smoking LS7 (n, %)** |  |  |  |
| Poor | 84(20.9) | 42(20.1) | 42(21.8) |
| Intermediate + Ideal | 318(79.1) | 167(79.9) | 151(78.2) |
| **Exercise (min/week, median, IQR)** | 0(0 -180) | 0(0-120) | 0(0-180) |
| **Physical Activity LS7 (n, %)** |  |  |  |
| Poor | 252(62.7) | 147(70.3) | 105(54.4) |
| Intermediate + Ideal | 150(37.3) | 62(29.7) | 88(45.6) |
| **Diet LS7 (n, %)** |  |  |  |
| Poor | 194(48.3) | 113(54.1) | 81(42.0) |
| Intermediate | 208(51.7) | 96(45.9) | 112(58.0) |
| **LS7 Total (mean, SD)** | 7.3(2.2) | 6.9(2.0) | 7.7(2.18) |
| **LS7 Tertiles (n, %)** |  |  |  |
| Poor: 1-6 | 177(44.0) | 95(45.5) | 82(42.5) |
| Intermediate: 7-8 | 140(34.8) | 70(33.5) | 70(36.3) |
| Ideal: 9-13 | 85(21.2) | 44(21.0) | 41(21.2) |

\* BMI - Body Mass Index

We implemented Generalized Estimating Equations (GEE) to assess the impact of LS7 total score and individual factors on repeated measured MMSE scores ^34^ using the gee() package in R ^35^. GEE is a robust statistical method well-suited for modeling longitudinal data, as it accounts for correlations within repeated measurements across time and provides robust estimates even when there is a departure from normality or misspecification of the correlation structure. We identified models to examine the association between individual LS7 factors, as well as the total LS7 score, with changes in cognitive function over time, using QIC to determine the optimal working correlation structure for the dataset. For the primary analysis, we investigated the interaction between LS7 score categories and time (in years, from baseline to each follow-up), adjusting for age, sex, income, education, and ethnicity (full sample models only). Analyses were performed for the total sample and stratified by ethnicity. In exploratory analyses, we followed the same approach, stratifying by neighborhood to better understand the nuances of income level on significant findings. Statistical significance was set at p-values <0.05.

## 3. Results

Table 1 shows participant characteristics at baseline in the total sample and by ethnicity. The total sample of 402 participants had an average age of 57.9 (±3.8) years at mid-life and 69.2 (±3.2) years in late life. Two hundred thirty-five (58.5%) were female, and 209 (52%) were Mexican American. The Mexican American group had fewer years of education (9.6 ± 4.4) than the non-Hispanic White group (13.4 ± 2.5). The monthly household income was also lower in the Mexican American group (10.9 ± 3.1) than in the non-Hispanic White group (13.1 ± 2.1). Median MMSE scores were lower in the Mexican American group at baseline and subsequent follow-ups.

Table 2 presents the American Heart Association’s LS7 characteristics of participants at mid-life, both in the total sample and by ethnicity. Overall, Mexican Americans tended to present with poorer cardiovascular health indicators compared to their non-Hispanic White counterparts. Mexican Americans consistently had higher percentages in the poor categories of blood glucose, blood pressure, and BMI LS7 scores (13.9%, 18.7%, and 36.4%, respectively) compared to their non-Hispanic White counterparts (4.7%, 11.4%, and 19.2%, respectively). Non-Hispanic Whites had a higher average number of minutes spent on physical activity per week and higher percentages of following an optimal diet than Mexican Americans. Overall, Mexican Americans scored lower on the LS7 total score (6.9 ± 2.0) compared to non-Hispanic Whites (7.72 ± 2.18), ranging from 1 – 13 for both groups.

We additionally examined the distribution of LS7 factors and the LS7 total score across neighborhoods and by ethnicity (Supplementary Figures 1-10). We observed that overall LS7 total scores, physical activity, and overall diet scores were higher among suburban neighborhoods, followed by transitional and barrio neighborhoods (Supplementary Figures 1, 7-8). Specific factors, including fasting blood glucose, BMI, and blood pressure measurements, were higher among Mexican Americans living in barrio neighborhoods compared to those residing in transitional or suburban neighborhoods. Similarly, non-Hispanic Whites exhibited lower BMI, blood pressure measurements, and fasting blood glucose levels than Mexican Americans, even within the same income neighborhoods (Supplementary Figures 2,3,5,6).

### 3.2 Association between LS7 and cognitive change

Association analyses between LS7 scores and cognitive change are presented in Table 3. Compared to poor categories, significant factors that slowed down the rate of cognitive decline included any physical activity, particularly in Mexican Americans, as well as fasting blood glucose levels <126 mg/dL in the non-Hispanic White cohort. Having a BMI below 30 was related to a steeper rate of cognitive decline in the Mexican American subsample.

**Table 3.**
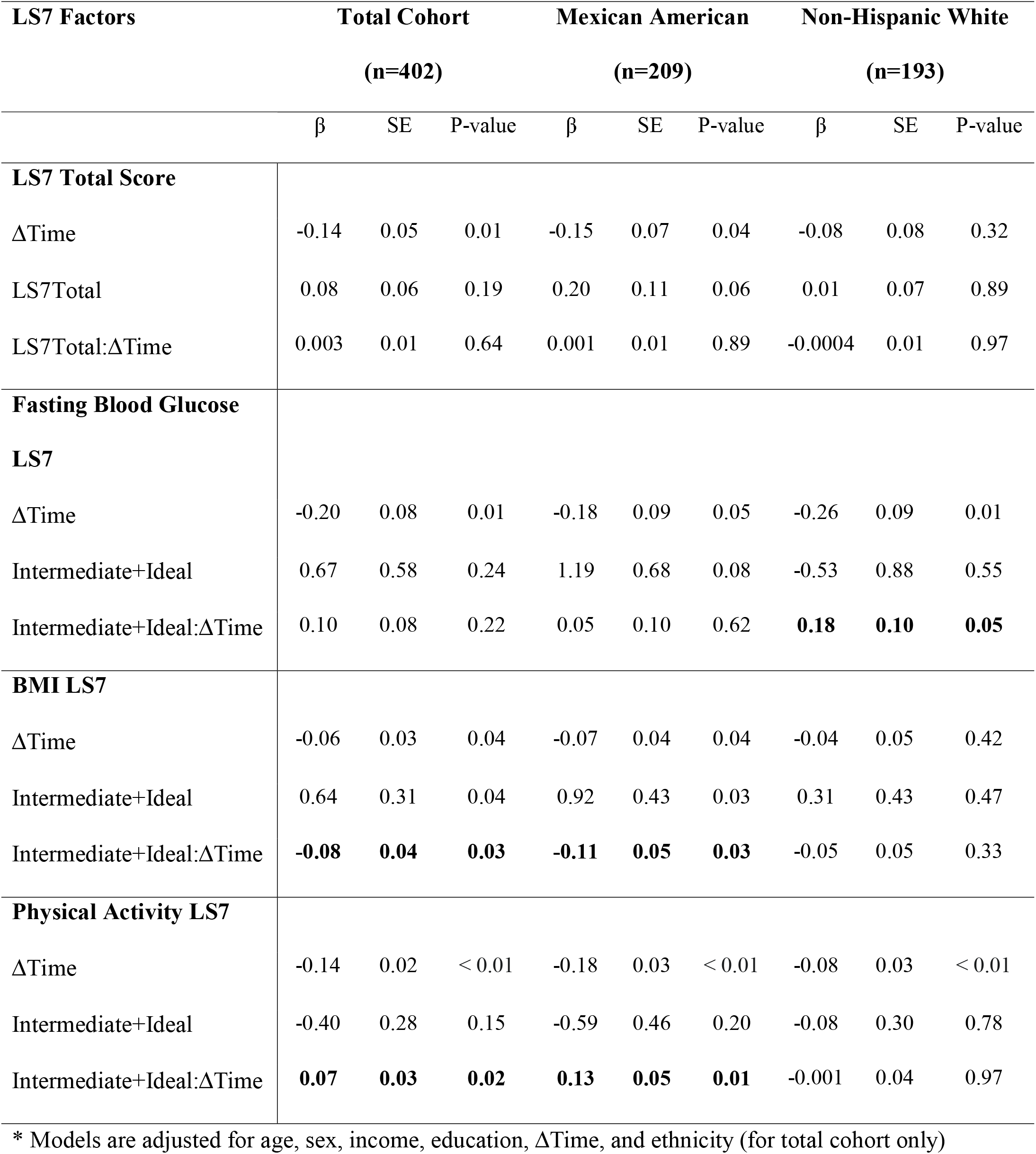
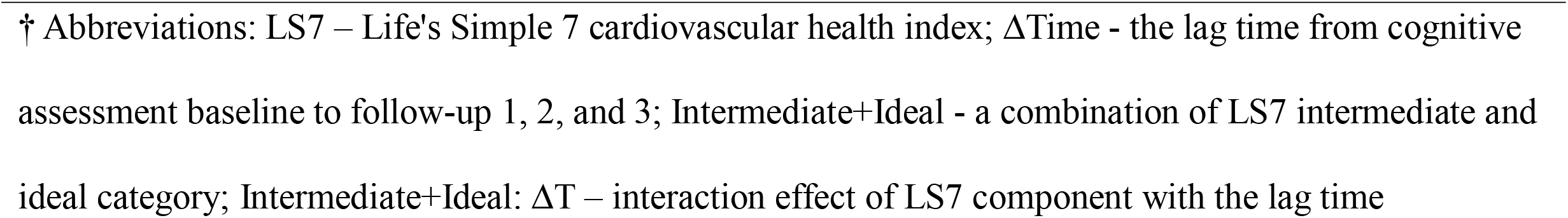
Association of cardiovascular health with cognitive change, overall and by ethnicity.

| LS7 Factors | Total Cohort |  |  | Mexican American |  |  | Non-Hispanic White |  |  |
| --- | --- | --- | --- | --- | --- | --- | --- | --- | --- |
|  | (n=402) |  |  | (n=209) |  |  | (n=193) |  |  |
| | $\beta$ | SE | P-value | $\beta$ | SE | P-value | $\beta$ | SE | P-value |
| **LS7 Total Score** |  |  |  |  |  |  |  |  |  |
| ΔTime | -0.14 | 0.05 | 0.01 | -0.15 | 0.07 | 0.04 | -0.08 | 0.08 | 0.32 |
| LS7Total | 0.08 | 0.06 | 0.19 | 0.20 | 0.11 | 0.06 | 0.01 | 0.07 | 0.89 |
| LS7Total:ΔTime | 0.003 | 0.01 | 0.64 | 0.001 | 0.01 | 0.89 | -0.0004 | 0.01 | 0.97 |
| **Fasting Blood Glucose LS7** |  |  |  |  |  |  |  |  |  |
| ΔTime | -0.20 | 0.08 | 0.01 | -0.18 | 0.09 | 0.05 | -0.26 | 0.09 | 0.01 |
| Intermediate+Ideal | 0.67 | 0.58 | 0.24 | 1.19 | 0.68 | 0.08 | -0.53 | 0.88 | 0.55 |
| Intermediate+Ideal:ΔTime | 0.10 | 0.08 | 0.22 | 0.05 | 0.10 | 0.62 | **0.18** | **0.10** | **0.05** |
| **BMI LS7** |  |  |  |  |  |  |  |  |  |
| ΔTime | -0.06 | 0.03 | 0.04 | -0.07 | 0.04 | 0.04 | -0.04 | 0.05 | 0.42 |
| Intermediate+Ideal | 0.64 | 0.31 | 0.04 | 0.92 | 0.43 | 0.03 | 0.31 | 0.43 | 0.47 |
| Intermediate+Ideal:ΔTime | **-0.08** | **0.04** | **0.03** | **-0.11** | **0.05** | **0.03** | -0.05 | 0.05 | 0.33 |
| **Physical Activity LS7** |  |  |  |  |  |  |  |  |  |
| ΔTime | -0.14 | 0.02 | < 0.01 | -0.18 | 0.03 | < 0.01 | -0.08 | 0.03 | < 0.01 |
| Intermediate+Ideal | -0.40 | 0.28 | 0.15 | -0.59 | 0.46 | 0.20 | -0.08 | 0.30 | 0.78 |
| Intermediate+Ideal:ΔTime | **0.07** | **0.03** | **0.02** | **0.13** | **0.05** | **0.01** | -0.001 | 0.04 | 0.97 |

\* Models are adjusted for age, sex, income, education, $\Delta$ Time, and ethnicity (for total cohort only)
† Abbreviations: LS7 – Life's Simple 7 cardiovascular health index; $\Delta$ Time - the lag time from cognitive assessment baseline to follow-up 1, 2, and 3; Intermediate+Ideal - a combination of LS7 intermediate and ideal category; Intermediate+Ideal: $\Delta$ T – interaction effect of LS7 component with the lag time

In the total cohort, meeting at least intermediate physical activity goals mitigated cognitive decline by 0.073 points per year on the MMSE (Figure 1a). These results seemed to be driven by Mexican American participants, in whom any physical activity attenuated cognitive decline by 0.127 points per year on the MMSE compared to no physical activity (Figure 1b). No significant results were observed between physical activity and cognitive change among non-Hispanic White participants in the stratified results.

**Figure 1.**
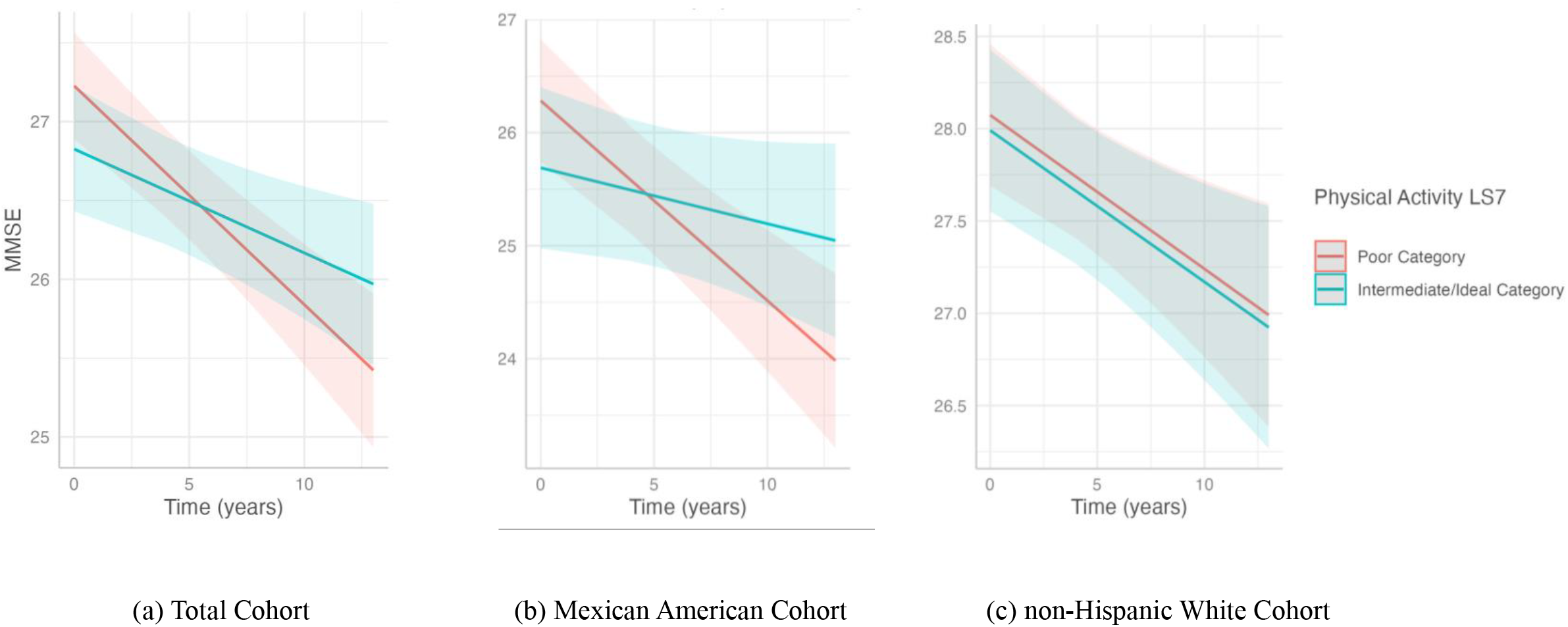
Association of physical activity with cognitive decline in the (a) total cohort, (b) Mexican American cohort, and the (c) non-Hispanic White cohort. This plot shows the slope of MMSE over time predicted by the interaction with physical activity LS7 categories, poor and a combination of intermediate and ideal, with cognitive decline. The poor physical activity LS7 category included individuals who reported 0 minutes of exercise a week. Intermediate/ideal physical activity included individuals who recorded any (>0) exercise minutes a week. Predicted outcomes and confidence interval bands are shown, with red representing the poor physical activity LS7 category and blue the intermediate/ideal category. In the analysis, only the (a) total cohort and (b) Mexican American cohort showed significant differences between the slope of the poor physical activity LS7 category and the intermediate/ideal category. Total Cohort (b) Mexican American Cohort (c) non-Hispanic White Cohort

Maintaining fasting blood glucose levels <126 mg/dL was associated with slower cognitive decline by 0.182 points per year among non-Hispanic Whites (Figure 2). However, no significant association was found between fasting blood glucose and cognitive decline in the Mexican American group.

**Figure 2.**
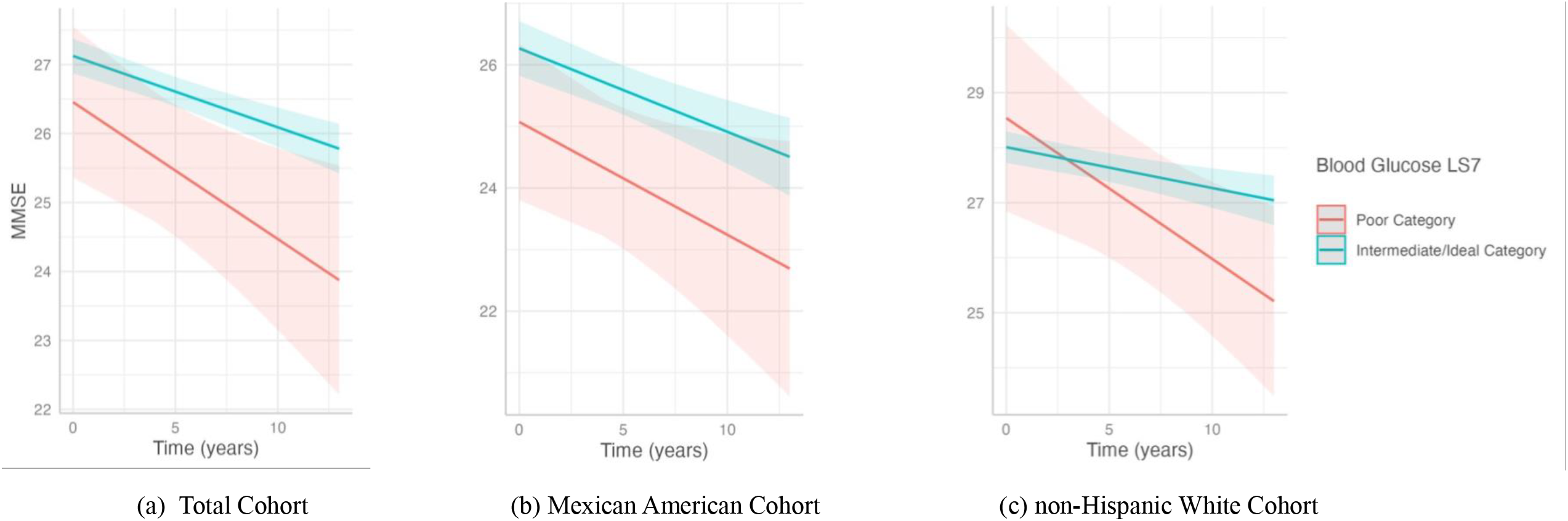
Association of blood glucose with cognitive decline in the (a) total cohort, (b) Mexican American cohort, and (c) non-Hispanic White cohort. This plot shows the slope of MMSE over time predicted by the interaction of the two LS7 blood glucose categories, poor and a combination of intermediate and ideal, with cognitive decline. The poor blood glucose LS7 category included individuals with a fasting blood glucose ≥ 126 mg/dL. Intermediate/ideal category included individuals with a fasting blood glucose < 125 mg/dL or < 100 mg/dL if treated. Predicted outcomes and confidence interval bands are shown, with red representing the poor blood glucose LS7 category and blue the intermediate/ideal category. The significant results from our analysis were in the (c) non-Hispanic White cohort.

Our findings also suggest that participants with a BMI between 18 and 30 kg/m^2^ had an accelerated cognitive decline of 0.078 points per year on the MMSE in the total cohort compared to those classified as obese. After stratification, the decline was more evident among Mexican Americans, who experienced a drop of 0.114 points per year (Figure 3). In the non-Hispanic White group, a similar pattern was observed, but it was not statistically significant. We conducted two additional exploratory analyses to further investigate these findings. First, we repeated the analysis stratifying by the sample median age (58 years) based on literature describing excess weight as a dementia risk factor in midlife, but an apparent protective factor in late life ^36^. Results showed that in the total cohort, individuals under 58 had a negative correlation with better LS7 BMI scores, with those with a BMI between 18 and 30 kg/m^2^ having a steeper cognitive decline of 0.102. The Mexican American cohort was not different across age strata (Supplementary Table 3). Second, we re-defined the exposure to integrate obesity and metabolic health, given that not all individuals classified as obese experience the same cardiometabolic risk (Supplementary Table 4) ^37-40^. Compared to metabolically unhealthy obese, Mexican American participants classified as metabolically healthy non-obese, metabolically healthy obese, or metabolically unhealthy non-obese showed a significant decline of 0.107 points in MMSE score (Supplementary Table 5). No significant associations were observed when these exposure categories were analyzed separately (Supplementary Table 6). We did not observe additional significant associations between cognitive change with continuous total LS7 scores, LS7 tertiles, or other cardiovascular health factors (Supplementary Table 2, Supplementary Figures 11-21).

**Figure 3.**
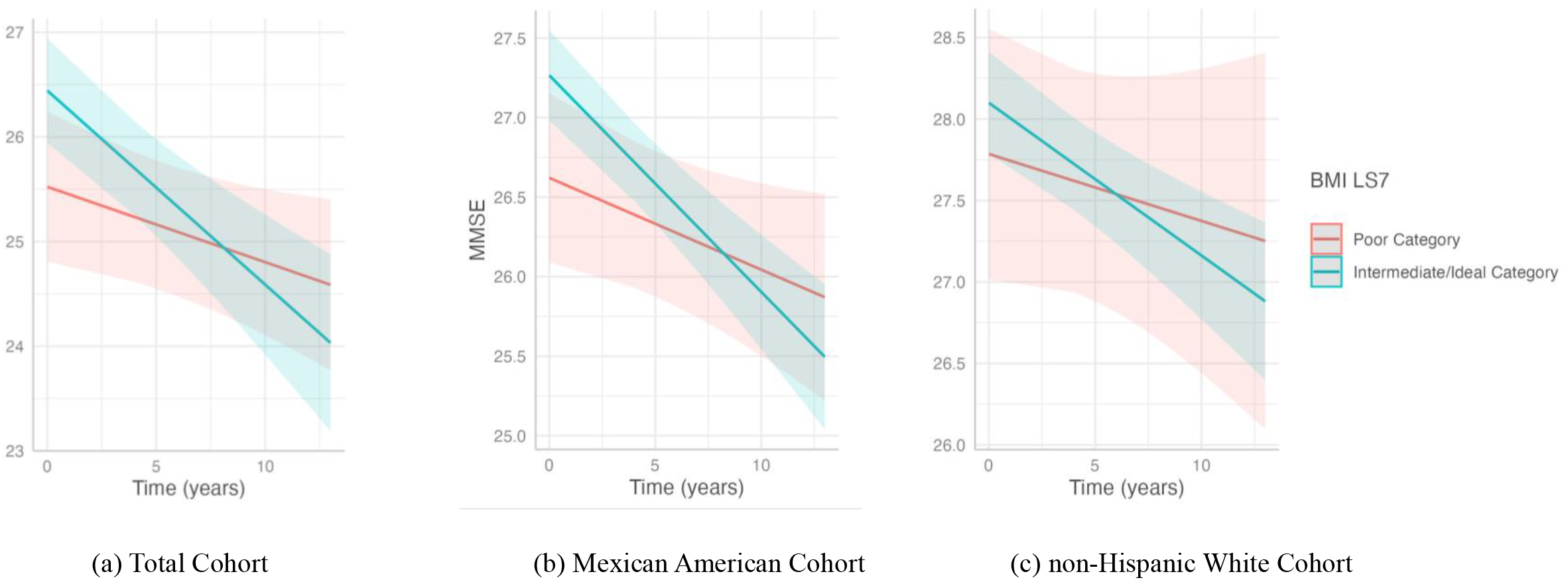
Association of BMI with cognitive decline in the (a) total cohort, (b) Mexican American cohort, and (c) non-Hispanic White cohort. This plot shows the slope of MMSE predicted over time by the interaction of BMI LS7 categories, poor and a combination of intermediate and ideal, with cognitive decline. The poor BMI LS7 category consisted of individuals with a BMI ≥30 kg/m^2^. Intermediate/ideal category included individuals with a BMI < 30 kg/m^2^.Predicted outcomes and confidence interval bands are shown, with red representing the poor BMI LS7 category and blue the intermediate/ideal category. Significant results from our analysis were in the (a) total cohort and (b) Mexican American cohort.

### 3.3 Secondary Analysis by Neighborhood Stratification

In the GEE analysis, we further stratified our significant results (p < 0.05) by neighborhood: barrio, transitional, and suburban (with only Mexican Americans recruited from barrio neighborhoods). Among Mexican American individuals, those residing in the ‘barrio’ neighborhood with an ideal or intermediate BMI (18 to 30 kg/m^2^) experienced an accelerated cognitive decline of 0.168 MMSE points per year, whereas those in suburban neighborhoods showed a decline of 0.205 (Supplementary Table 7). Mexican Americans in barrio neighborhoods demonstrated a slower decline of 0.214 when moderate physical activity was present. No significant associations were found for Mexican Americans in transitional neighborhoods (Supplementary Table 7). Within the non-Hispanic White group, individuals in the suburban neighborhoods had a significant association with fasting blood glucose levels below 126 mg/dL, with a slower decline of 0.247 associated with optimal blood glucose (Supplementary Table 7).

These findings highlight the potential role of socioeconomic status, represented by neighborhood type, in moderating the association between LS7 factors and cognitive decline. We observed that certain LS7 factors, such as physical activity and fasting blood glucose levels, had an impact on cognitive trajectories across different neighborhood types.

## 4. Discussion

This study examined the association of midlife LS7 scores and its factors with cognitive trajectories in a bi-ethnic cohort, and explored whether these differed between Mexican Americans and non-Hispanic Whites. Although total LS7 scores did not correlate significantly with cognitive change, the impact of LS7 individual factors varied between ethnic groups. In Mexican American participants, any physical activity mitigated cognitive decline, whereas those classified as normal or overweight experienced an accelerated cognitive decline. Additionally, optimal blood glucose levels were linked to attenuated cognitive decline in non-Hispanic Whites.

Several studies have investigated the impact of multiple cardiovascular health dimensions on cognitive function. Still, only a handful have focused on Hispanic samples despite their increased risk for cardiometabolic disease and earlier cognitive decline ^26,29^. An analysis from the Health and Aging Brain Study: Health Disparities ^41^ reported distinct associations between cardiovascular risk factors and cognitive function in Mexican Americans and Non-Hispanic Whites. However, those patterns were not replicated in our findings. For instance, in the study, cardiovascular disease, higher abdominal circumference, and smoking were related to lower cognitive function in non-Hispanic Whites, whereas only higher abdominal circumference and smoking were associated with poorer memory in Mexican Americans. These differences could be due to study designs, as theirs was cross-sectional. Investigations from the Hispanic Community Health Study/Study of Latinos (HCHS/SOL), which includes participants of diverse Hispanic backgrounds, suggest that optimal LS7 scores in middle- and older-aged Hispanics/Latinos (age 45 – 74) are associated with better cognitive function ^24^. Similar analyses in HCHS/SOL have shown that the cumulative burden of cardiovascular risk factors (i.e., hypertension, diabetes, smoking, and obesity) and indicators of arterial stiffness correlate with worse cognitive function ^42,43^. The 10/66 Dementia Research Group further showed that optimal cardiovascular health inversely relates to dementia incidence among Latin American adults aged 65 and older, with moderate and ideal levels of cardiovascular health associated with lower dementia incidence ^25^. The Mexico-SAGE project, which included a sample of older Mexican adults, showed that ideal cardiovascular health factors were associated with better cognitive function in those aged 50-64 years old ^44^.

Existing literature has also shown that adherence to LS7’s ideal cardiovascular health recommendations during mid-life is associated with a reduced risk of dementia in later life, reinforcing the importance of studying the relationship between cardiovascular health during mid-life and cognitive aging. For example, the Brazilian Longitudinal Study of Adult Health (baseline mean age 51.4) found that higher Life’s Essential 8 scores, an updated composite that adds sleep health to the LS7, were associated with slower decline in global cognition, memory, verbal fluency, and executive functioning over eight years of follow-up ^45^. The Honolulu Asia Aging Study (baseline mean age of 52) revealed that participants with mid-life systolic blood pressure above 160 mmHg had an increased risk of dementia 25 years later compared to those with systolic blood pressure between 110 and 139 mmHg^46^. In the same cohort, those with ideal cardiovascular health behaviors were less likely to develop dementia over the following 25 years, with higher BMI (> 25.0 kg/m^2^) being a strong predictor of dementia risk ^47^. In the Framingham Heart Study, a higher burden of cardiovascular risk factors was associated with reduced brain volume, particularly in younger individuals (mean age 55) ^48^. Longitudinal data from the Atherosclerosis Risk in Communities study supported these findings, showing that higher mid-life (mean age of 54) LS7 scores were associated with better global cognition, which persisted over 20 years ^22^. The Northern Manhattan Study, which included a large proportion of Caribbean Hispanic participants, reported a positive association between the number of ideal scores for cardiovascular health factors with less decline in processing speed ^19^.

Our findings align with previous research underscoring the importance of physical activity as an essential lifestyle factor in mitigating cognitive decline in later life ^13,16,49^. Our findings also support the World Health Organization’s guidelines on risk reduction for cognitive decline and dementia, which recommend physical activity for adults with normal cognition to reduce the risk of cognitive decline ^50^. Potential mechanisms through which physical activity counteracts cognitive decline include molecular mechanisms such as the activation of insulin-like growth factor, which promotes both synaptogenesis and angiogenesis, and increasing brain-derived neurotrophic factor levels, crucial for neurogenesis and synaptogenesis ^51^. Angiogenesis, the increase in blood flow to the brain, is suggested to be induced by physical activity and may help mitigate cognitive decline associated with aging ^51^. Neurogenesis studies have focused on identifying stimuli that modulate the proliferation and survival of newly formed neurons, with physical activity shown to positively impact this process ^51^. Experiments in animal models demonstrate that aerobic exercise stimulates hippocampal neurogenesis, facilitating the induction and maintenance of long-term potentiation and enhancing short-term memory ^51^.

Our results indicating that having optimal fasting blood glucose relates to better cognition are supported by previous studies suggesting that the toxic effects of hyperglycemia can lead to slow, progressive function and structural abnormalities in the brain ^52^, with a large body of evidence showing the association between diabetes and cognitive decline and dementia ^52-55^. Moreover, consistently high blood glucose levels have been found to affect cognitive performance in otherwise healthy individuals and could be implicated in the underlying neuropathological processes associated with cognitive decline ^52,56^. However, it is unclear why these results were only observed among non-Hispanic Whites. The Project Facing Rural Obstacles to health Now Through Intervention, Education & Research study reported associations between diabetes or higher HbA1c levels and MCI in non-Hispanic White participants, but not among Mexican Americans ^57^, similar to our results. However, these analyses were cross-sectional, and exposures were measured at different ages.

Conversely, Mexican American participants with intermediate and ideal BMI experienced a steeper cognitive decline. These results were largely unchanged in exploratory analyses stratifying by age or re-defining the exposure by categories of obesity and metabolic health. Epidemiological studies have reported conflicting findings on the role of BMI in cognitive aging ^36^. Whereas midlife obesity has been related to an increased risk of late-onset dementia, a higher BMI in older ages has been linked to both increased and decreased risk of dementia ^58-61^. These differences may be explained by the age at when BMI is measured and the cognitive status of the sample. A meta-analysis of more than 1.3 million participants suggests that a higher BMI measured more than 20 years before dementia onset is associated with increased dementia risk, but the opposite is seen when BMI is measured only 10 years before onset ^62^. This may be explained by reverse causality, given that accelerated weight loss can occur during the years preceding MCI and dementia onset ^63^. Because Mexican Americans are reported to experience cognitive impairment at younger ages compared to non-Hispanic Whites^64^, it is conceivable that the reverse causality phenomenon occurs earlier in this group. Although not entirely clear, potential mechanisms linking excess weight in midlife to cognitive decline may involve the promotion of chronic low-grade inflammation, metabolic dysregulation, and oxidative stress processes leading to an increased cardiovascular burden and neurodegeneration ^65^.

### 4.1 Strengths and limitations

The strengths of our work include the study of a representative sample of Mexican American and non-Hispanic White adults from diverse socioeconomic areas in the South Texas region, with a comprehensive set of sociodemographic and cardiovascular risk factors collected during mid-life and the longitudinal collection of cognitive data during late life. Additionally, we utilized robust analytical techniques to handle the non-parametric measures and misspecifications of correlations in our data. However, our study also has some limitations. First, our sample size was relatively small, potentially limiting the power to detect associations, especially by ethnic/neighborhood subgroups. Second, participant recall bias, particularly regarding health behaviors such as physical exercise and diet, may have affected data accuracy. Third, although our LS7 index closely matched the American Heart Association guidelines, the analytical strategies used to fit health measure patterns inherent to our sample could affect comparability with other studies.

### 4.2 Conclusions

Our findings suggest that maintaining optimal fasting glucose and engaging in any level of physical activity in midlife can significantly reduce the rate of cognitive decline. However, these results differed between Mexican American and non-Hispanic White participants. Further research is needed in larger samples to verify and replicate these differences. The longitudinal exploration of cardiovascular health throughout life, rather than assessing it at a single time point, could also provide further insight into the trajectory phenotypes contributing to cognitive decline by ethnicity. This last point is particularly important to understand the impact of excess weight on Mexican Americans. By improving cardiovascular health through modifiable lifestyle changes, there is a potential to significantly alleviate the public health burden of Alzheimer’s and other dementias, particularly in high-risk populations.

## Data Availability

All data produced in the present study are available upon reasonable request to the authors.

## Acknowledgments

We would like to thank the participants who contributed to the San Antonio Heart Study (SAHS) and the San Antonio Longitudinal Study of Aging (SALSA).

## Disclosures

There are no competing interests to declare.

## Funding

This work was supported by the National Institutes of Health T32 training grant - Pathobiology of Occlusive Vascular Disease (T32 HL007446), the San Antonio Heart and Mind Study (R01 AG082360), San Antonio Longitudinal Study of Aging (R01 AG010444, R01 AG016518), and the Burroughs Wellcome Fund Postdoctoral Diversity Enrichment Program. Dr. Satizabal receives additional support from NINDS (UH1 NS125513). Drs. Satizabal, Wang, Melgarejo, Maestre, and Seshadri are partially supported by the South Texas Alzheimer’s Disease Research Center (P30 AG066546). Dr. Seshadri receives support from The Bill and Rebecca Reed Endowment for Precision Therapies and Palliative Care and an endowment from the Barker Foundation as the Robert R Barker Distinguished University Professor of Neurology, Psychiatry and Cellular and Integrative Physiology.

## Notes

### Competing Interest Statement

The authors have declared no competing interest.

### Author Declarations

The study was approved by the Institutional Review Board of the University of Texas Health Science Center at San Antonio, and all subjects gave informed consent.

