## Supplementary Material for "Impact of mid-life cardiovascular health on cognitive change in a bi-ethnic cohort"

Supplementary Tables and Figures

[Supplementary Table 7. Association of fasting blood glucose LS7, BMI LS7, and physical activity LS7 scores with cognitive change, stratified by ethnicity and neighborhood for previously reported significant interaction results (p < 0.05)	7](#_Toc478737288)

### Supplementary Table 1. Definition of cardiovascular health metrics based on the American Heart Association's LS7 1

| **Goal/Metric** | **Poor Health**  **(score = 0)** | **Intermediate Health**  **(score = 1)** | **Ideal Health**  **(score = 2)** |
| --- | --- | --- | --- |
| **Blood pressure** | SBP ≥ 140 or SBP < 90  DBP ≥ 90 mmHg or DBP < 60 | SBP 120-139 or DBP 80-89 mmHg or  SBP < 120 and DBP < 80 mmHg treated | SBP < 120 and DBP < 80 mmHg untreated |
| **Total fasting cholesterol** | ≥ 240 mg/dL | 200-239 mg/dL or < 200 mg/dL treated | < 200 mg/dL untreated |
| **Fasting blood glucose** | ≥ 126 mg/dL | Glucose: 100-125 mg/dL or < 100 mg/dL treated | 70 mg/dL - 100 mg/dL untreated |
| **Body mass index (BMI)** | ≥30 kg/m^2^ | 25-30 kg/m^2^ | 18.0-25.0 kg/m^2^ |
| **Smoking** | Current smoker | Former smoker | Never smoked |
| **Diet**  Based on the eating frequency for the following three components:  1) Fruit/Salad  - 1 serving a day coded as 1  - never or less than 1 serving a day coded as 0  2) Fish  - 2 servings a week coded as 1  - never or less than 2 servings a week coded as 0  3) Sweetened Beverages  - less than 36 oz a week coded as 1  - more than 36 oz a week coded as 0 | Total score of 0 or 1 component (0-1) | Total score of 2 or 3 components (2-3) |  |
| **Physical activity**  Based on Moderate Intensity minutes of activity/week (only) | Never | 1-150 min/week moderate or vigorous | >150 min/week moderate or vigorous |

**Abbreviations: SBP – systolic blood pressure; DBP – diastolic blood pressure*

### Supplementary Table 2. Association of LS7 tertile, LS7 factors, and cardiovascular health measurements with cognitive change, overall and by ethnicity

|  | **Total Cohort (n=402)** | | | | **Mexican American (n=209)** | | | | **Non-Hispanic White (n=193)** | | |
| --- | --- | --- | --- | --- | --- | --- | --- | --- | --- | --- | --- |
|  | **β** | | **SE** | **p-value** | **β** | **SE** | **p-value** | **β** | | **SE** | **p-value** |
| **LS7 Category (by Tertiles)** |  |  |  |  |  |  |  |  |  |  |  |
| ΔTime | -0.12 | | 0.02 | < 0.01 | -0.16 | 0.03 | < 0.01 | -0.07 | | 0.03 | 0.04 |
| Intermediate | 0.31 | | 0.28 | 0.28 | 0.32 | 0.45 | 0.48 | 0.30 | | 0.32 | 0.36 |
| Ideal | 0.59 | | 0.31 | 0.08 | 0.13 | 0.54 | 0.04 | 0.08 | | 0.38 | 0.84 |
| Intermediate:ΔTime | -0.02 | | 0.04 | 0.68 | 0.06 | 0.07 | 0.34 | -0.03 | | 0.05 | 0.44 |
| Ideal:ΔTime | -0.02 | | 0.04 | 0.70 | -0.01 | 0.06 | 0.88 | -0.02 | | 0.05 | 0.64 |
| **Blood Pressure** |  |  |  |  |  |  |  |  |  |  |  |
| ΔTime | -0.08 | | 0.04 | 0.05 | -0.12 | 0.06 | 0.03 | -0.03 | | 0.06 | 0.63 |
| Intermediate+Ideal | 0.08 | | 0.35 | 0.82 | 0.24 | 0.49 | 0.63 | -0.04 | | 0.50 | 0.94 |
| Intermediate+Ideal:ΔTime | -0.04 | | 0.05 | 0.45 | -0.03 | 0.06 | 0.64 | -0.06 | | 0.06 | 0.35 |
| **Total Cholesterol** |  |  |  |  |  |  |  |  |  |  |  |
| ΔTime | -0.09 | | 0.03 | < 0.01 | -0.11 | 0.04 | 0.01 | -0.08 | | 0.04 | 0.04 |
| Intermediate+Ideal | 0.32 | | 0.27 | 0.25 | 0.37 | 0.43 | 0.39 | 0.22 | | 0.31 | 0.49 |
| Intermediate+Ideal:ΔTime | -0.04 | | 0.03 | 0.31 | -0.05 | 0.05 | 0.32 | -0.01 | | 0.04 | 0.86 |
| **Smoking** |  |  |  |  |  |  |  |  |  |  |  |
| ΔTime | -0.13 | | 0.03 | < 0.01 | -0.14 | 0.05 | < 0.01 | -0.13 | | 0.05 | 0.01 |
| Intermediate+Ideal | 0.31 | | 0.31 | 0.31 | 0.45 | 0.49 | 0.36 | 0.21 | | 0.36 | 0.56 |
| Intermediate+Ideal:ΔTime | 0.02 | | 0.04 | 0.63 | -0.01 | 0.06 | 0.88 | 0.06 | | 0.05 | 0.30 |
| **Diet** |  |  |  |  |  |  |  |  |  |  |  |
| ΔTime | -0.10 | | 0.02 | < 0.01 | -0.15 | 0.04 | < 0.01 | -0.05 | | 0.03 | 0.09 |
| Intermediate+Ideal | -0.08 | | 0.26 | 0.73 | -0.21 | 0.41 | 0.61 | 0.17 | | 0.28 | 0.55 |
| Intermediate+Ideal:ΔTime | -0.02 | | 0.03 | 0.60 | 0.01 | 0.06 | 0.84 | -0.06 | | 0.04 | 0.12 |
| **Fasting blood glucose** |  |  |  |  |  |  |  |  |  |  |  |
| ΔTime | -0.02 | | 0.06 | 0.74 | -0.07 | 0.07 | 0.34 | 0.03 | | 0.11 | 0.77 |
| Fasting blood glucose | -0.003 | | 0.004 | 0.55 | -0.01 | 0.01 | 0.20 | 0.01 | | 0.01 | 0.07 |
| Fasting blood glucose:ΔTime | -0.001 | | 0.001 | 0.13 | -0.001 | 0.001 | 0.30 | -0.001 | | 0.001 | 0.29 |
| **Systolic blood pressure** |  |  |  |  |  |  |  |  |  |  |  |
| ΔTime | -0.11 | | 0.12 | 0.34 | -0.15 | 0.17 | 0.40 | -0.15 | | 0.16 | 0.36 |
| Systolic blood pressure | -0.01 | | 0.01 | 0.30 | -0.02 | 0.01 | 0.10 | 0.001 | | 0.01 | 0.88 |
| Systolic blood pressure:ΔTime | -0.00001 | | 0.001 | 0.99 | 0.00004 | 0.001 | 0.98 | 0.001 | | 0.001 | 0.69 |
| **Diastolic blood pressure** |  |  |  |  |  |  |  |  |  |  |  |
| ΔTime | -0.28 | | 0.15 | 0.06 | -0.34 | 0.22 | 0.12 | -0.30 | | 0.20 | 0.13 |
| Diastolic blood pressure | 0.001 | | 0.02 | 0.95 | -0.01 | 0.02 | 0.69 | 0.003 | | 0.02 | 0.87 |
| Diastolic blood pressure:ΔTime | 0.002 | | 0.002 | 0.24 | 0.003 | 0.003 | 0.36 | 0.003 | | 0.003 | 0.27 |
| **Total cholesterol** |  |  |  |  |  |  |  |  |  |  |  |
| ΔTime | -0.22 | | 0.10 | 0.02 | -0.26 | 0.14 | 0.07 | -0.14 | | 0.12 | 0.25 |
| Total cholesterol | -0.004 | | 0.004 | 0.29 | -0.004 | 0.01 | 0.52 | -0.004 | | 0.004 | 0.35 |
| Total cholesterol:ΔTime | 0.0005 | | 0.0004 | 0.25 | 0.001 | 0.001 | 0.39 | 0.0002 | | 0.001 | 0.66 |
| **BMI** |  |  |  |  |  |  |  |  |  |  |  |
| ΔTime | -0.25 | | 0.09 | 0.006 | -0.33 | 0.15 | 0.03 | -0.24 | | 0.13 | 0.07 |
| BMI | -0.06 | | 0.03 | 0.06 | -0.09 | 0.05 | 0.06 | -0.03 | | 0.03 | 0.38 |
| BMI:ΔTime | 0.005 | | 0.003 | 0.13 | 0.01 | 0.005 | 0.18 | 0.01 | | 0.005 | 0.24 |
| **Exercise (minutes/week)** |  |  |  |  |  |  |  |  |  |  |  |
| ΔTime | -0.13 | | 0.02 | < 0.01 | -0.17 | 0.03 | < 0.01 | -0.08 | | 0.02 | < 0.01 |
| Exercise | -0.0005 | | 0.001 | 0.54 | -0.001 | 0.001 | 0.29 | 0.0001 | | 0.001 | 0.88 |
| Exercise:ΔTime | 0.0002 | | 0.0001 | 0.06 | **0.0004** | **0.0001** | **0.01** | -0.00004 | | 0.0001 | 0.64 |
| *Models adjusted for age, sex, income, education, ΔTime, and ethnicity (for total cohort only)  † Abbreviations: LS7–Life's Simple 7 cardiovascular health index; ΔTime - the lag time from cognitive assessment baseline to follow-up 1, 2, and 3; Intermediate+Ideal - combination of LS7 intermediate and ideal category; Intermediate+Ideal:ΔT – interaction effect of LS7 component with the lag time | | | | | | | | | | | |

### Supplementary Table 3. Association between LS7 BMI categories and cognitive change stratified by age, overall and by ethnicity

|  | | **Total Cohort (n=183)** | | | | **Mexican American (n=97)** | | | **Non-Hispanic White (n=86)** | | |
| --- | --- | --- | --- | --- | --- | --- | --- | --- | --- | --- | --- |
| **< 58 years** | | **β** | | **SE** | **p-value** | **β** | **SE** | **p-value** | **β** | **SE** | **p-value** |
| ΔTime | | -0.01 | | 0.04 | 0.76 | -0.03 | 0.06 | 0.61 | 0.005 | 0.07 | 0.94 |
| Intermediate+Ideal | | 0.83 | | 0.50 | 0.10 | 1.10 | 0.67 | 0.10 | 0.52 | 0.71 | 0.46 |
| Intermediate+Ideal:ΔTime | | **-0.10** | | **0.05** | **0.04** | -0.12 | 0.07 | 0.09 | -0.09 | 0.07 | 0.22 |
|  | | **Total Cohort (n=219)** | | | | **Mexican American (n=112)** | | | **Non-Hispanic White (n=107)** | | |
| **>= 58 years** | | **β** | | **SE** | **p-value** | **β** | **SE** | **p-value** | **β** | **SE** | **p-value** |
| ΔTime | | -0.08 | | 0.04 | 0.02 | -0.10 | 0.05 | 0.03 | -0.08 | 0.08 | 0.33 |
| Intermediate+Ideal | | 0.52 | | 0.40 | 0.20 | 0.83 | 0.58 | 0.15 | 0.14 | 0.50 | 0.78 |
| Intermediate+Ideal:ΔTime | | -0.07 | | 0.05 | 0.16 | -0.13 | 0.08 | 0.10 | -0.03 | 0.08 | 0.76 |
|  | *Models adjusted for age, sex, income, education, ΔTime, and ethnicity (for total cohort only)  † Abbreviations: LS7–Life's Simple 7 cardiovascular health index; ΔTime - the lag time from cognitive assessment baseline to follow-up 1, 2, and 3; Intermediate+Ideal - combination of LS7 intermediate and ideal category; Intermediate+Ideal:ΔT – interaction effect of LS7 component with the lag time | | | | | | | | | | |

### Supplementary Table 4. Definition of metabolic health status categories: metabolic health normal (MHN), metabolic unhealthy normal (MUN), metabolic health obesity (MHO), and metabolic unhealthy obesity (MUO)2 and their distribution by ethnicity

| **Metabolic Health Status** | **Metabolically Health Normal (MHN)** | **Metabolically Unhealthy Normal (MUN)** | **Metabolically Healthy Obese (MHO)** | **Metabolically Unhealthy Obese (MUO)** |
| --- | --- | --- | --- | --- |
| **Fasting Triglycerides** | <150 mg/dL | ≥150 mg/dL or treated | < 50 mg/dL | ≥150 mg/dL or treated |
| **High Density Lipoprotein Cholesterol** | ≥40 mg/dL in men or ≥50 mg/dL in women | <40 mg/dL in men or <50 mg/dL in women or treated | ≥40 mg/dL in men or ≥50 mg/dL in women | <40 mg/dL in men or <50 mg/dL in women or treated |
| **Blood Pressure** | Systolic <130 mm Hg and/or  diastolic <85 mm Hg | Systolic ≥130 mm Hg and/or  diastolic ≥85 mm Hg or treated | Systolic <130 mm Hg and/or  diastolic <85 mm Hg | Systolic ≥130 mm Hg and/or  diastolic ≥85 mm Hg  or treated |
| **Fasting Blood Glucose** | < 100 mg/dL | ≥100 mg/dL or treated | <100 mg/dL | ≥100 mg/dL or treated |
| **BMI** | < 30 kg/m^2^ | | ≥ 30 kg/m^2^ | |
| **Distribution of metabolic health status** | | | | |
| Ethnicity (n, %)  Mexican American  Non-Hispanic White | 36 (9)  44 (10.9) | 97 (24.1)  112 (27.9) | 5 (1.2)  4 (1) | 71 (17.7)  33 (8.2) |

**Abbreviations: BMI - body mass index*

### Supplementary Table 5. Association of metabolic status (MUO vs. MUN, MHO, MHN) with cognitive change, overall and by ethnicity

|  | **Total Cohort (n = 402)** | | | **Mexican American (n=209)** | | | **Non-Hispanic White (n=193)** | | |
| --- | --- | --- | --- | --- | --- | --- | --- | --- | --- |
|  | **β** | **SE** | **p-value** | **β** | **SE** | **p-value** | **β** | **SE** | **p-value** |
| ΔTime | -0.07 | 0.03 | 0.03 | -0.07 | 0.04 | 0.05 | -0.06 | 0.05 | 0.23 |
| MUN,MHO,MHN | 0.59 | 0.32 | 0.06 | 1.08 | 0.43 | 0.01 | -0.15 | 0.40 | 0.71 |
| ΔTime:MUN,MHO,MHN | -0.06 | 0.04 | 0.08 | **-0.11** | **0.05** | **0.04** | 0.02 | 0.06 | 0.69 |

**Models were adjusted for age, sex, income, education,* *ΔTime, and ethnicity (for total cohort only) with metabolically unhealthy obese as the reference*

*† Abbreviations: MUO – metabolically unhealthy obese (reference category); MUN – metabolically unhealthy normal; MHO – metabolically healthy obese; MHN – metabolically healthy normal; ΔTime - the lag time from cognitive assessment baseline to follow-up 1, 2, and 3; ΔTime:MUN – interaction effect of metabolic score with the lag time*

### Supplementary Table 6. Association of metabolic status categories with cognitive change, overall and by ethnicity

|  | **Total Cohort (n = 402)** | | | **Mexican American (n=209)** | | | **Non-Hispanic White (n=193)** | | |
| --- | --- | --- | --- | --- | --- | --- | --- | --- | --- |
|  | **β** | **SE** | **p-value** | **β** | **SE** | **p-value** | **β** | **SE** | **p-value** |
| ΔTime | -0.07 | 0.03 | 0.03 | -0.07 | 0.04 | 0.05 | -0.06 | 0.05 | 0.23 |
| MUN | 0.49 | 0.33 | 0.14 | 0.68 | 0.45 | 0.13 | -0.01 | 0.41 | 0.98 |
| MHO | -0.32 | 1.28 | 0.80 | 1.95 | 1.28 | 0.13 | -3.56 | 2.01 | 0.08 |
| MHN | 0.93 | 0.39 | 0.02 | 2.01 | 0.58 | < 0.01 | -0.20 | 0.45 | 0.66 |
| ΔTime:MUN | -0.07 | 0.04 | 0.06 | -0.09 | 0.06 | 0.10 | -0.05 | 0.06 | 0.37 |
| ΔTime:MHO | 0.10 | 0.10 | 0.30 | 0.02 | 0.10 | 0.84 | 0.21 | 0.13 | 0.10 |
| ΔTime:MHN | -0.06 | 0.05 | 0.25 | -0.16 | 0.08 | 0.06 | 0.03 | 0.06 | 0.61 |

**Models were adjusted for age, sex, income, education, ΔTime, and ethnicity (for total cohort only), with metabolically unhealthy obese as the reference*

*† Abbreviations: MUO – metabolically unhealthy obese (reference category); MUN – metabolically unhealthy normal; MHO – metabolically healthy obese; MHN – metabolically healthy normal; ΔTime - the lag time from cognitive assessment baseline to follow-up 1, 2, and 3; ΔTime:MUN – interaction effect of metabolic score with the lag time*

*‡ In the total cohort, 43 participants were metabolically healthy normal-weight (BMI < 30 kg/m^2^ and healthy metabolic status), 32 participants were metabolically healthy overweight/obese (BMI ≥ 30 kg/m^2^ and healthy metabolic status), 84 participants were metabolically unhealthy normal-weight (BMI < 30 kg/m^2^ and healthy metabolic status), and 243 participants were metabolically unhealthy overweight/obese (BMI ≥ 30 kg/m^2^ and unhealthy metabolic status).*

### Supplementary Table 7. Association of fasting blood glucose LS7, BMI LS7, and physical activity LS7 scores with cognitive change, stratified by ethnicity and neighborhood for previously reported significant interaction results (p < 0.05)

|  | **Mexican American** | | | | | | | | | | **Non-Hispanic White** | | | | | | |
| --- | --- | --- | --- | --- | --- | --- | --- | --- | --- | --- | --- | --- | --- | --- | --- | --- | --- |
|  | | **Barrio (n = 107)** | | | **Transitional (n= 45)** | | | **Suburban (n = 57)** | | | | **Transitional (n = 100)** | | | **Suburban (n = 93)** | | |
|  | | **β** | **SE** | **p-value** | **β** | **SE** | **p-value** | **β** | **SE** | **p-value** | | **β** | **SE** | **p-value** | **β** | **SE** | **p-value** |
| **Blood Glucose** | |  |  |  |  |  |  |  |  |  | |  |  |  |  |  |  |
| ΔTime | | - | - | - | - | - | - | - | - | - | | -0.24 | 0.12 | 0.04 | -0.35 | 0.02 | < 0.01 |
| Intermediate+Ideal | | - | - | - | - | - | - | - | - | - | | -0.67 | 1.20 | 0.58 | -1.11 | 0.23 | < 0.01 |
| Intermediate+Ideal: ΔTime | | - | - | - | - | - | - | **-** | **-** | **-** | | 0.18 | 0.12 | 0.13 | **0.25** | **0.03** | **< 0.01** |
| **BMI LS7** | |  |  |  |  |  |  |  |  |  | |  |  |  |  |  |  |
| ΔTime | | -0.09 | 0.04 | 0.03 | -0.09 | 0.09 | 0.31 | 0.09 | 0.08 | 0.24 | | - | - | - | - | - | - |
| Intermediate+Ideal | | 0.98 | 0.66 | 0.14 | 0.88 | 0.81 | 0.28 | 0.71 | 0.65 | 0.28 | | - | - | - | - | - | - |
| Intermediate+Ideal: ΔTime | | **-0.17** | **0.08** | **0.04** | -0.05 | 0.11 | 0.65 | **-0.20** | **0.09** | **0.02** | | - | - | - | - | - | - |
| **Physical Activity** | |  |  |  |  |  |  |  |  |  | |  |  |  |  |  |  |
| ΔTime | | -0.21 | 0.05 | < 0.01 | -0.12 | 0.07 | 0.06 | -0.12 | 0.05 | 0.03 | | - | - | - | - | - | - |
| Intermediate+Ideal | | -0.95 | 0.82 | 0.25 | -0.80 | 0.81 | 0.32 | 0.07 | 0.53 | 0.90 | | - | - | - | - | - | - |
| Intermediate+Ideal: ΔTime | | **0.21** | **0.09** | **0.02** | 0.01 | 0.09 | 0.90 | 0.07 | 0.07 | 0.35 | | - | - | - | - | - | - |
| *Models adjusted for age, sex, income, education, and ΔTime  † Abbreviations: LS7–Life's Simple 7 cardiovascular health index; ΔTime - the lag time from cognitive assessment baseline to follow-up 1, 2, and 3; Intermediate+Ideal - combination of LS7 intermediate and ideal category; Intermediate+Ideal:ΔT – interaction effect of LS7 component with the lag time  ‡ Blanks in table indicate no significance in beta coefficient in primary analysis for corresponding LS7 component | | | | | | | | | | | | | | | | | |

### Supplementary Figure 1. Violin plot showing the distribution of LS7 total score across ethnicity and neighborhoods.

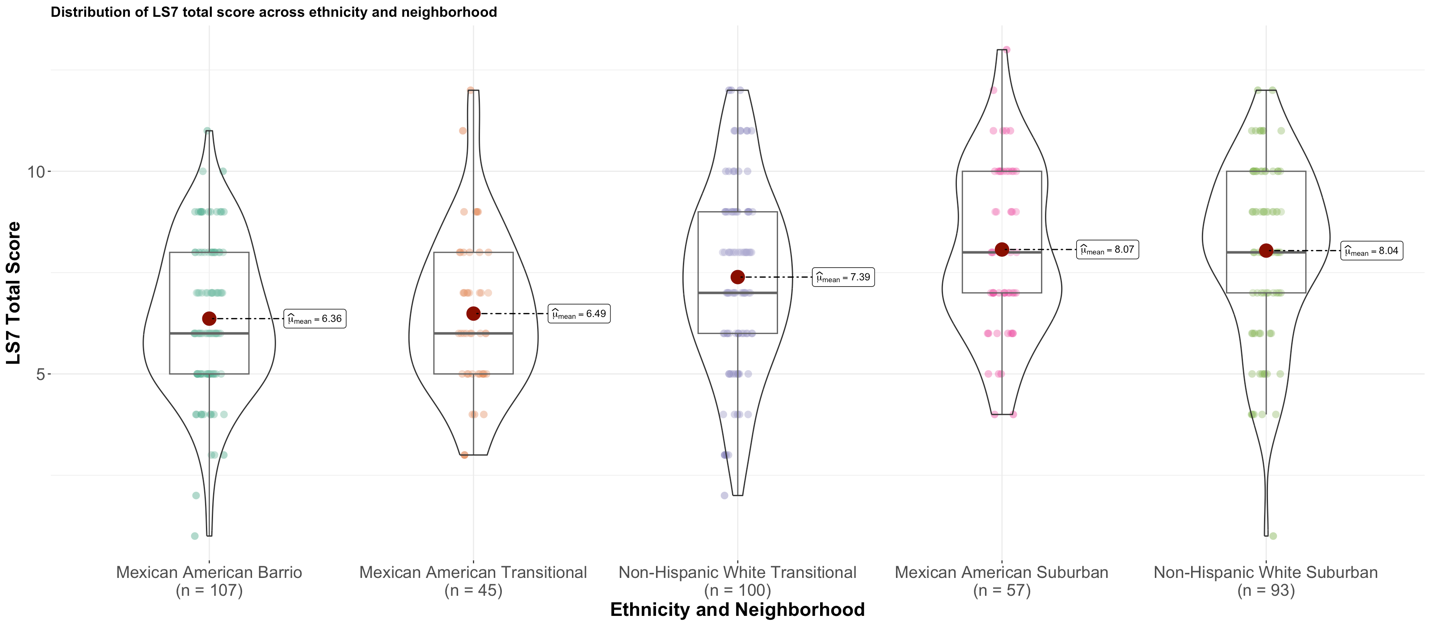

### Supplementary Figure 2. Violin plot showing the distribution of BMI across ethnicity and neighborhoods.

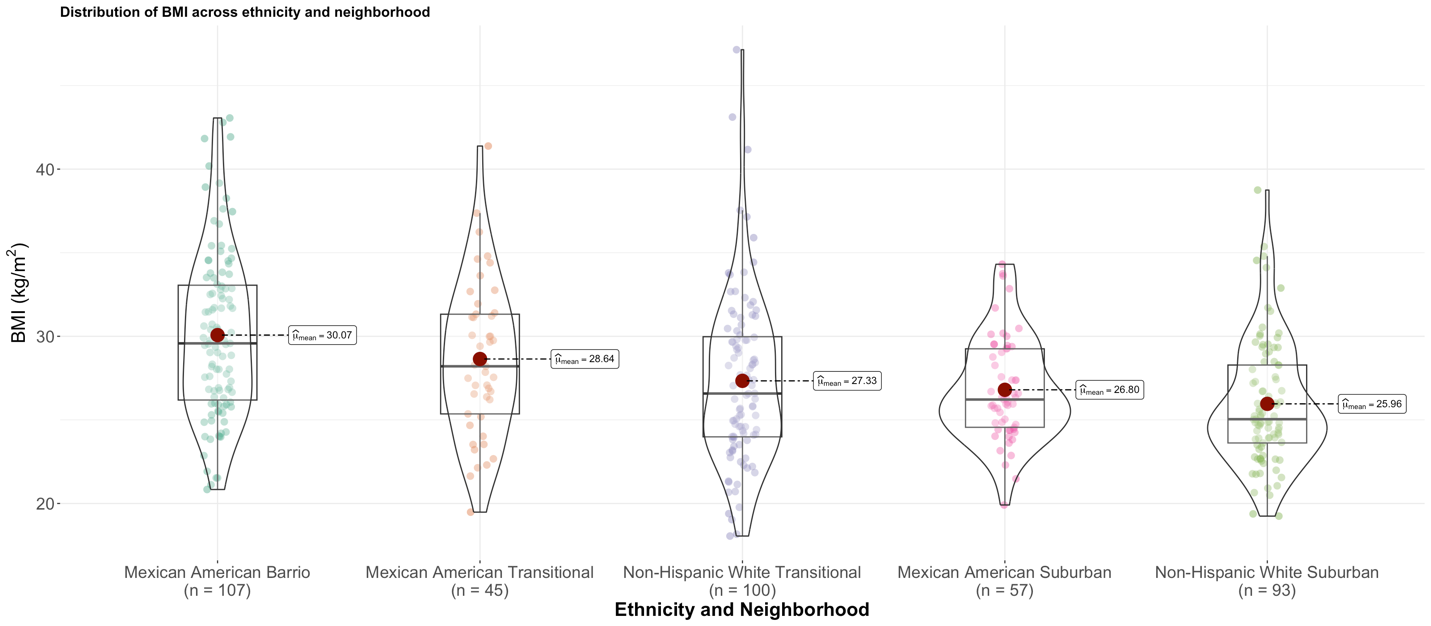

### Supplementary Figure 3. Violin plot showing the distribution of fasting blood glucose across ethnicity and neighborhood.

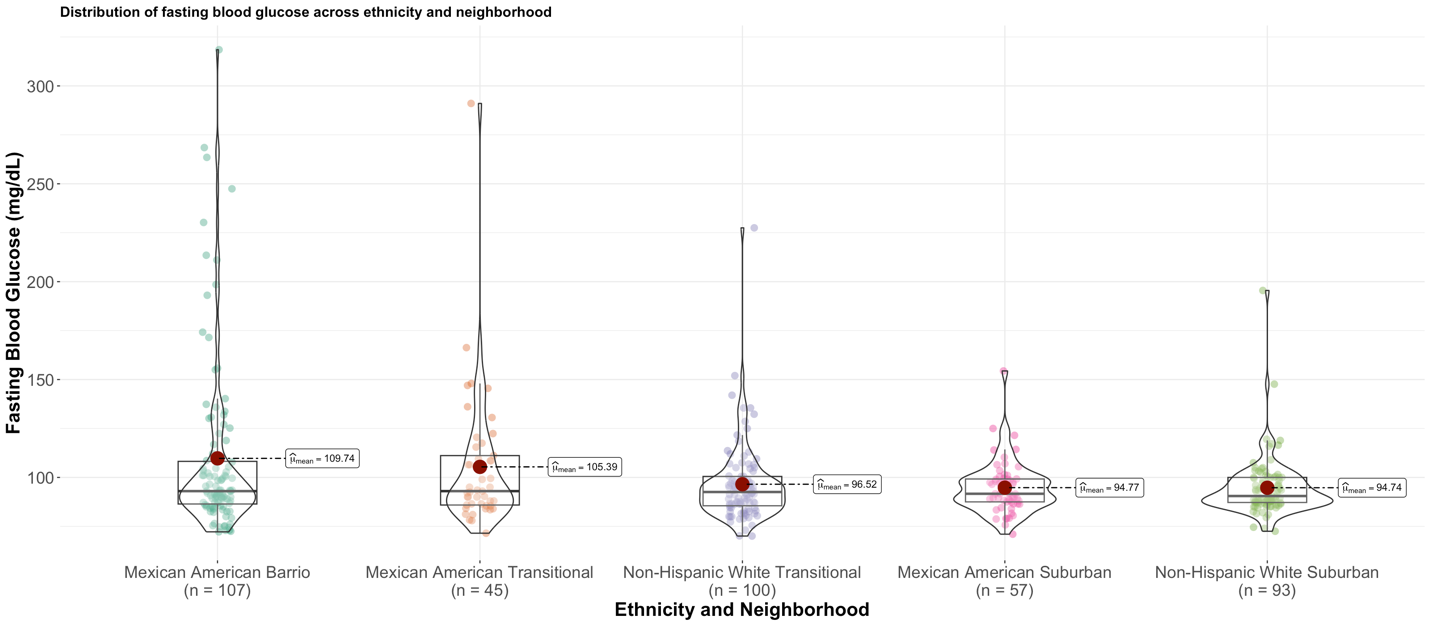

### Supplementary Figure 4. Violin plot showing the distribution of total cholesterol across ethnicity and neighborhood.

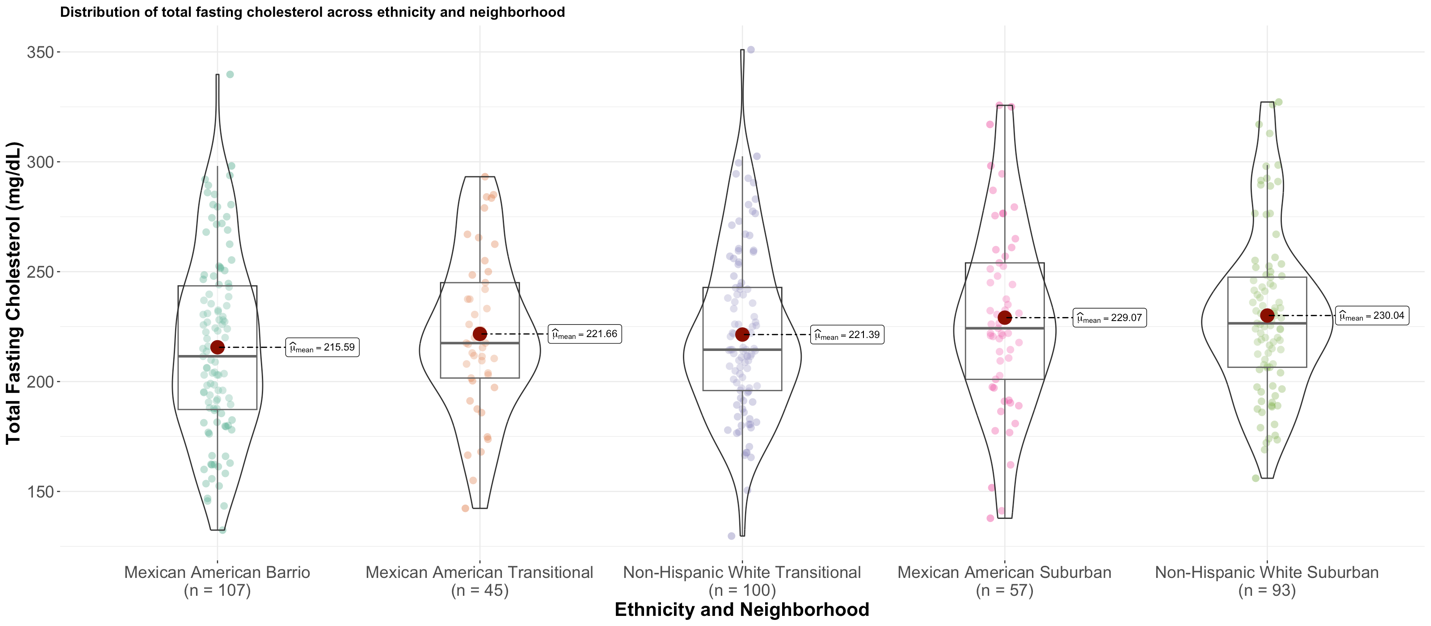

### Supplementary Figure 5. Violin plot showing the distribution of systolic blood pressure across ethnicity and neighborhood.

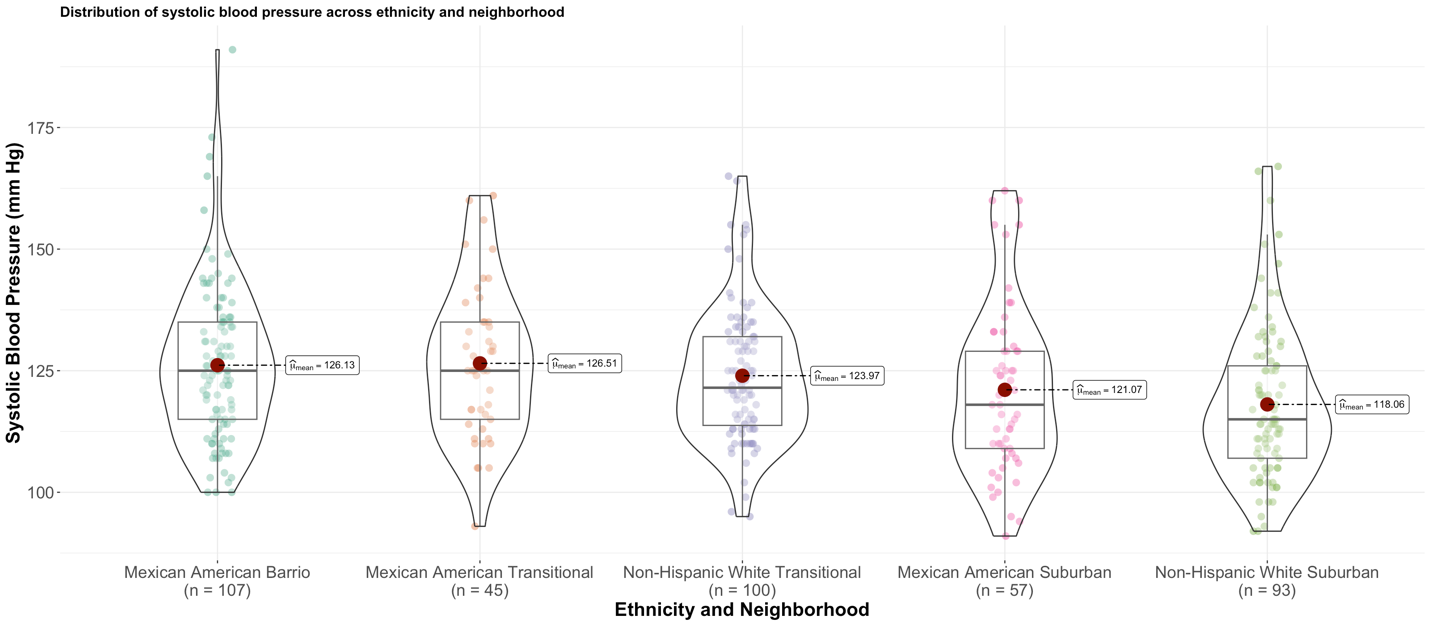

### Supplementary Figure 6. Violin plot showing the distribution of diastolic blood pressure across ethnicity and neighborhood.

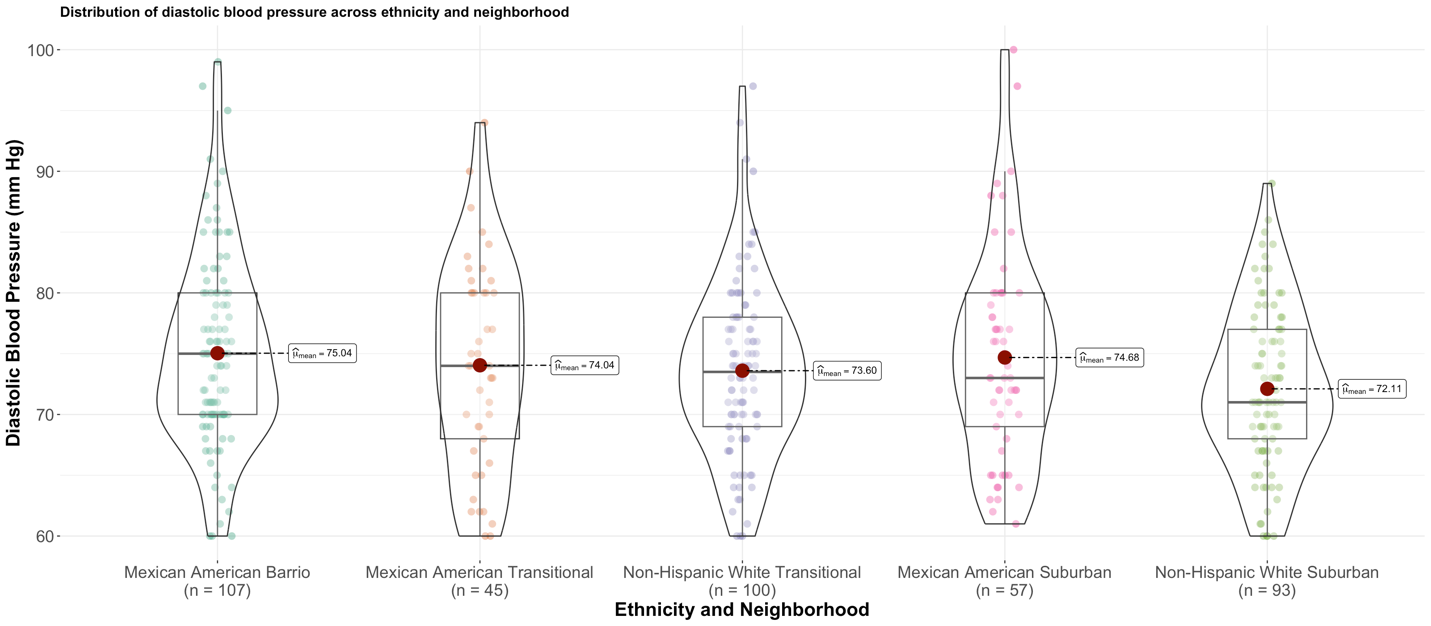

### Supplementary Figure 7. Violin plot showing the distribution of physical activity (in minutes/week) across ethnicity and neighborhood.

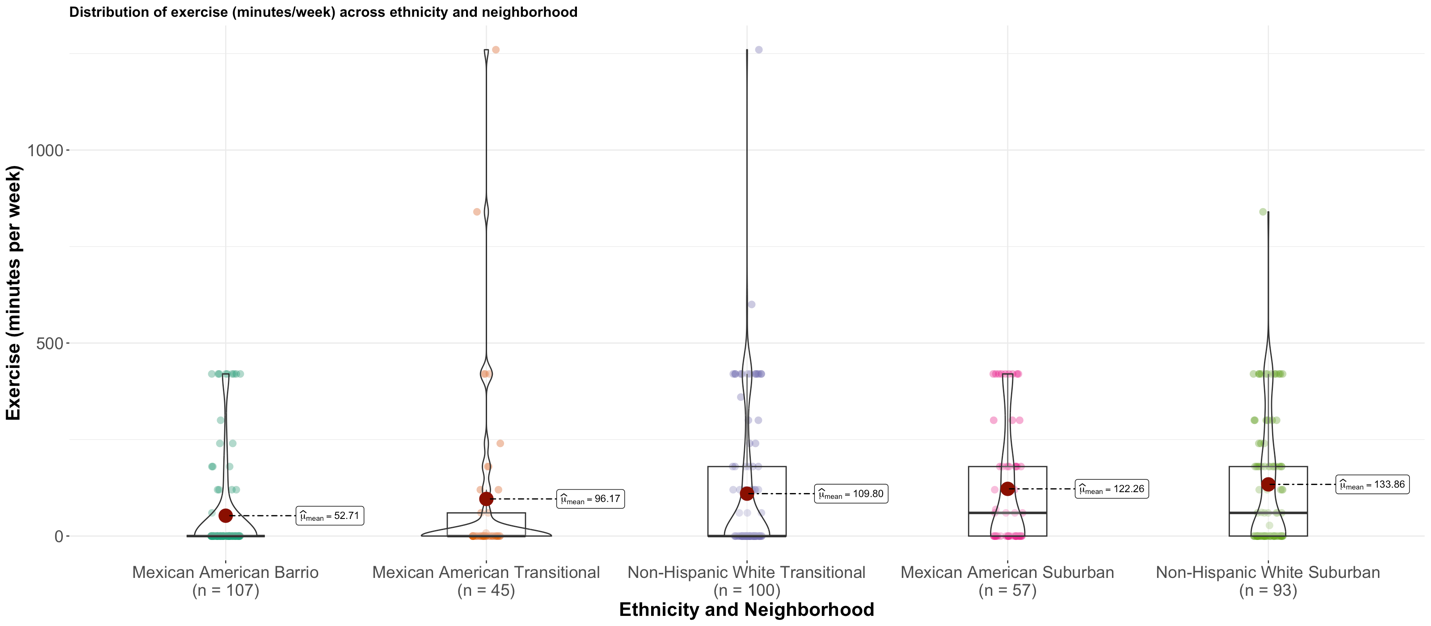

### Supplementary Figure 8. Violin plot showing the distribution of diet score (as calculated by the AHA Life’s Simple 7) across ethnicity and neighborhood.

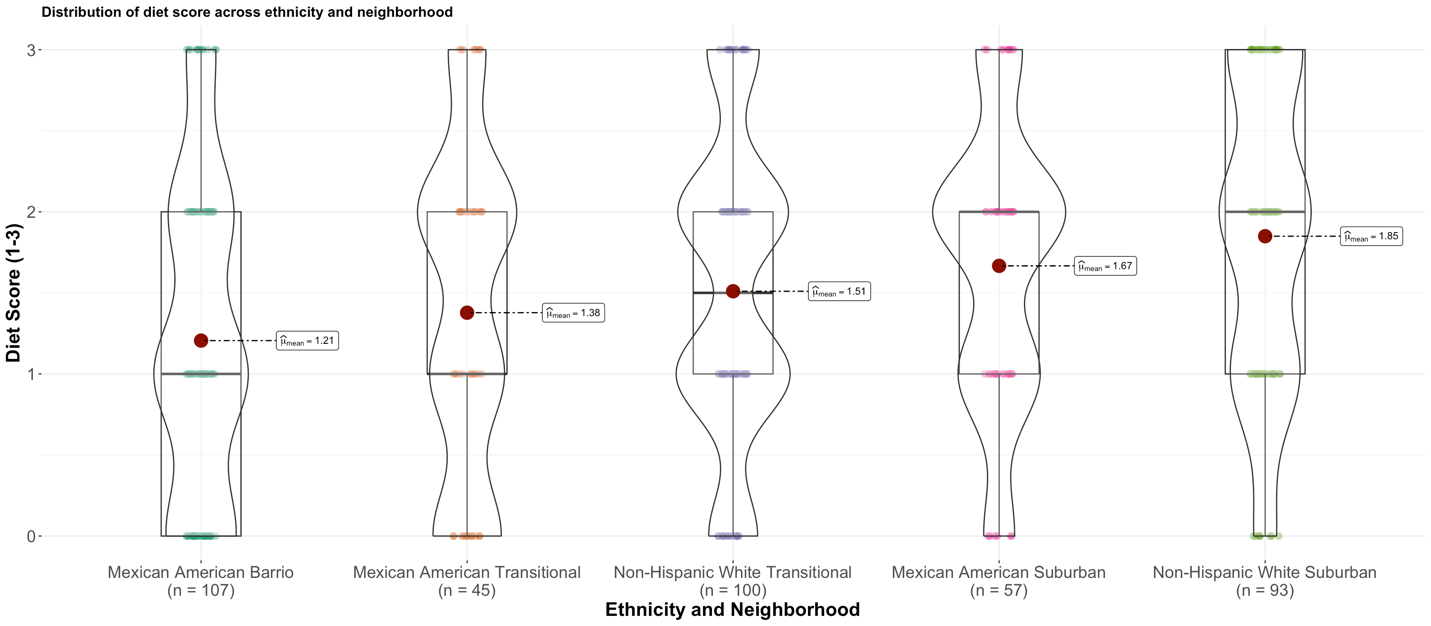

### Supplementary Figure 9. Bar plot showing the distribution of LS7 tertile categories across ethnicity and neighborhood.

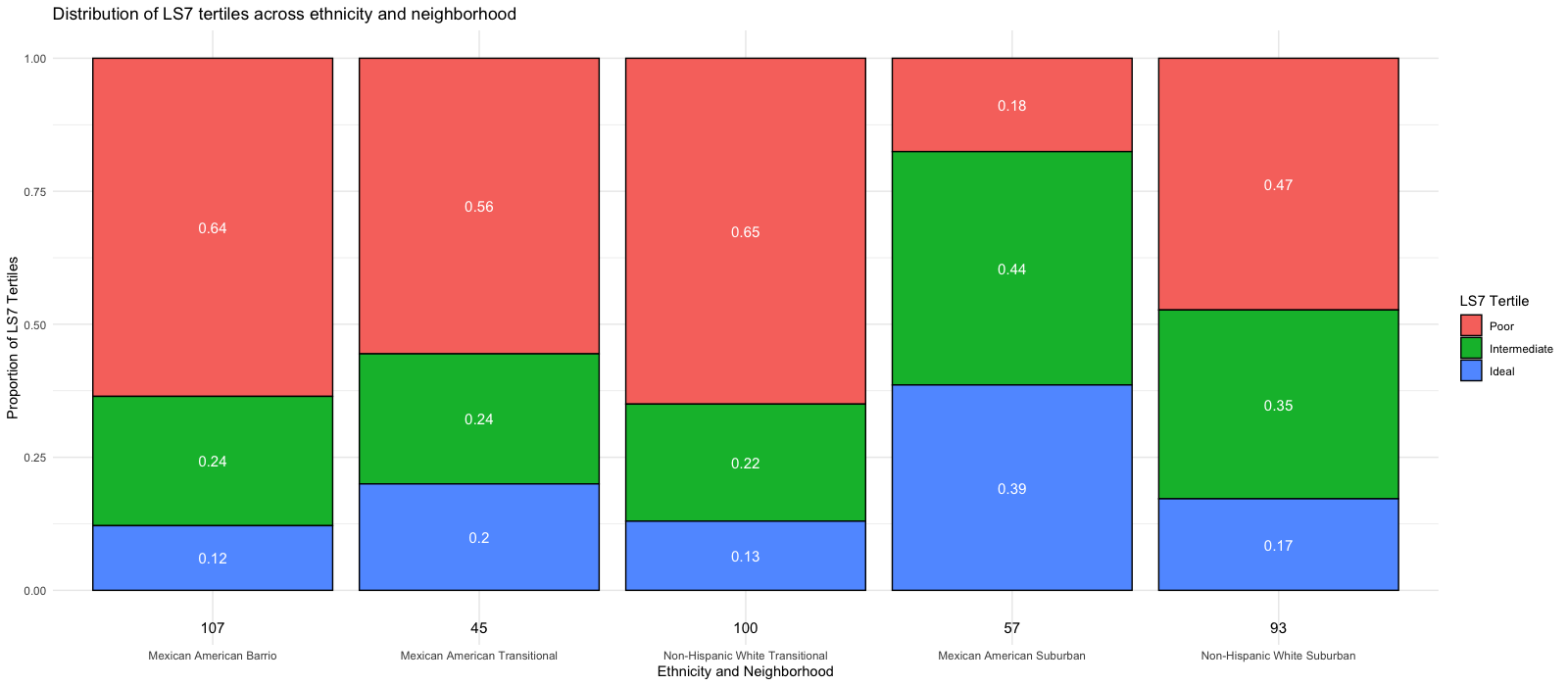

### Supplementary Figure 10. Bar plot showing the distribution of Smoking LS7 categories across ethnicity and neighborhood.

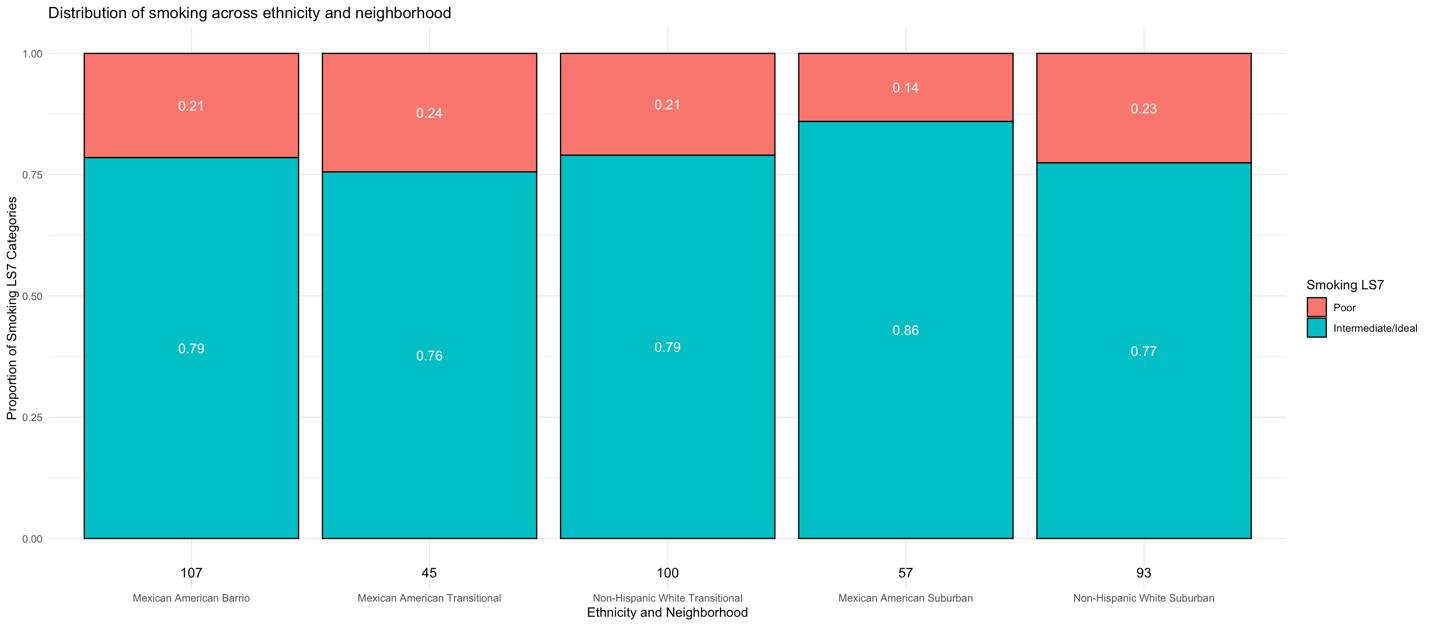

Supplementary Figure 11. Figure shows the decrease in cognitive state predicted by the interaction of LS7 score with cognitive decline in the (a) total cohort, (b) Mexican American cohort, and (c) non-Hispanic White Cohort. Predicted outcomes and confidence interval bands are shown, with red showing the lowest LS7 scores, blue the intermediate LS7 scores, and green the highest LS7 scores.

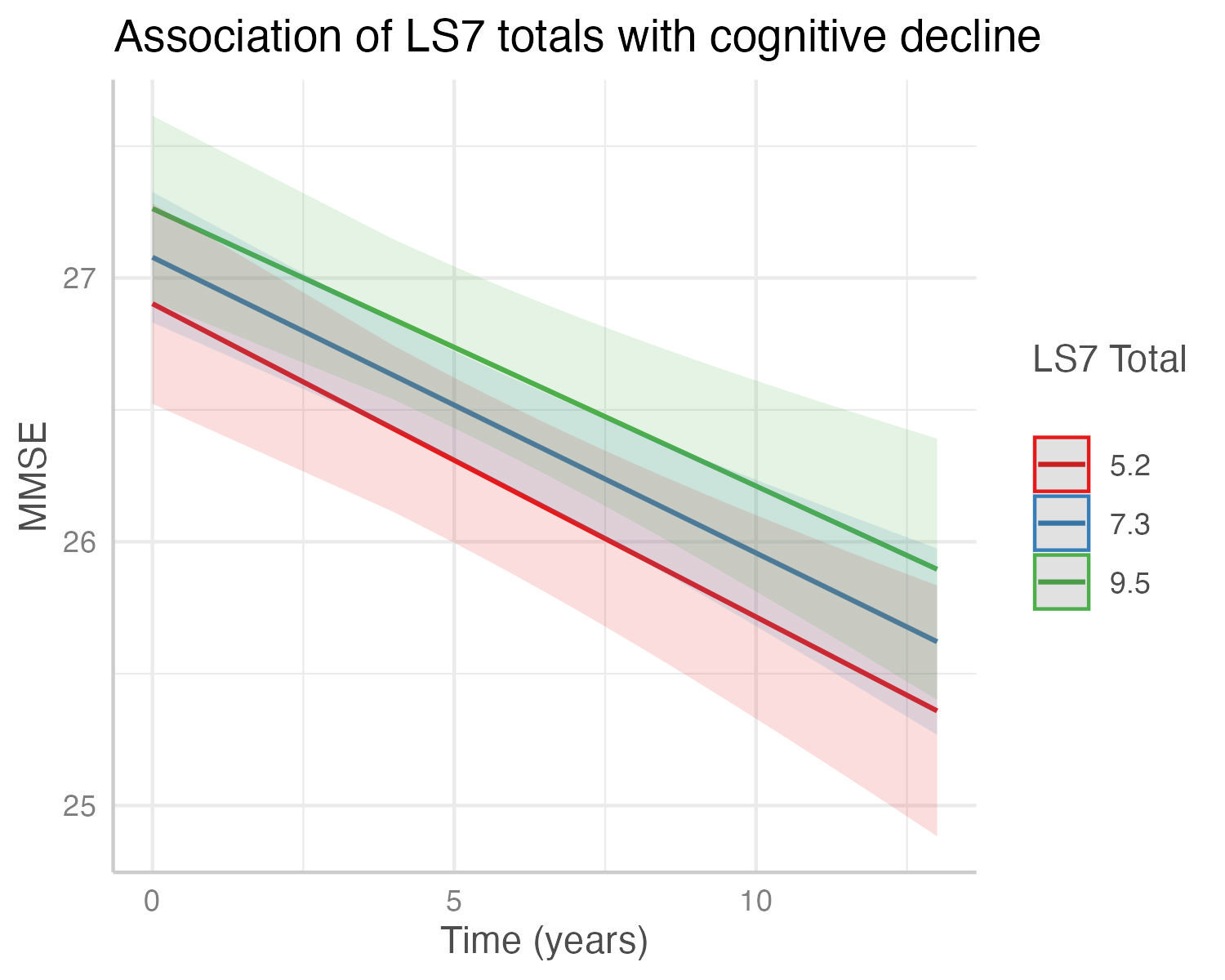

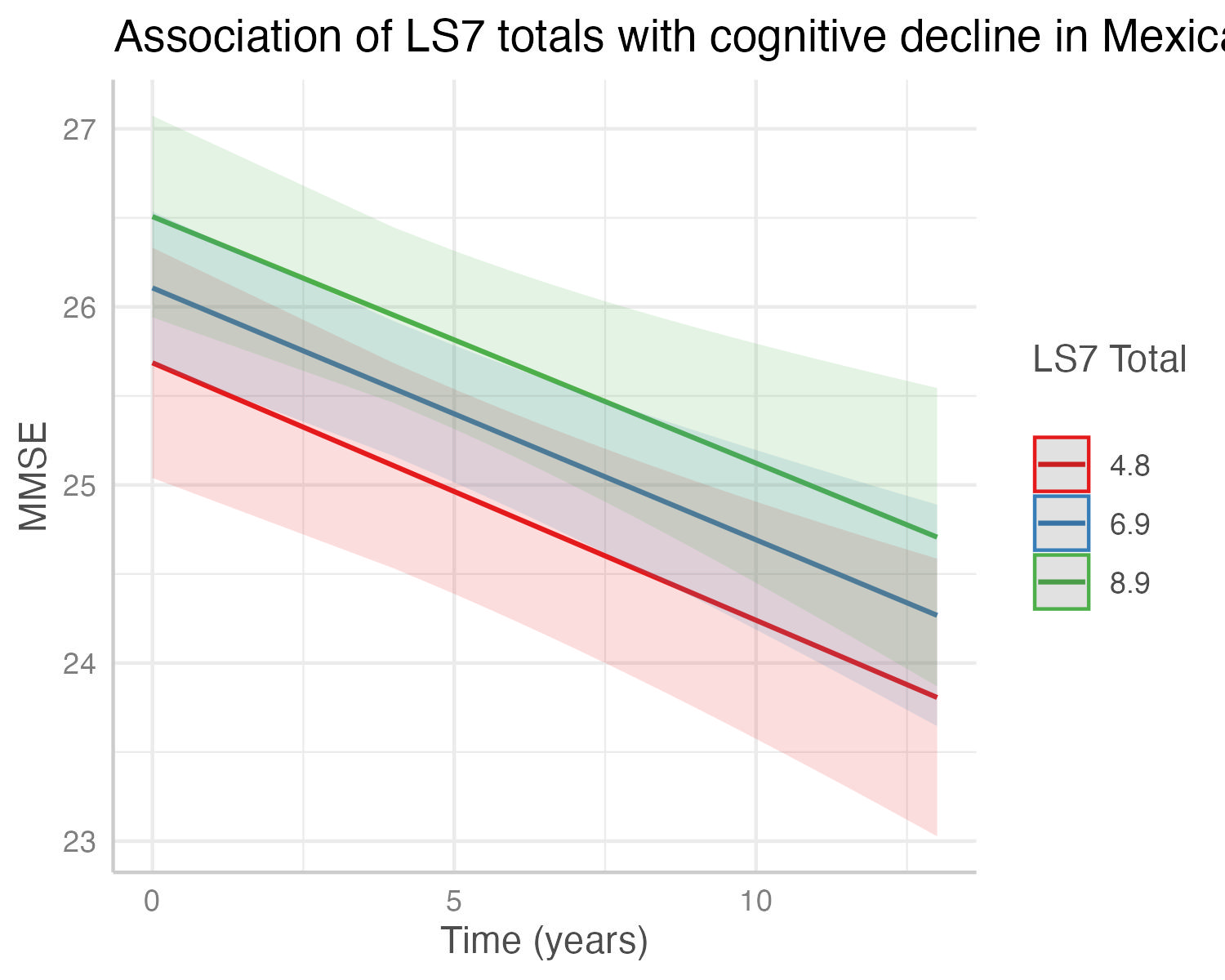
.
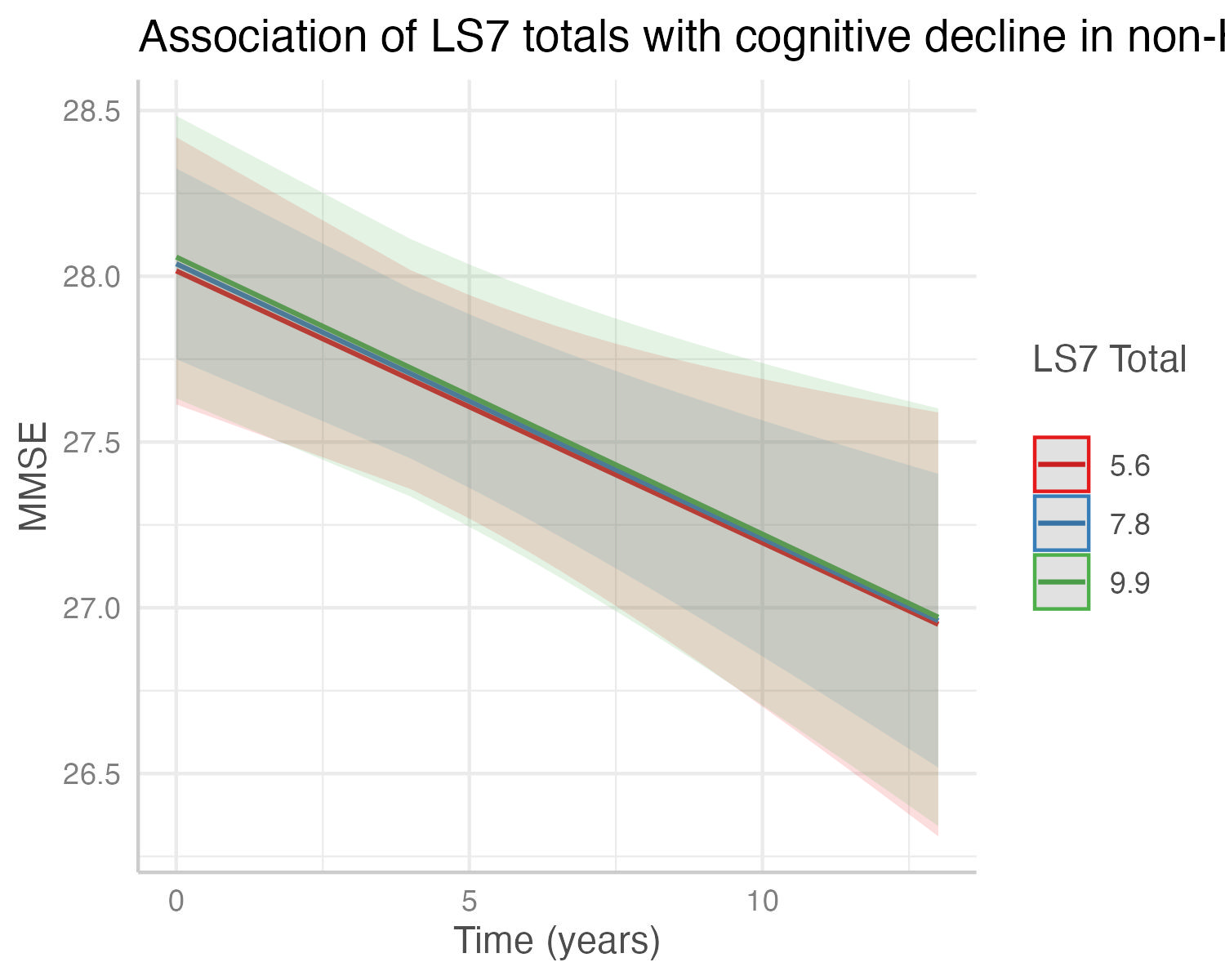

(a) Total Cohort	(b) Mexican American Cohort	(c) non-Hispanic White Cohort

### Supplementary Figure 12. Figure shows the decrease in cognitive state predicted by the interaction of LS7 tertile categories with cognitive decline in the (a) total cohort, (b) Mexican American cohort, and (c) non-Hispanic White Cohort. Predicted outcomes and confidence interval bands are shown, with red showing lowest LS7 tertile, blue the middle LS7 tertile, and green the highest LS7 tertile.

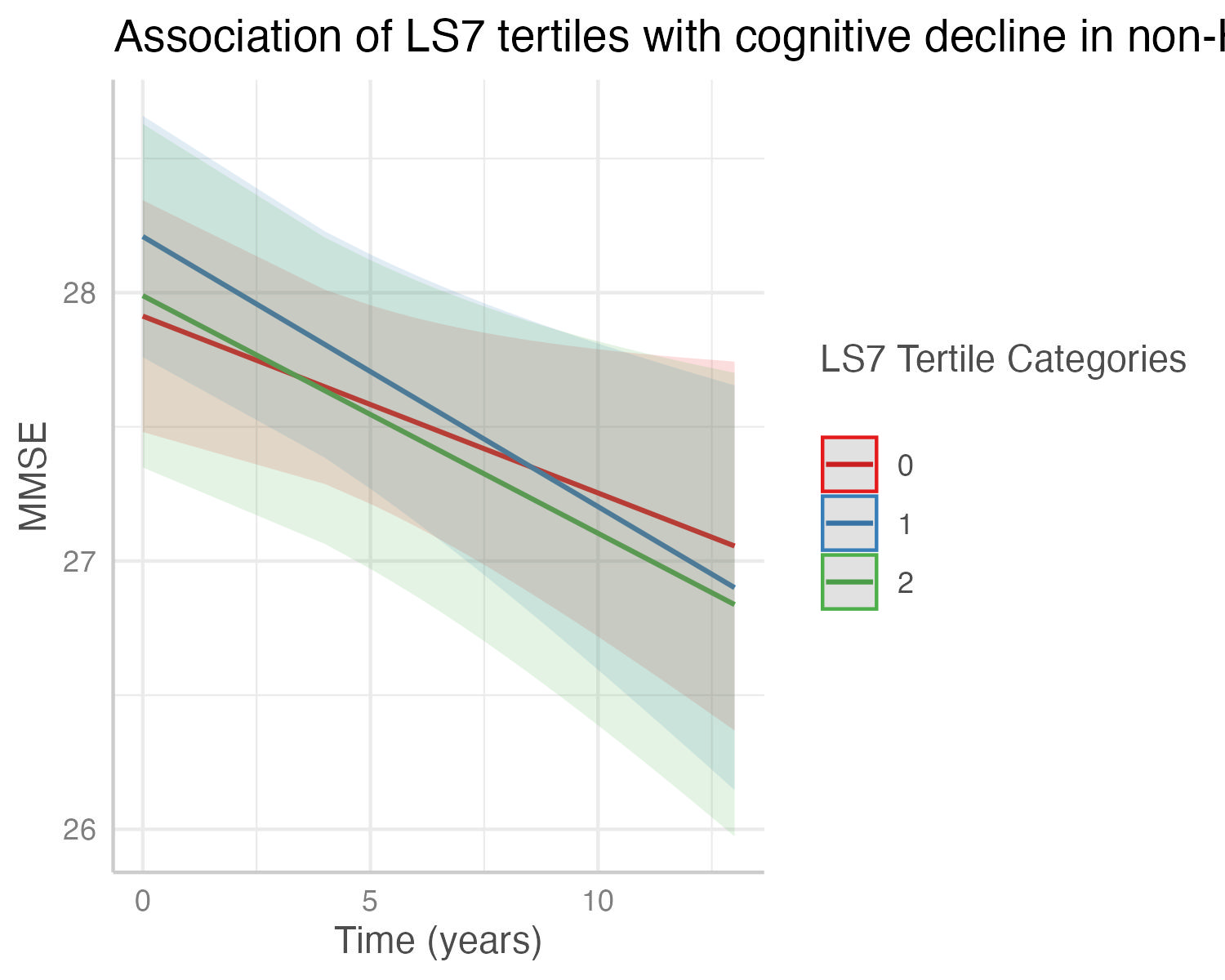

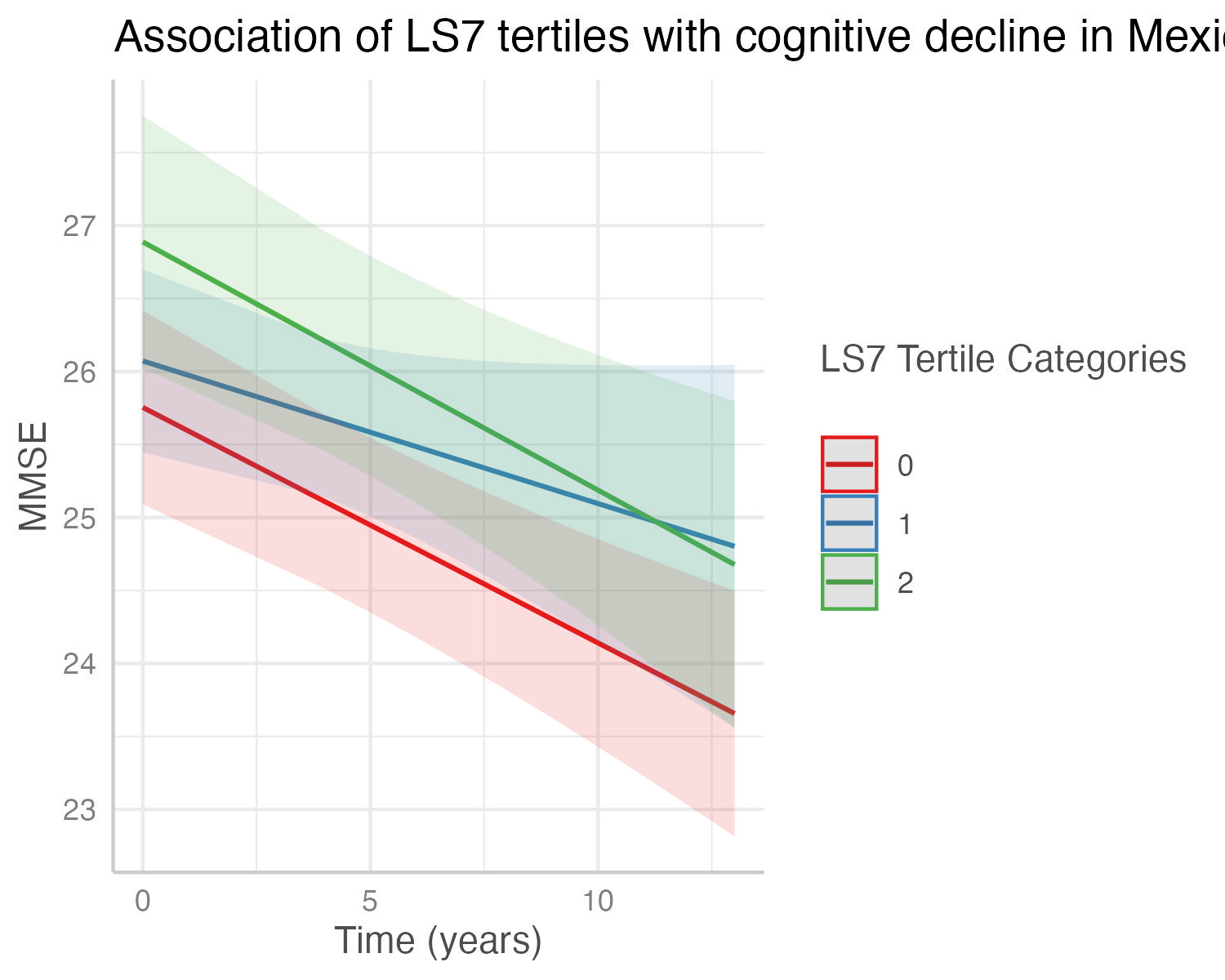

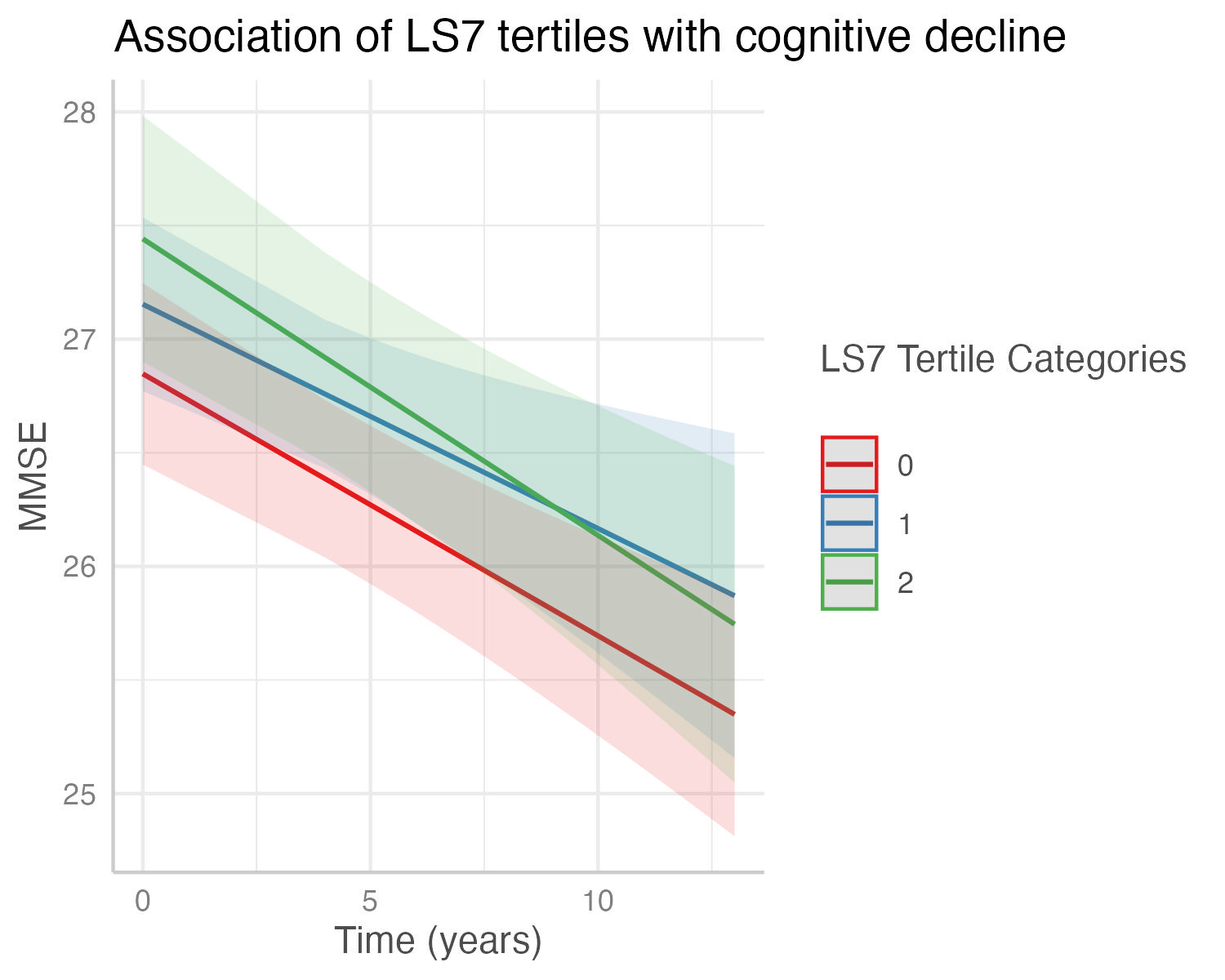

(a) Total Cohort	(b) Mexican American Cohort	(c) non-Hispanic White Cohort

### Supplementary Figure 13. Figure shows the decrease in cognitive state predicted by the interaction of total cholesterol LS7 with cognitive decline in the (a) total cohort, (b) Mexican American cohort, and (c) non-Hispanic White Cohort. Predicted outcomes and confidence interval bands are shown, with red showing the poor LS7 category and green the intermediate and ideal LS7 category.

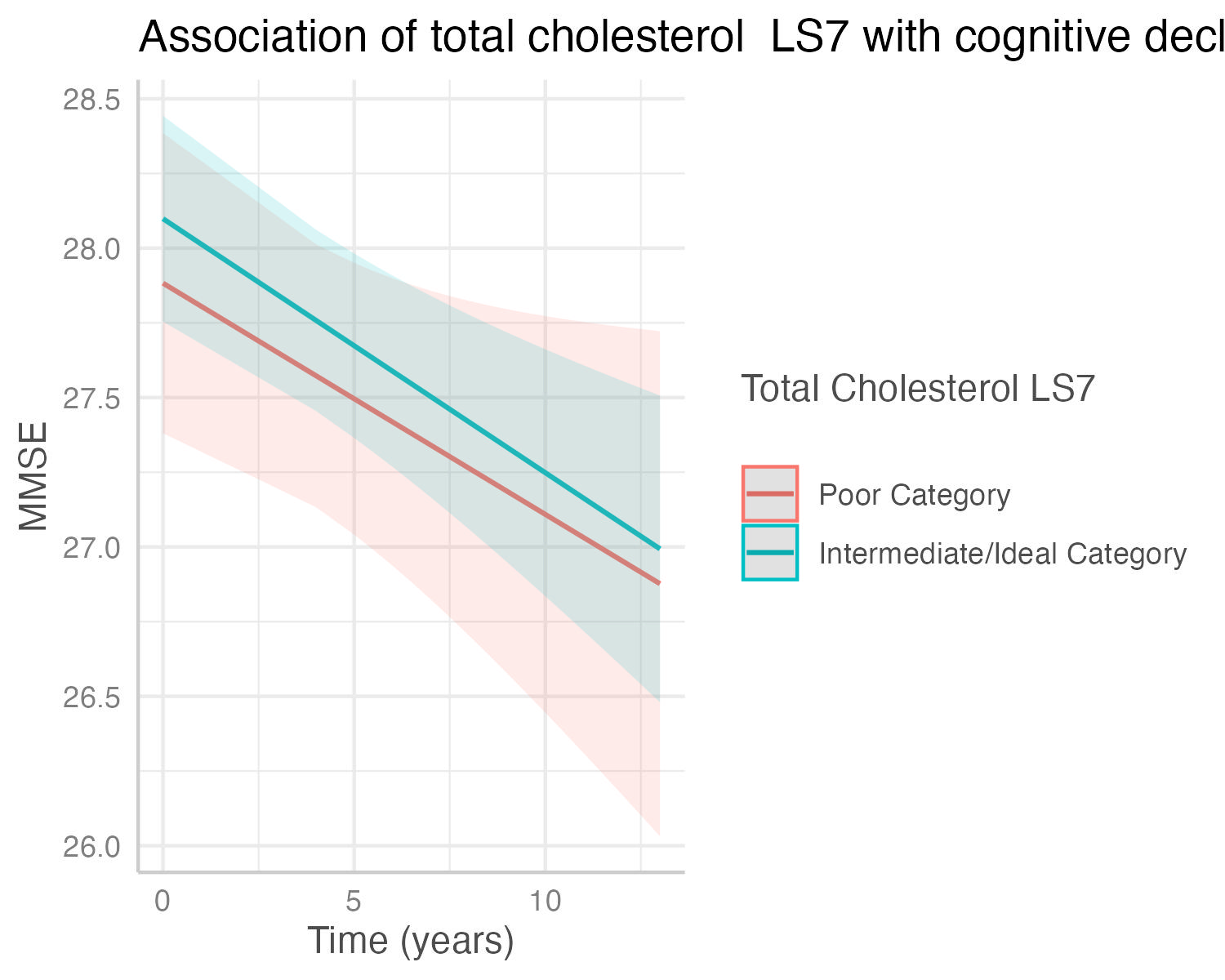

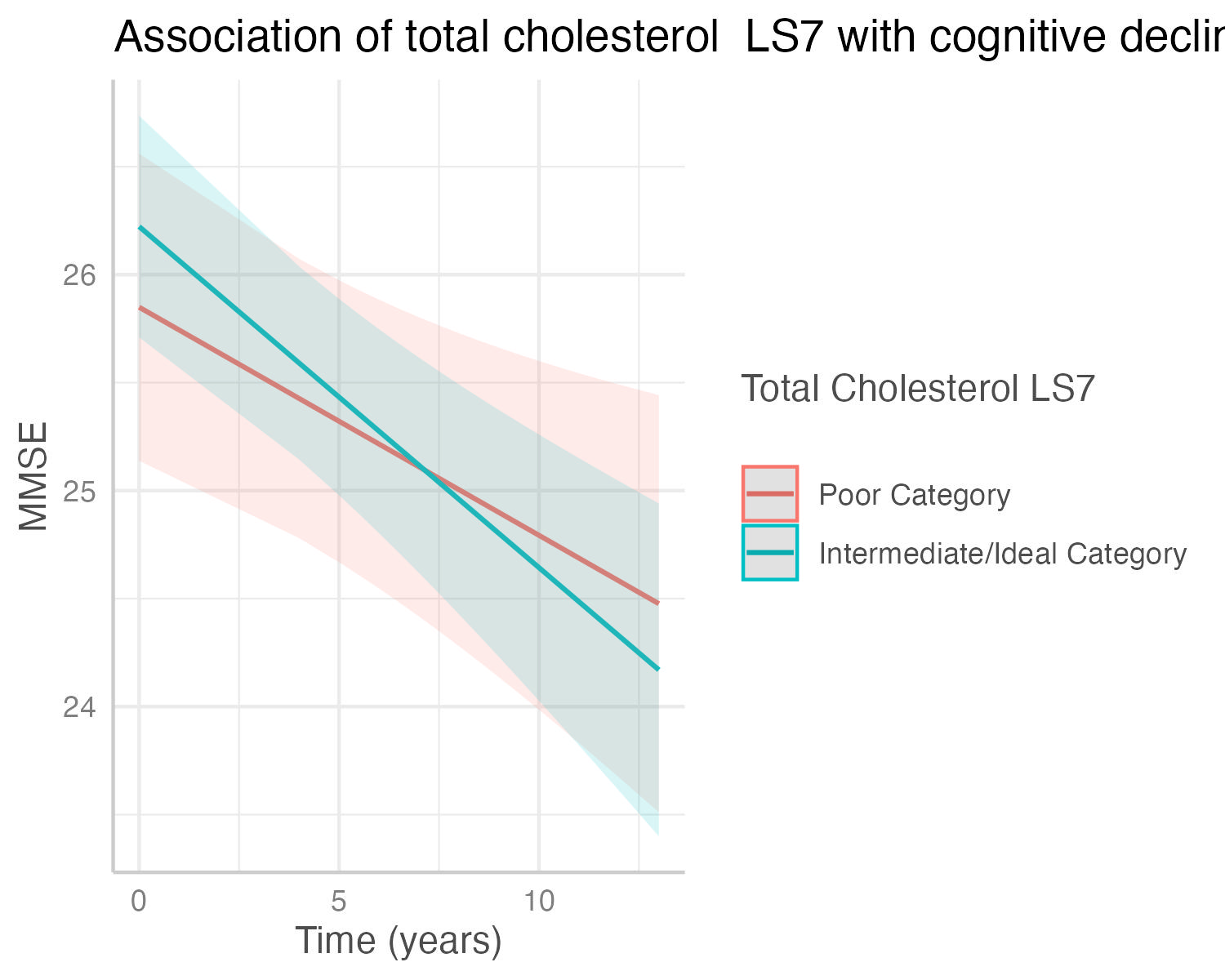

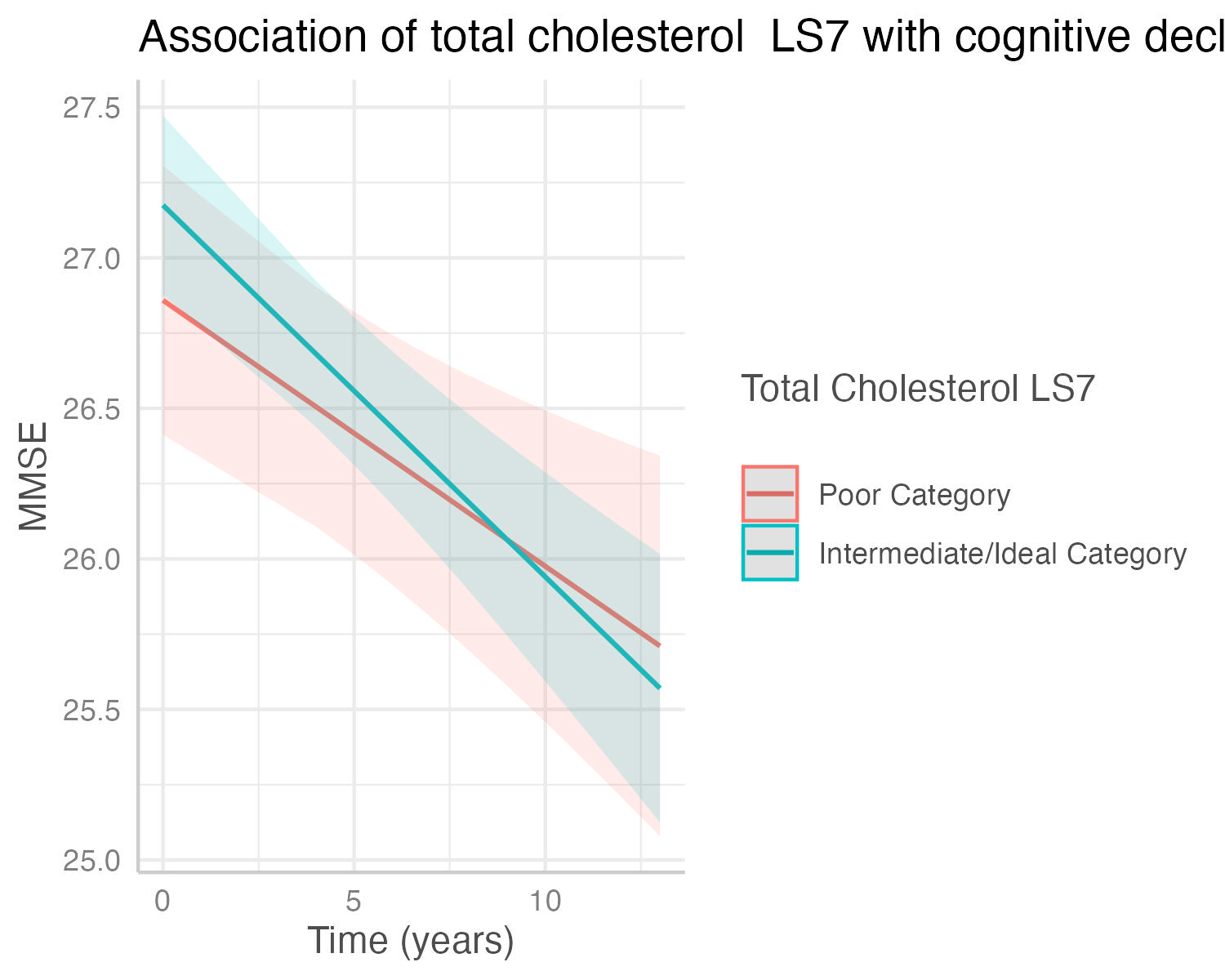

(a) Total Cohort	(b) Mexican American Cohort	(c) non-Hispanic White Cohort

### Supplementary Figure 14. Figure shows the decrease in cognitive state predicted by the interaction of blood pressure LS7 with cognitive decline in the (a) total cohort, (b) Mexican American cohort, and (c) non-Hispanic White Cohort. Predicted outcomes and confidence interval bands are shown, with red showing the poor LS7 category and green the intermediate and ideal LS7 category.

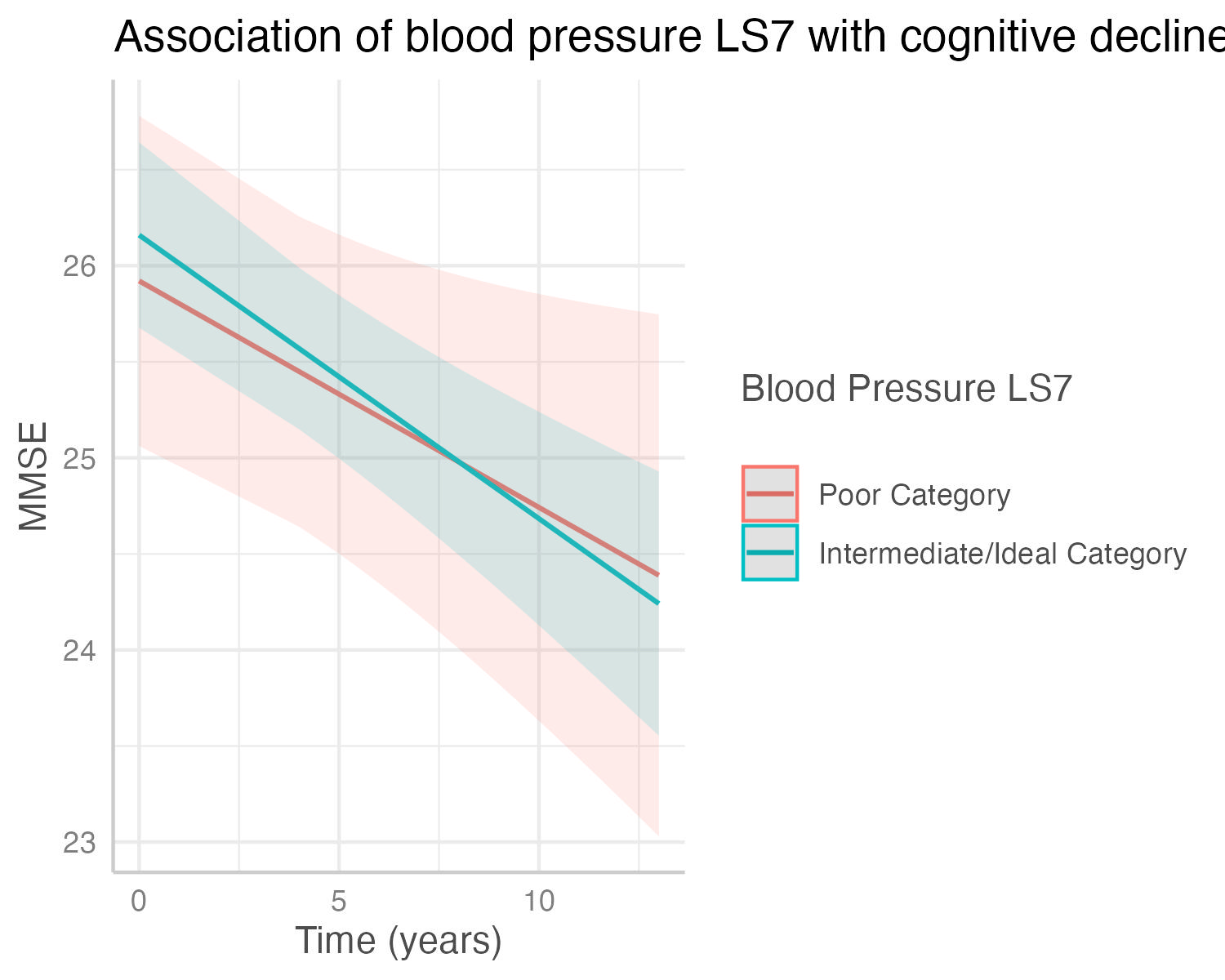

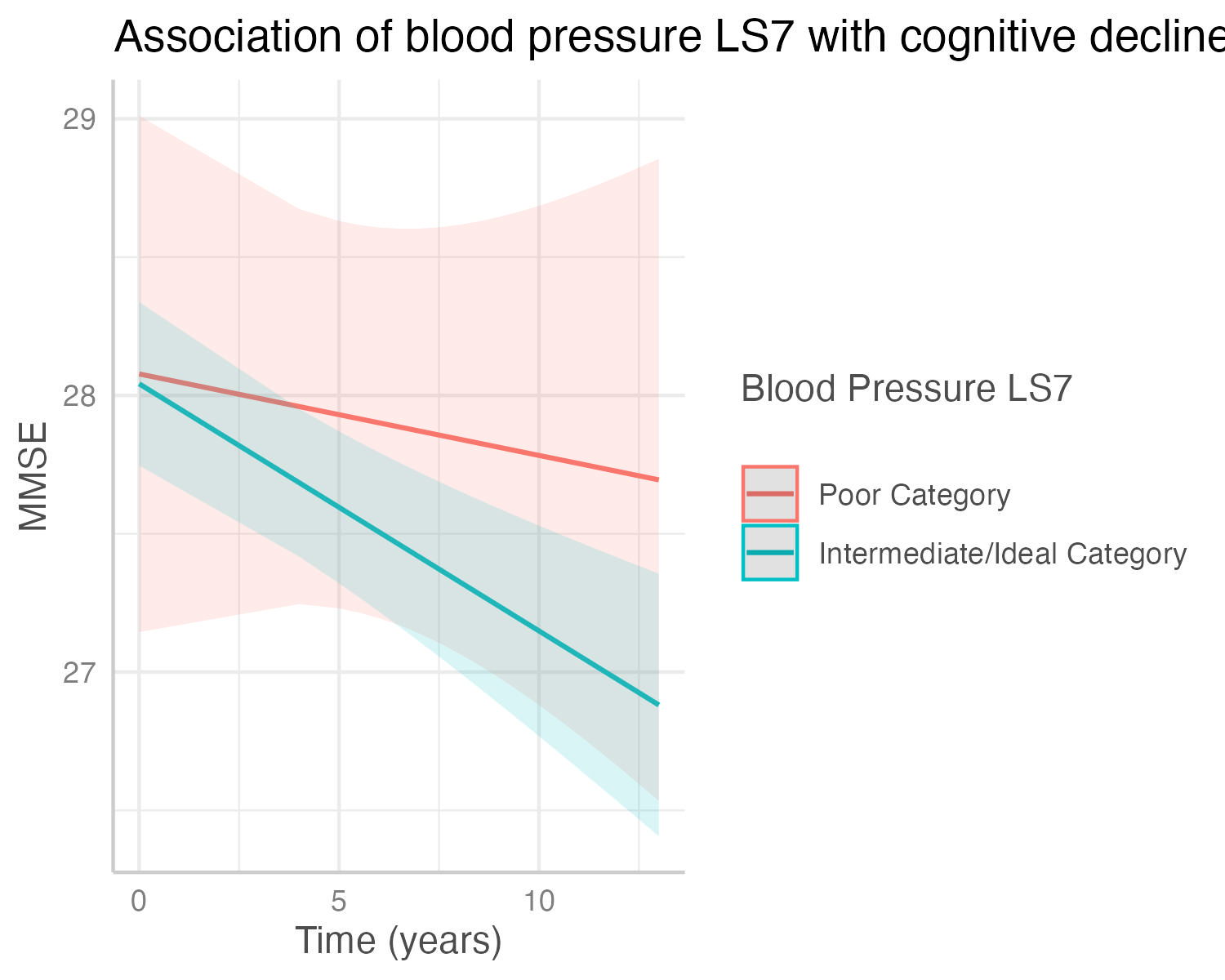

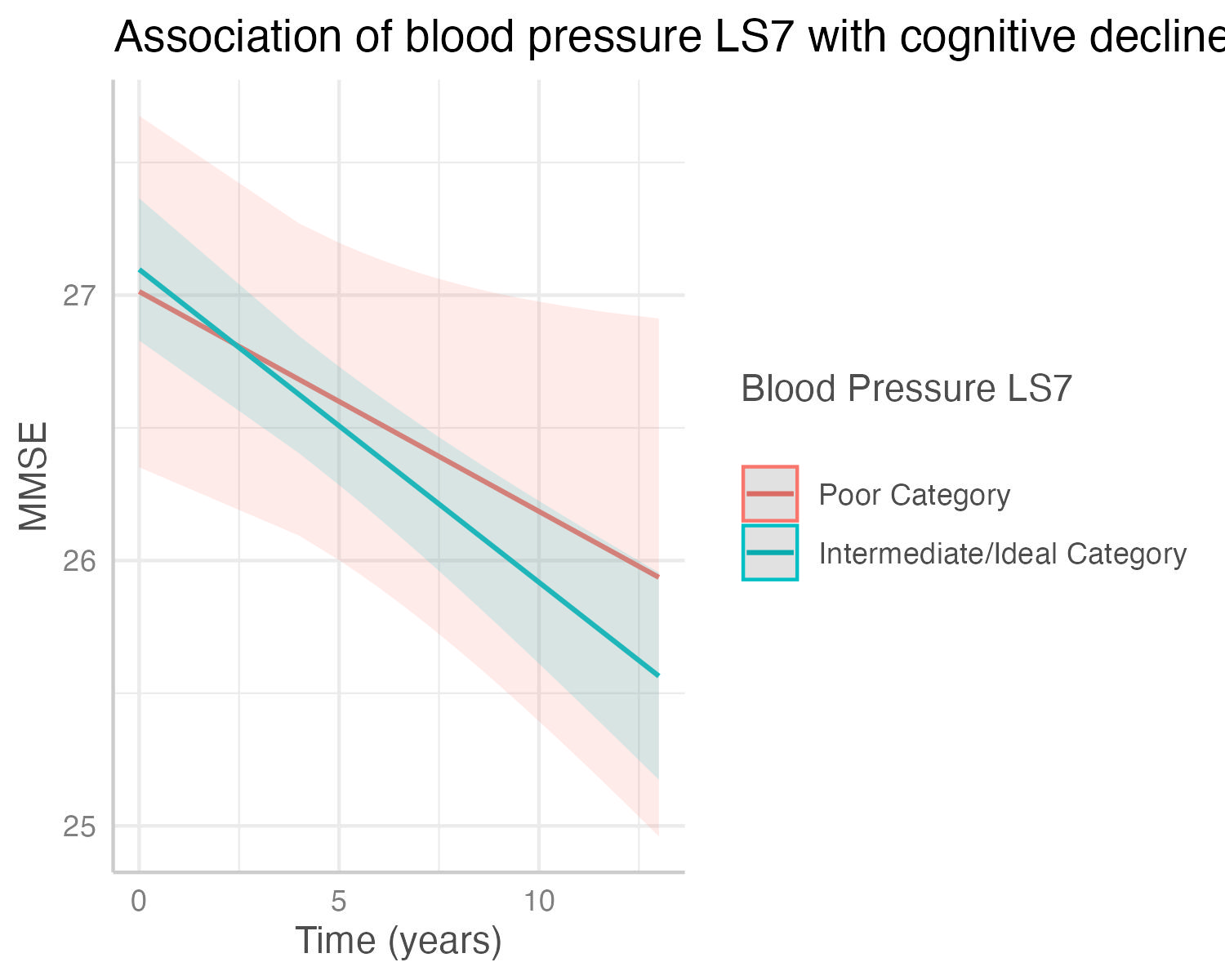

(a) Total Cohort	(b) Mexican American Cohort	(c) non-Hispanic White Cohort

### Supplementary Figure 15. Figure shows the decrease in cognitive state predicted by the interaction of smoking LS7 with cognitive decline in the (a) total cohort, (b) Mexican American cohort, and (c) non-Hispanic White Cohort. Predicted outcomes and confidence interval bands are shown, with red showing the poor LS7 category and green the intermediate and ideal LS7 category.

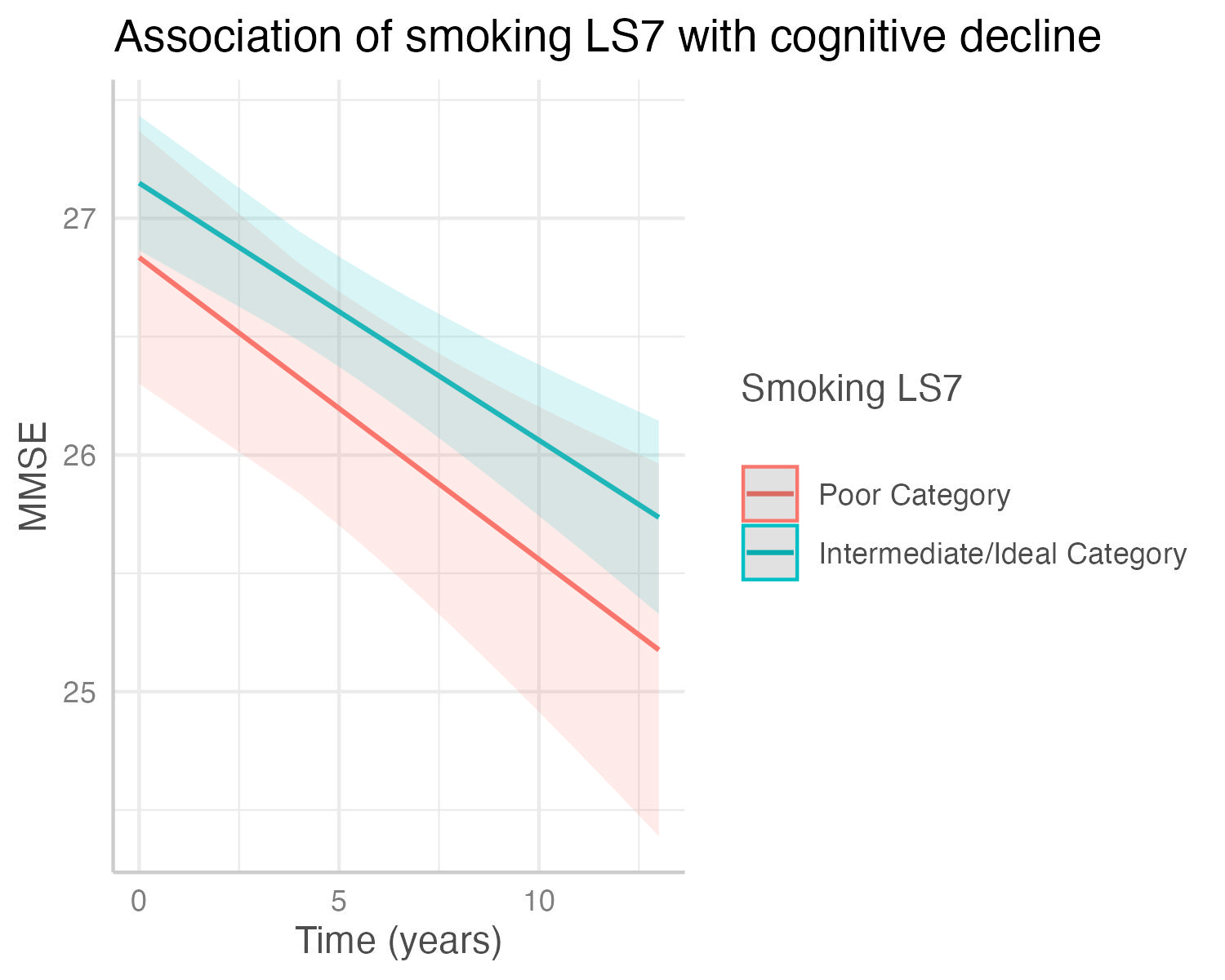

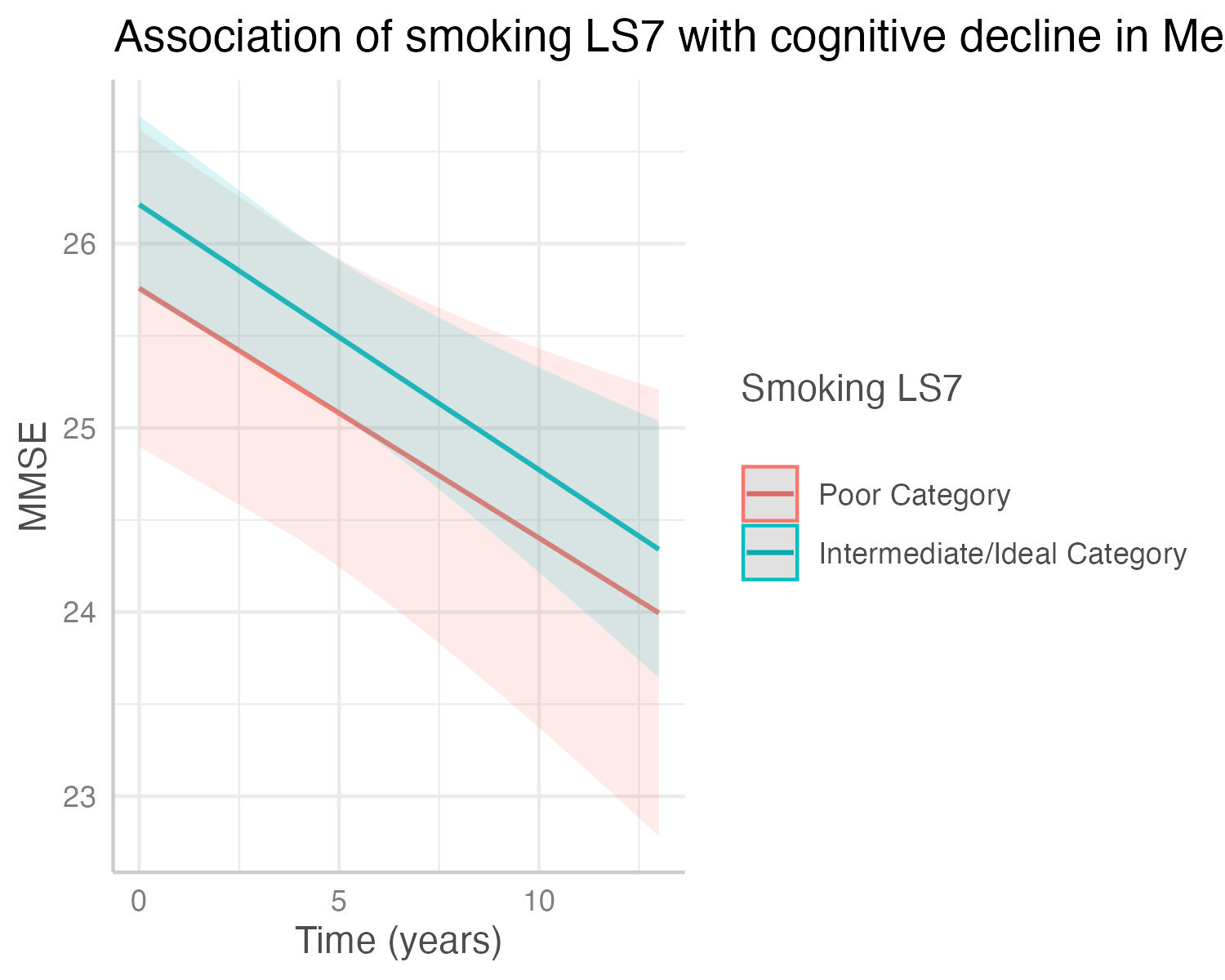

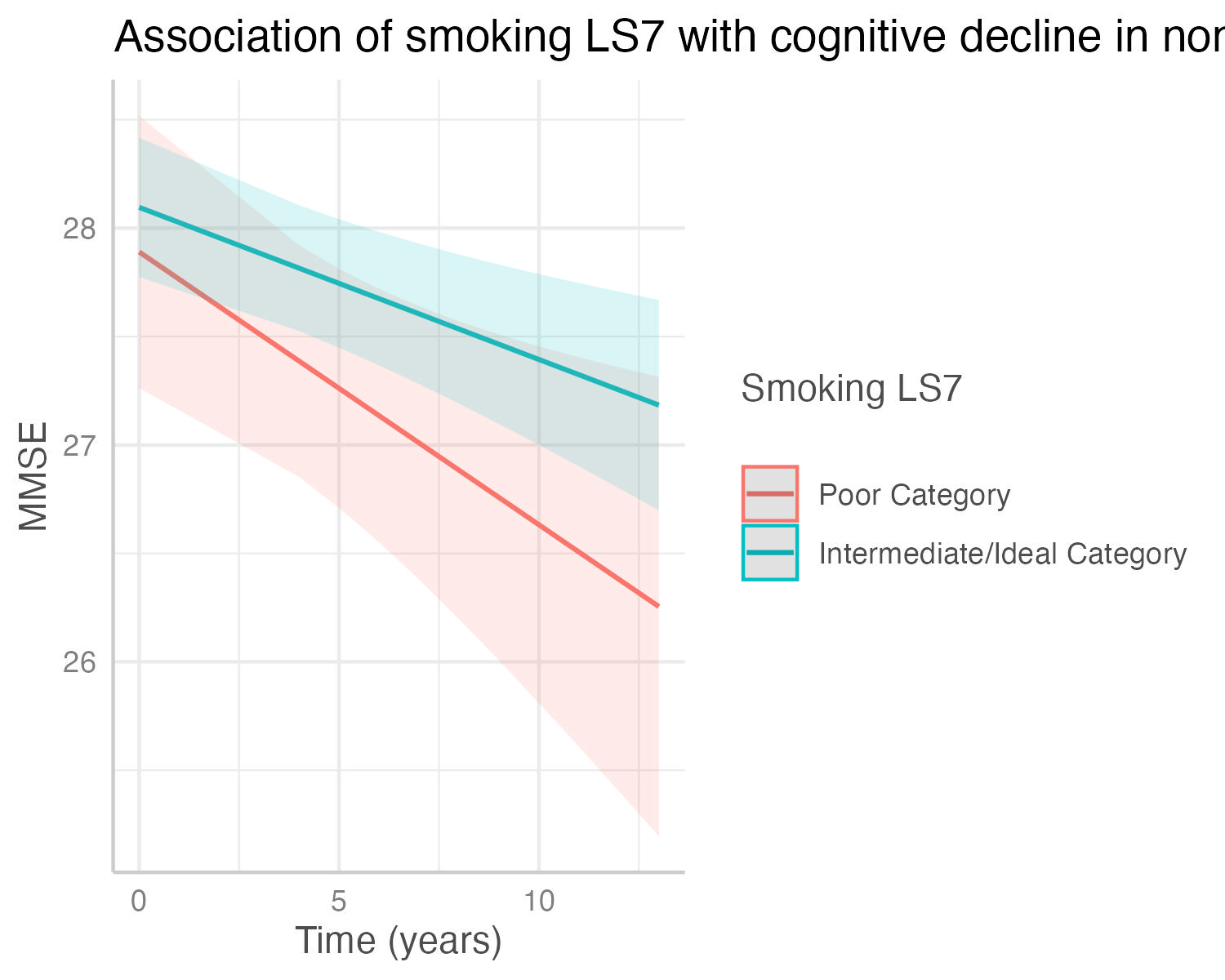

(a) Total Cohort	(b) Mexican American Cohort	(c) non-Hispanic White Cohort

### Supplementary Figure 16. Figure shows the decrease in cognitive state predicted by the interaction of LS7 diet categories with cognitive decline in the (a) total cohort, (b) Mexican American cohort, and (c) non-Hispanic White Cohort. Predicted outcomes and confidence interval bands are shown, with red showing the poor LS7 category and green the intermediate and ideal LS7 category.

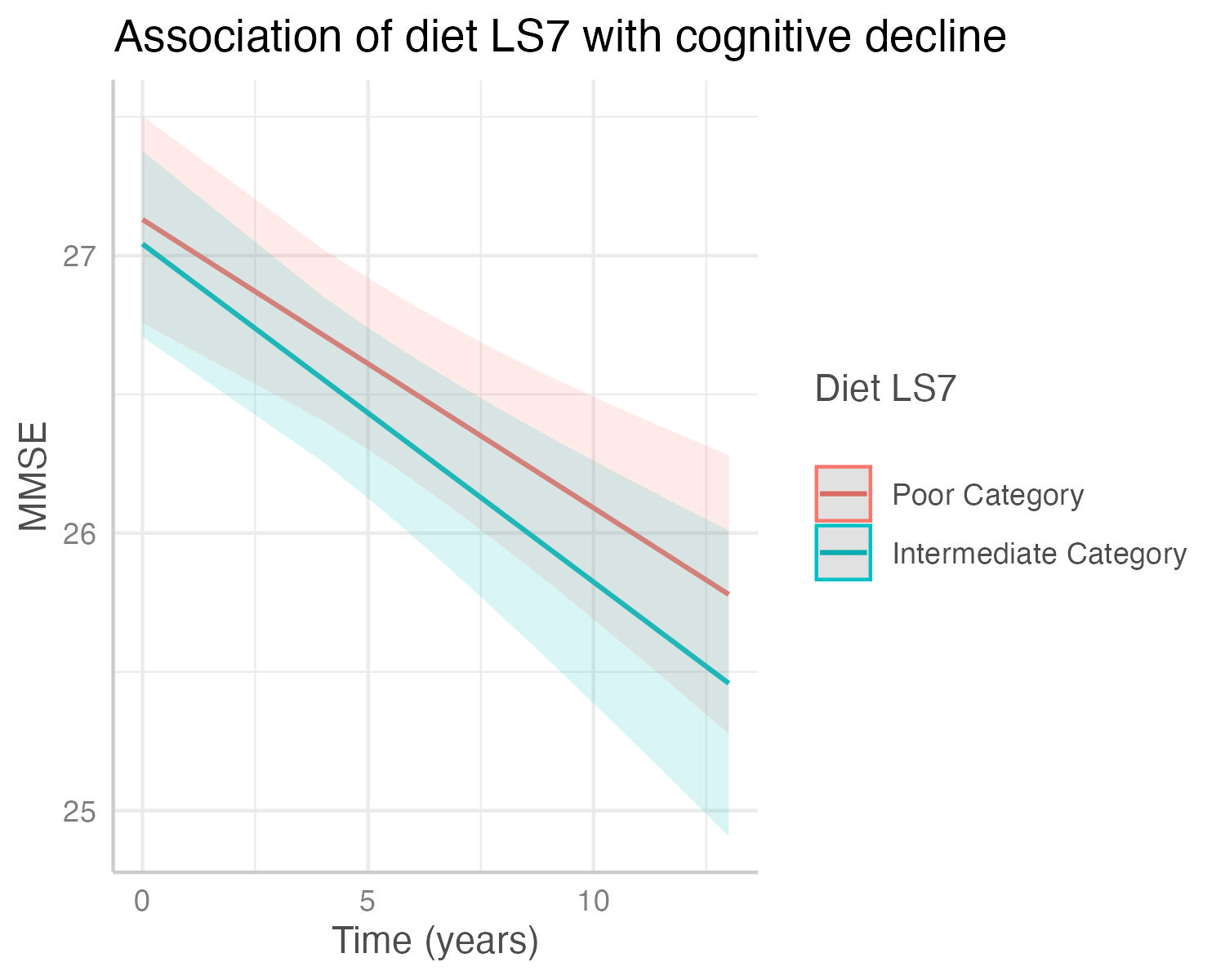

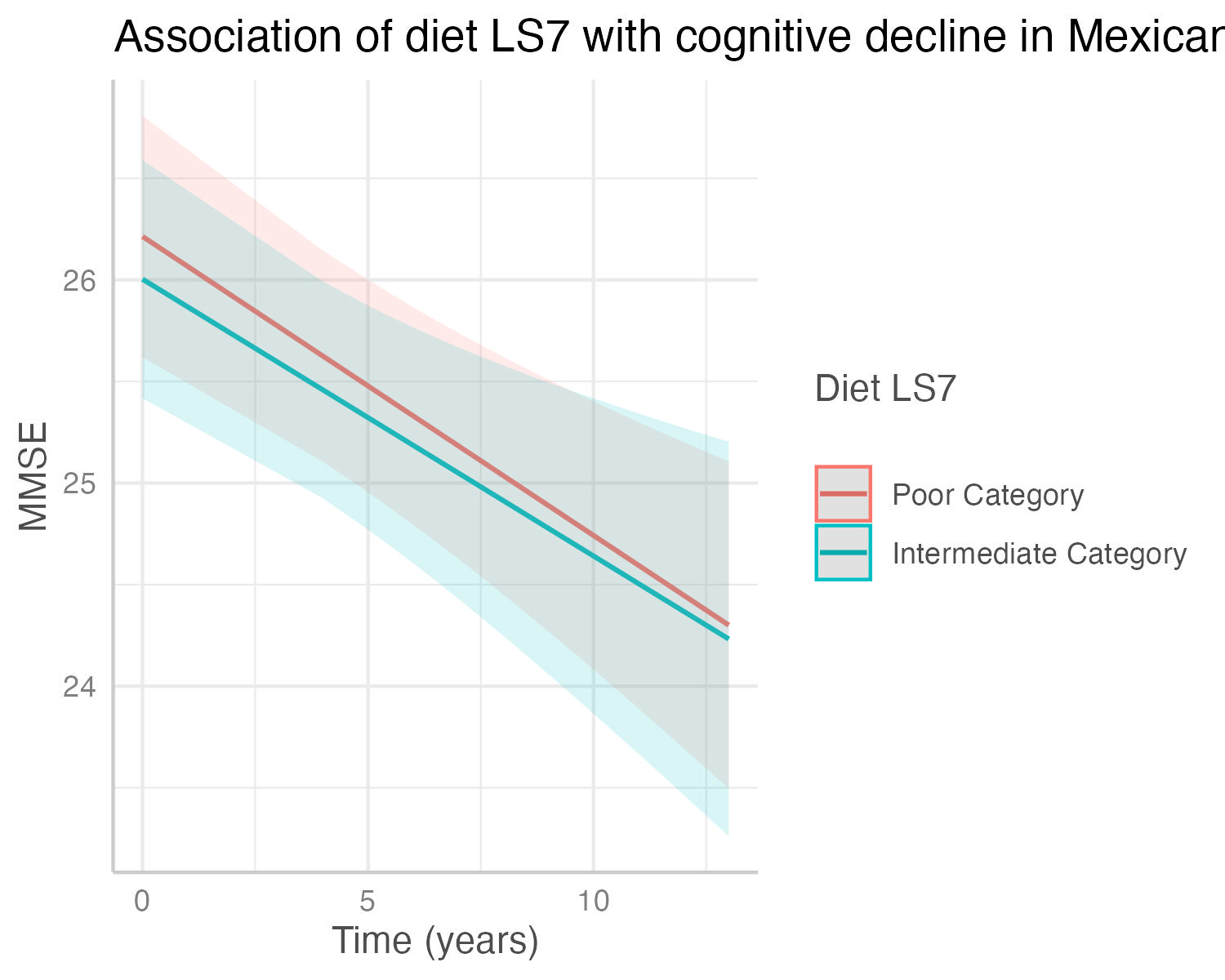

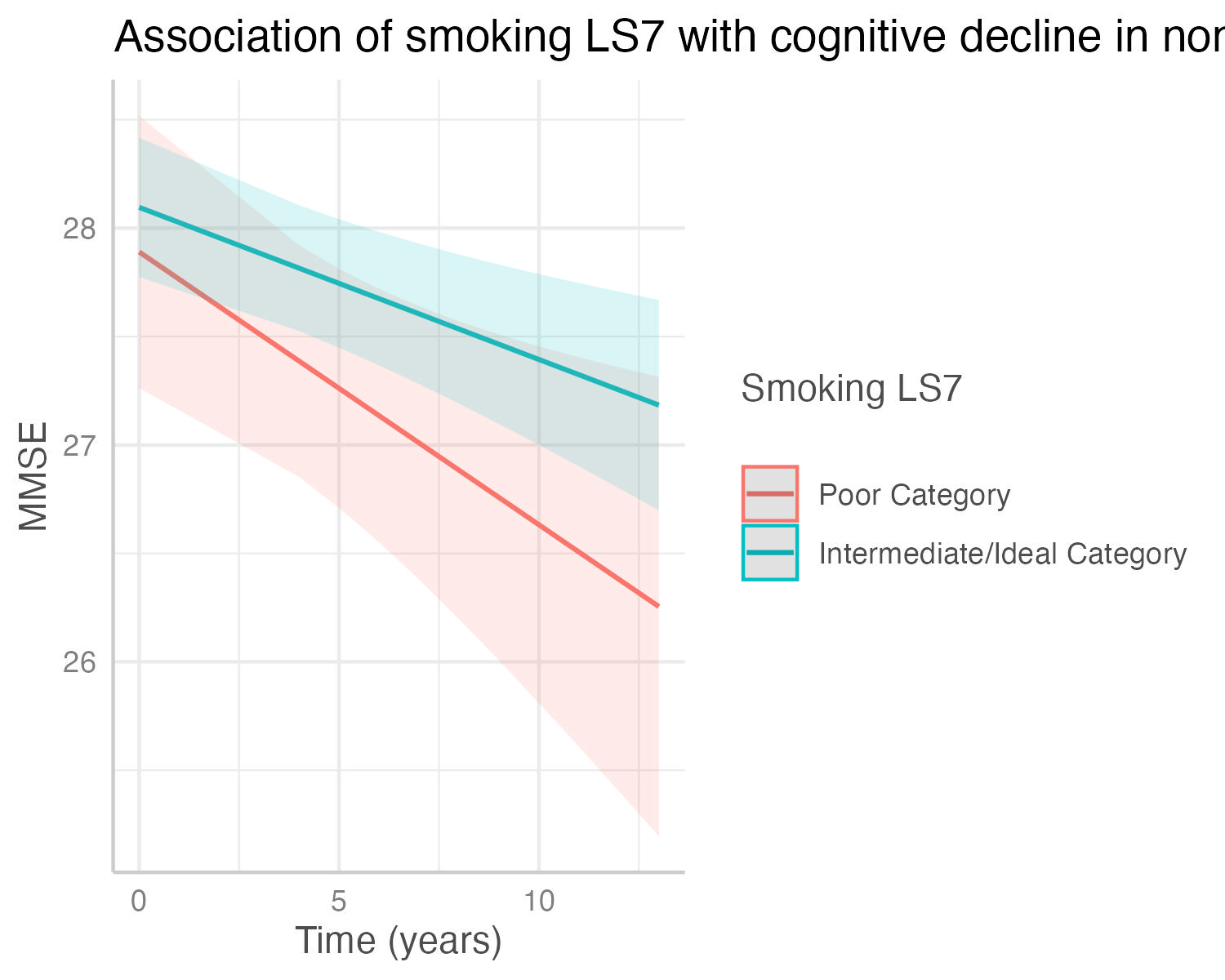

(a) Total Cohort	(b) Mexican American Cohort	(c) non-Hispanic White Cohort

### Supplementary Figure 17. Figure shows the decrease in cognitive state predicted by the interaction of BMI (kg/m2) with cognitive decline in the (a) total cohort, (b) Mexican American cohort, and (c) non-Hispanic White Cohort. Predicted outcomes and confidence interval bands are shown, with red showing individuals with lower BMIs, blue showing intermediate BMIs, and green higher BMIs.

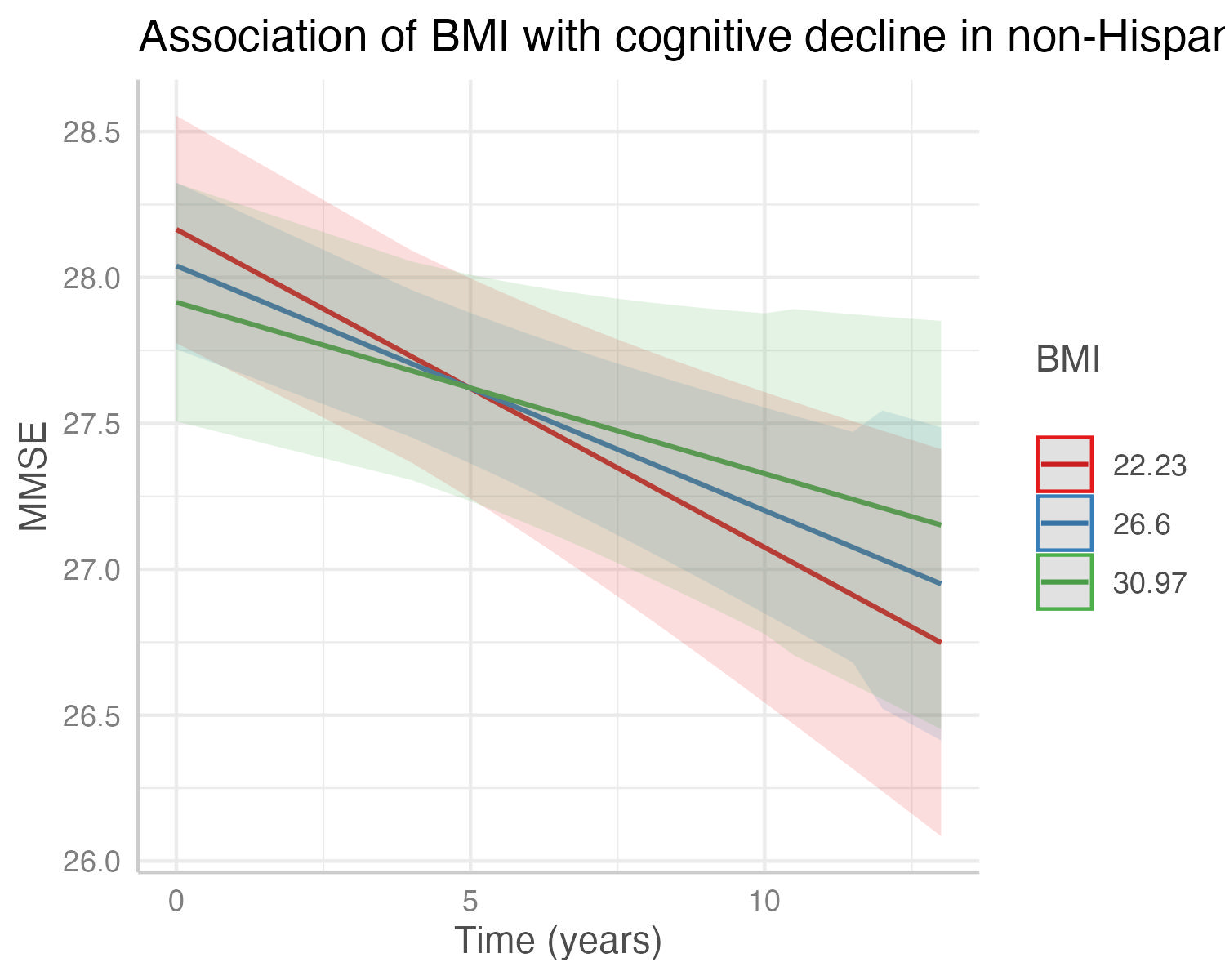

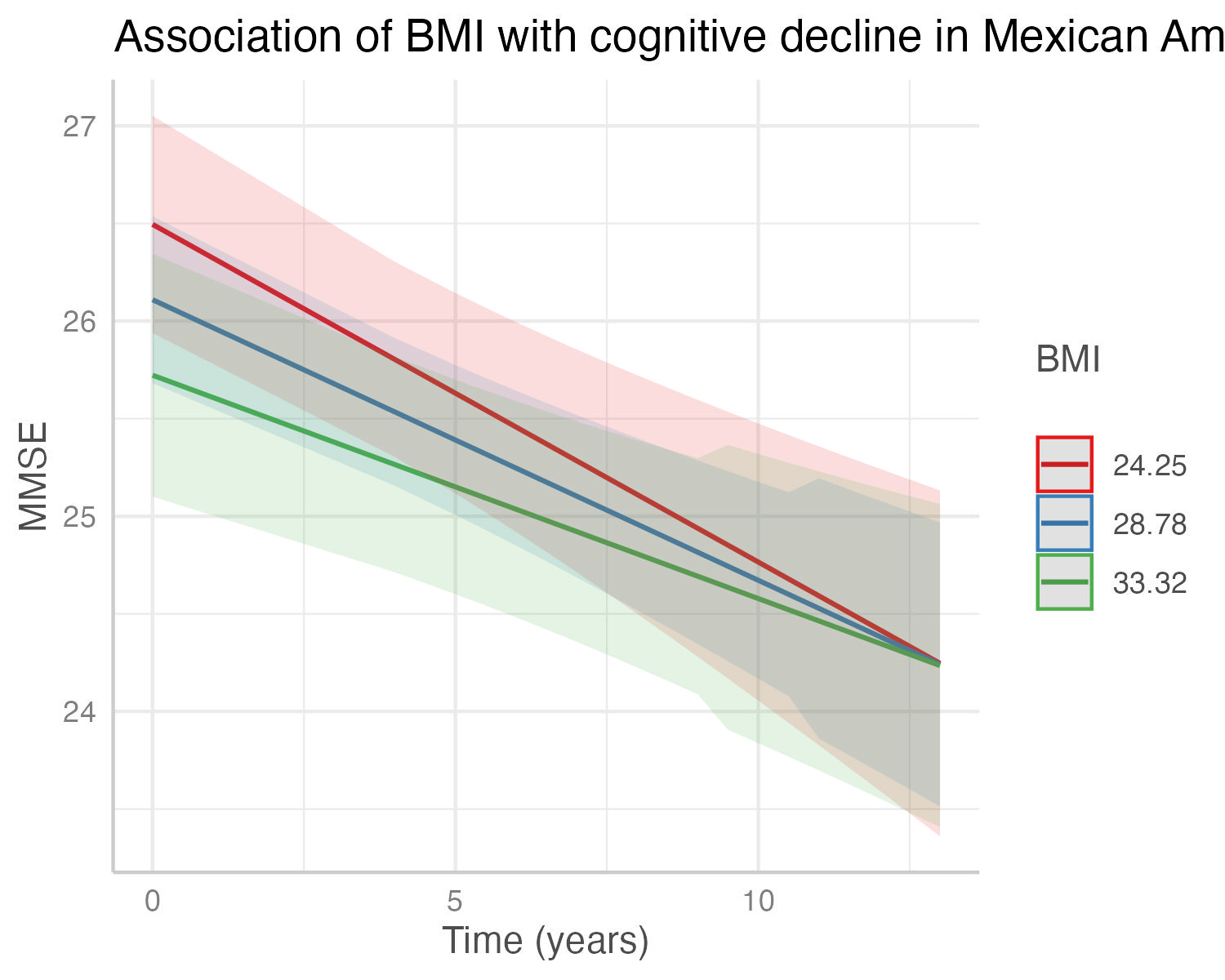

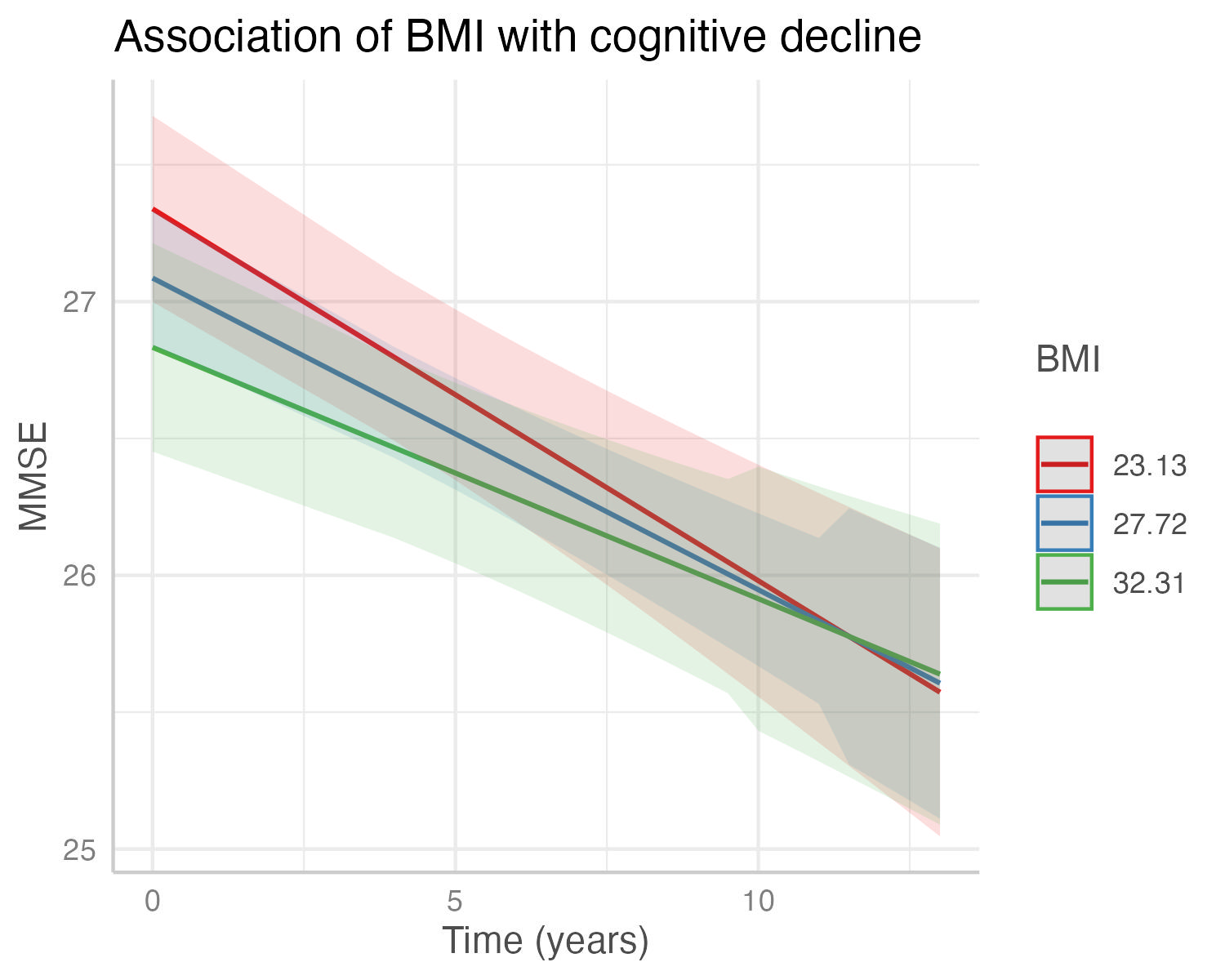

(a) Total Cohort	(b) Mexican American Cohort	(c) non-Hispanic White Cohort

### Supplementary Figure 18. Figure shows the decrease in cognitive state predicted by the interaction of systolic blood pressure (mmHg) with cognitive decline in the (a) total cohort, (b) Mexican American cohort, and (c) non-Hispanic White Cohort. Predicted outcomes and confidence interval bands are shown, with red showing individuals with lower systolic blood pressure, blue showing intermediate systolic blood pressure, and green higher systolic blood pressure.

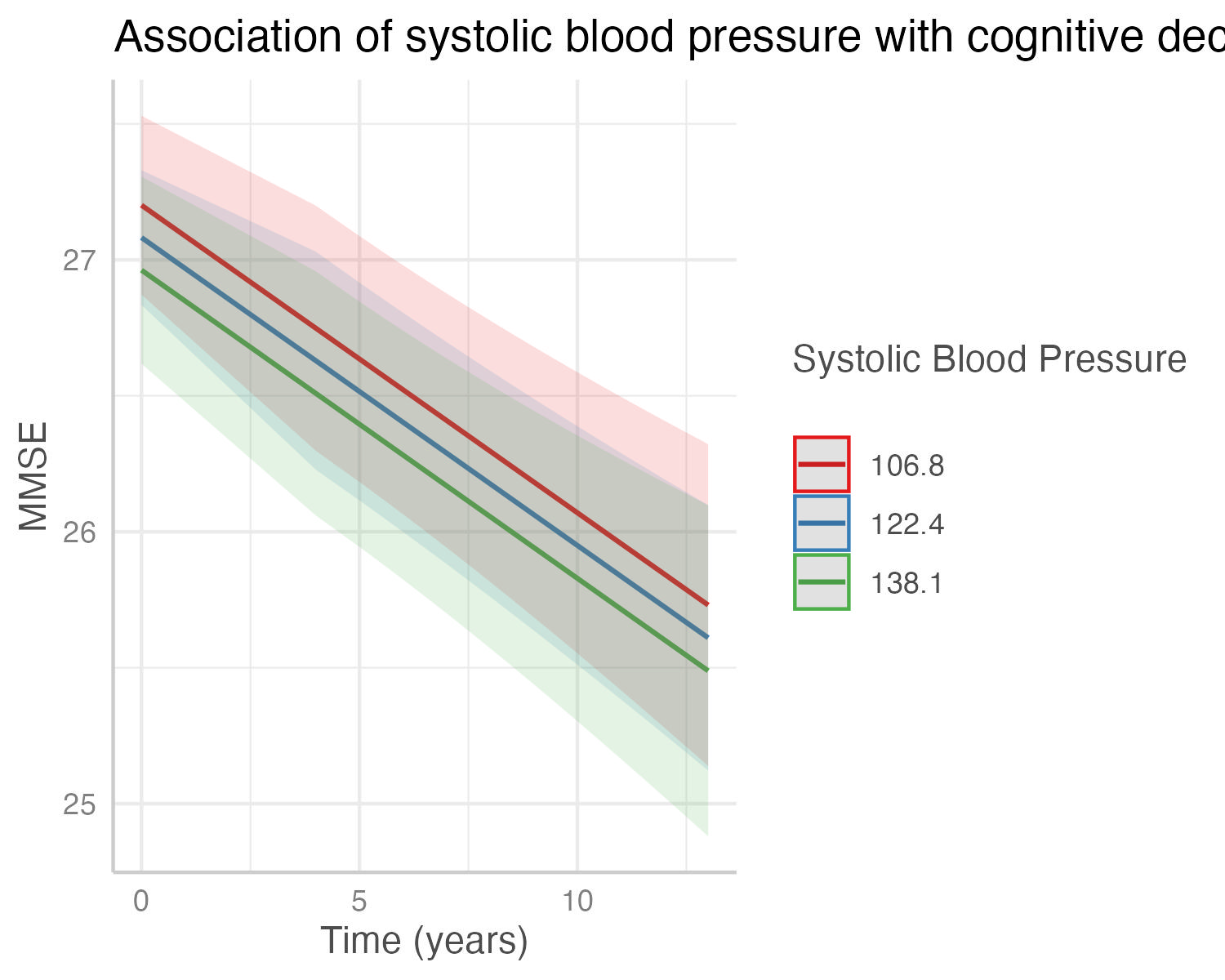

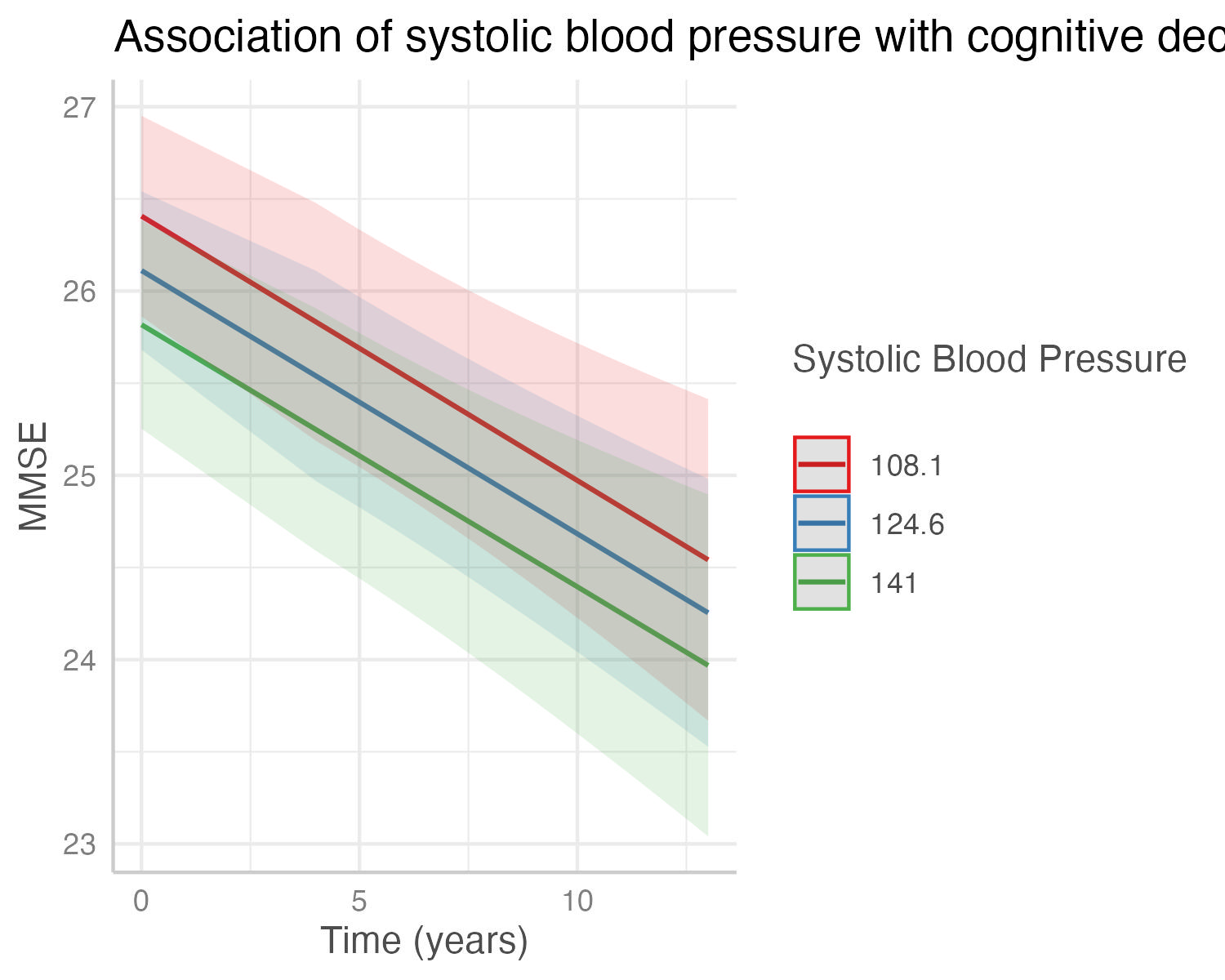

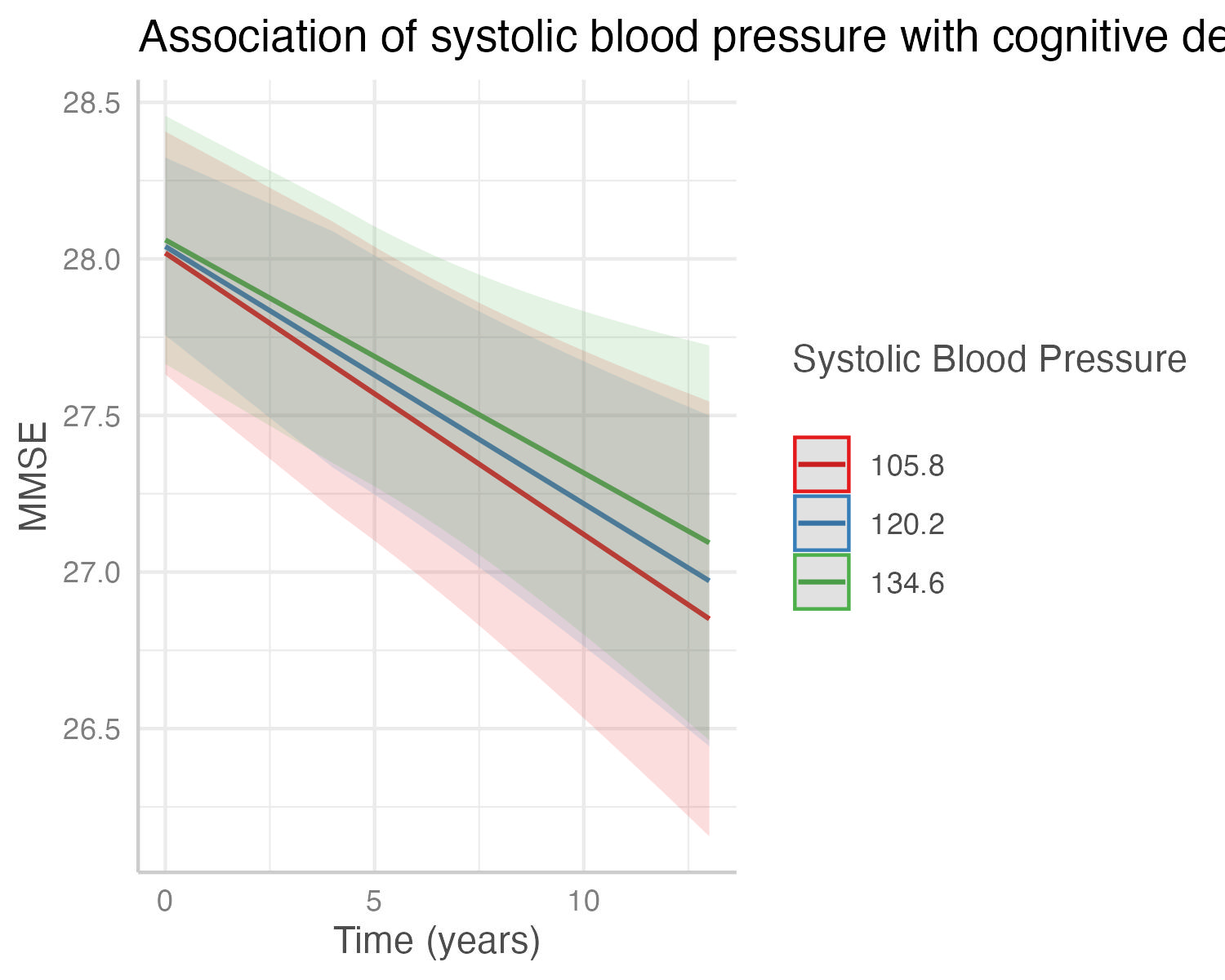

(a) Total Cohort	(b) Mexican American Cohort	(c) non-Hispanic White Cohort

### Supplementary Figure 19. Figure shows the decrease in cognitive state predicted by the interaction of diastolic blood pressure (mmHg) with cognitive decline in the (a) total cohort, (b) Mexican American cohort, and (c) non-Hispanic White Cohort. Predicted outcomes and confidence interval bands are shown, with red showing individuals with lower diastolic blood pressure, blue showing intermediate diastolic blood pressure, and green higher diastolic blood pressure.

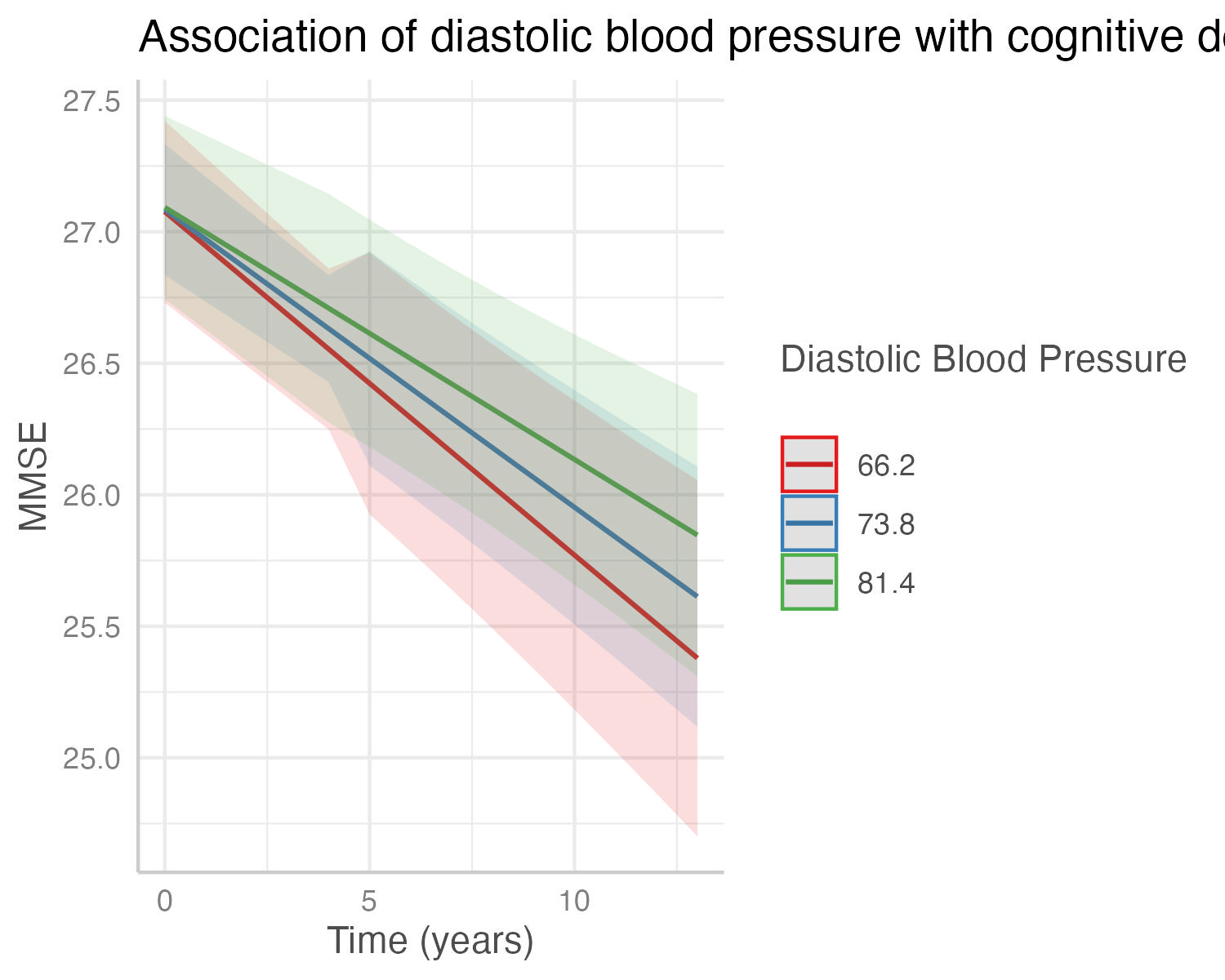

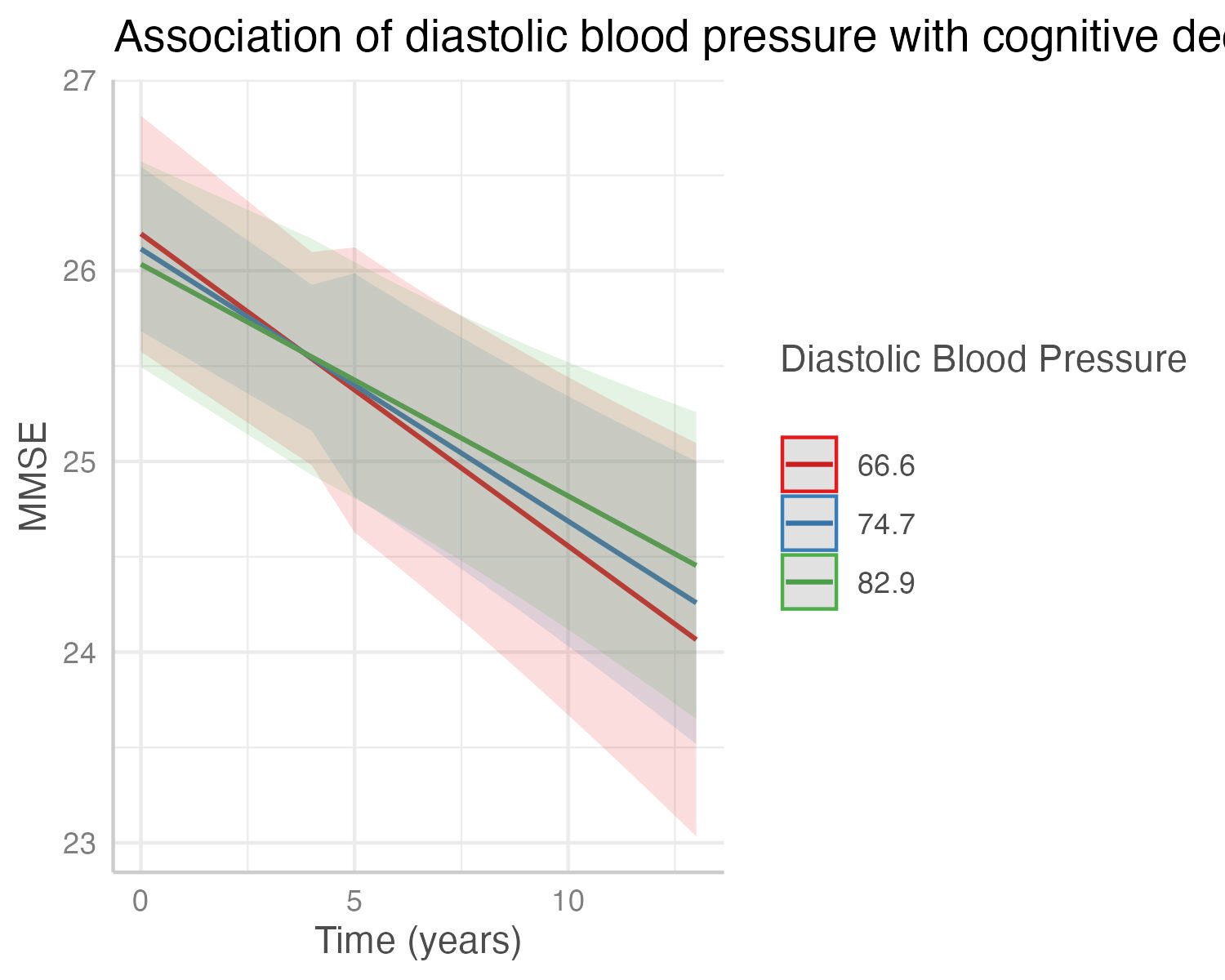

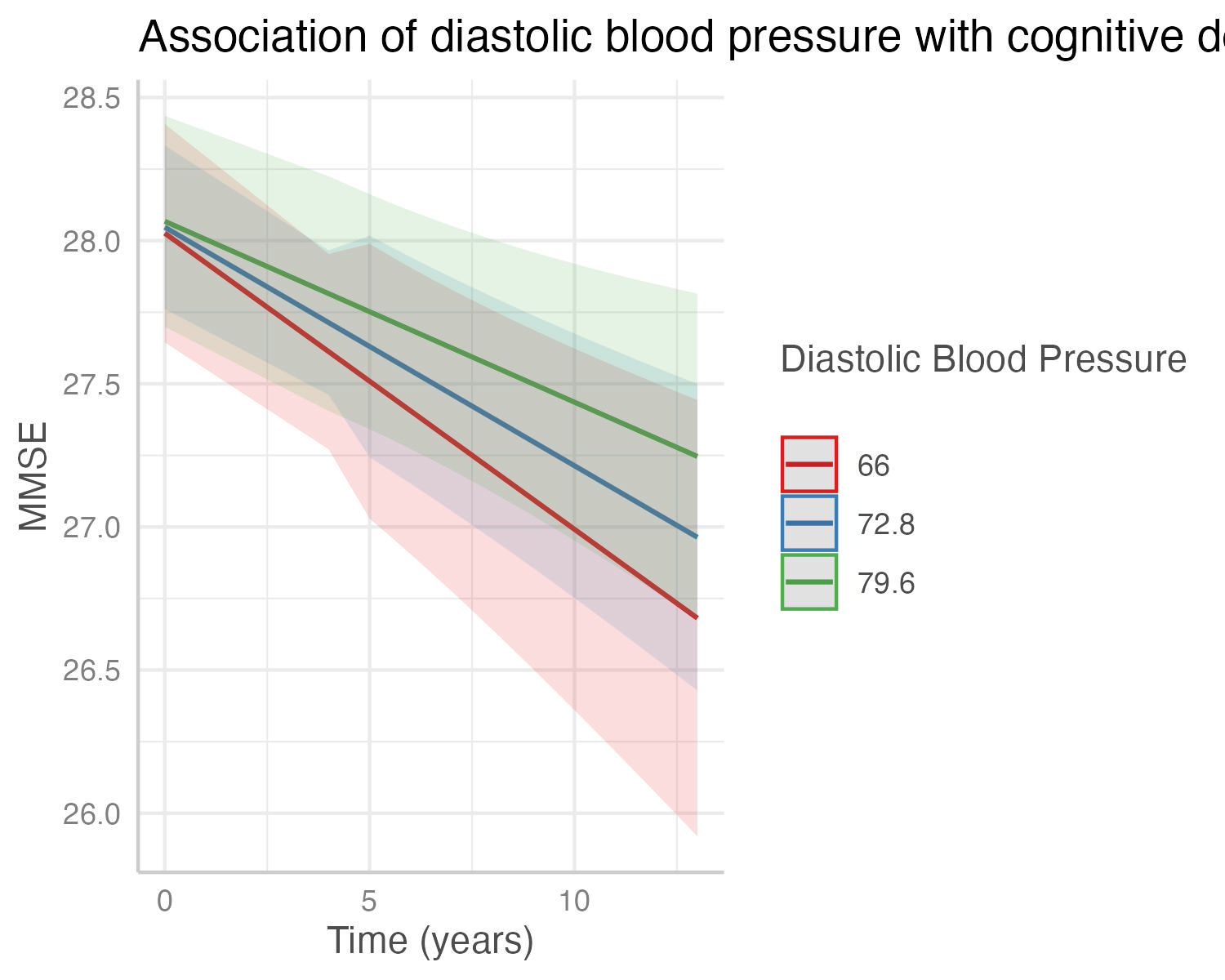

(a) Total Cohort	(b) Mexican American Cohort	(c) non-Hispanic White Cohort

### Supplementary Figure 20. Figure shows the decrease in cognitive state predicted by the interaction of fasting blood glucose (mg/dL) with cognitive decline in the (a) total cohort, (b) Mexican American cohort, and (c) non-Hispanic White Cohort. Predicted outcomes and confidence interval bands are shown, with red showing individuals with lower fasting blood glucose, blue showing intermediate fasting blood glucose, and green higher fasting blood glucose.

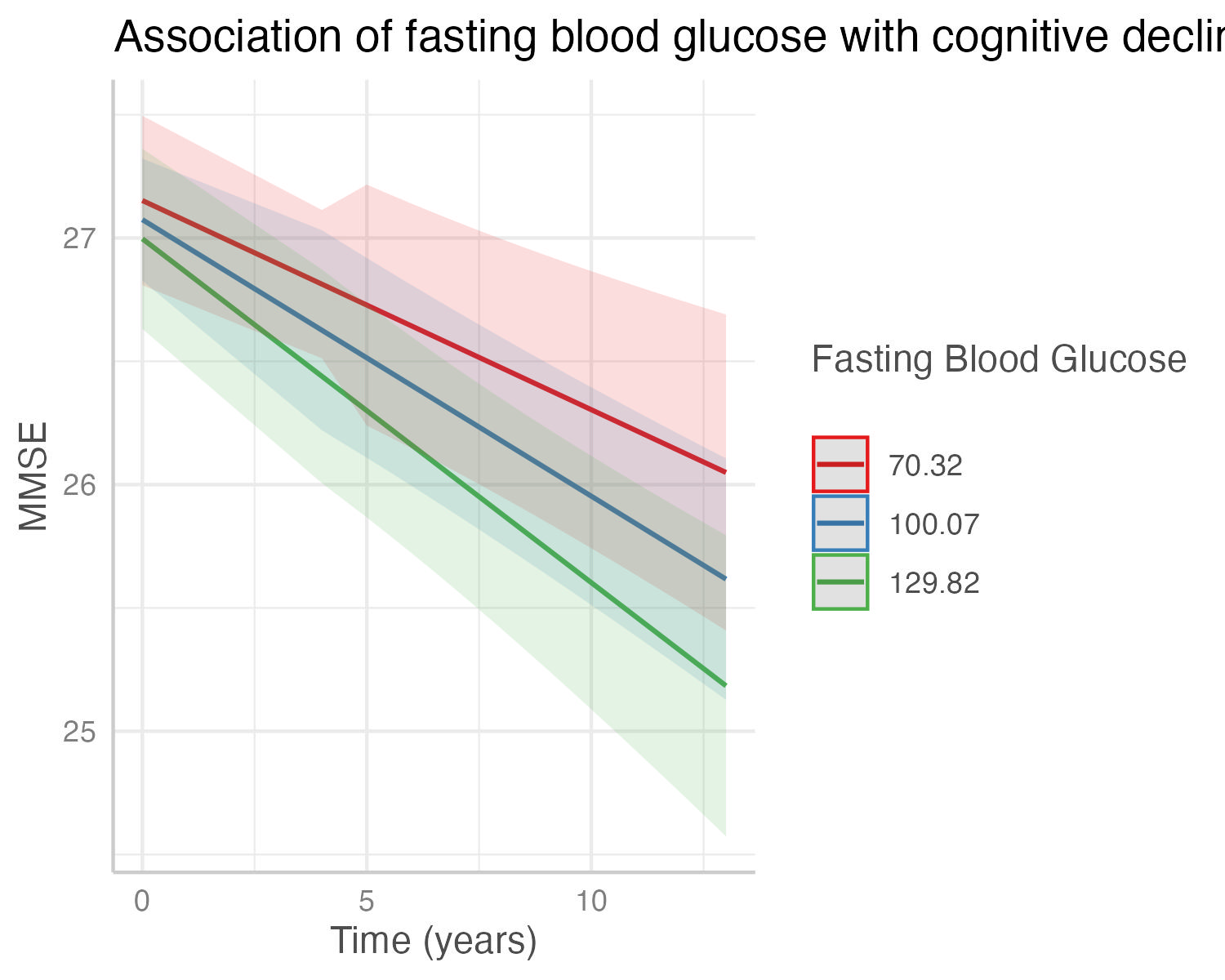

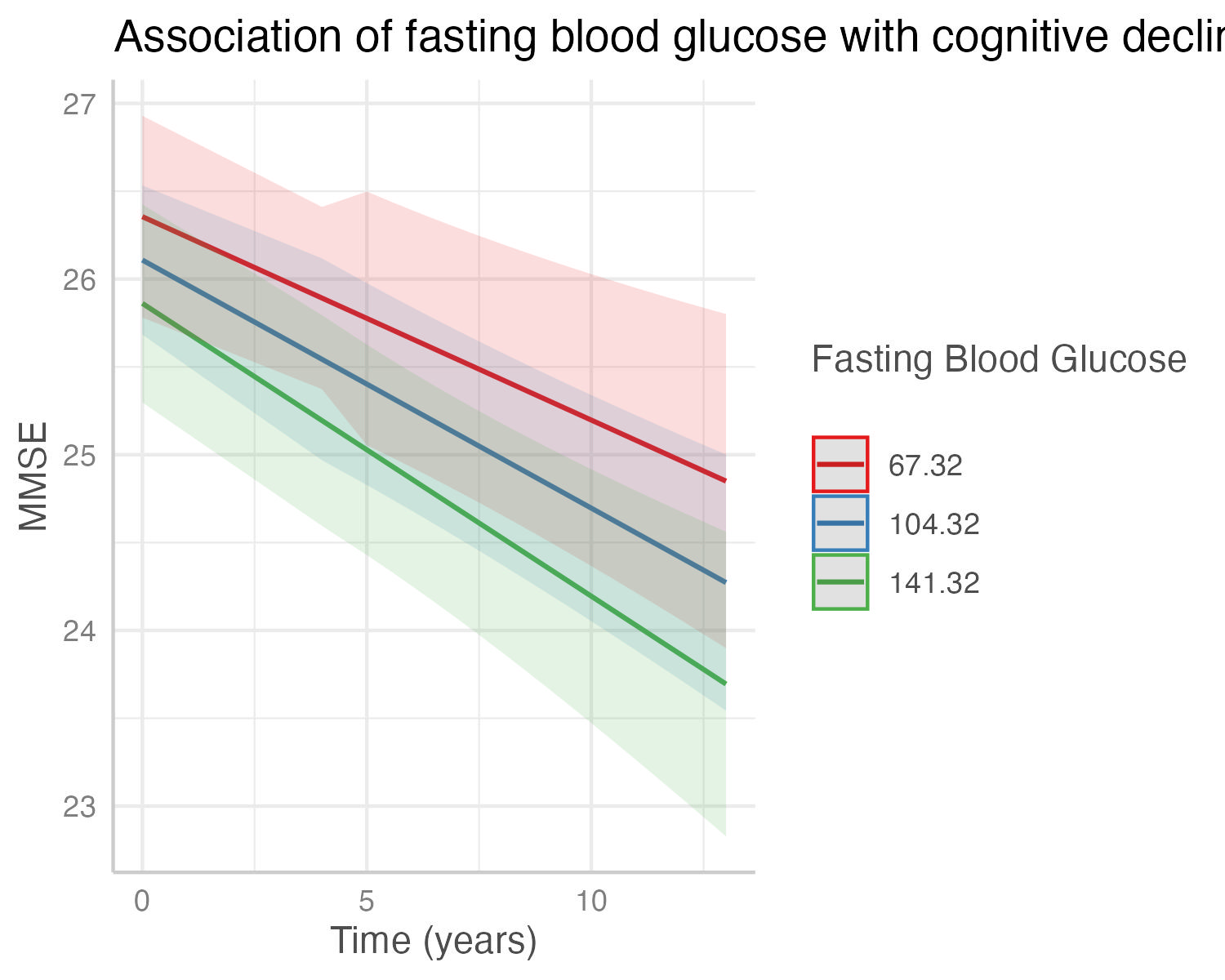

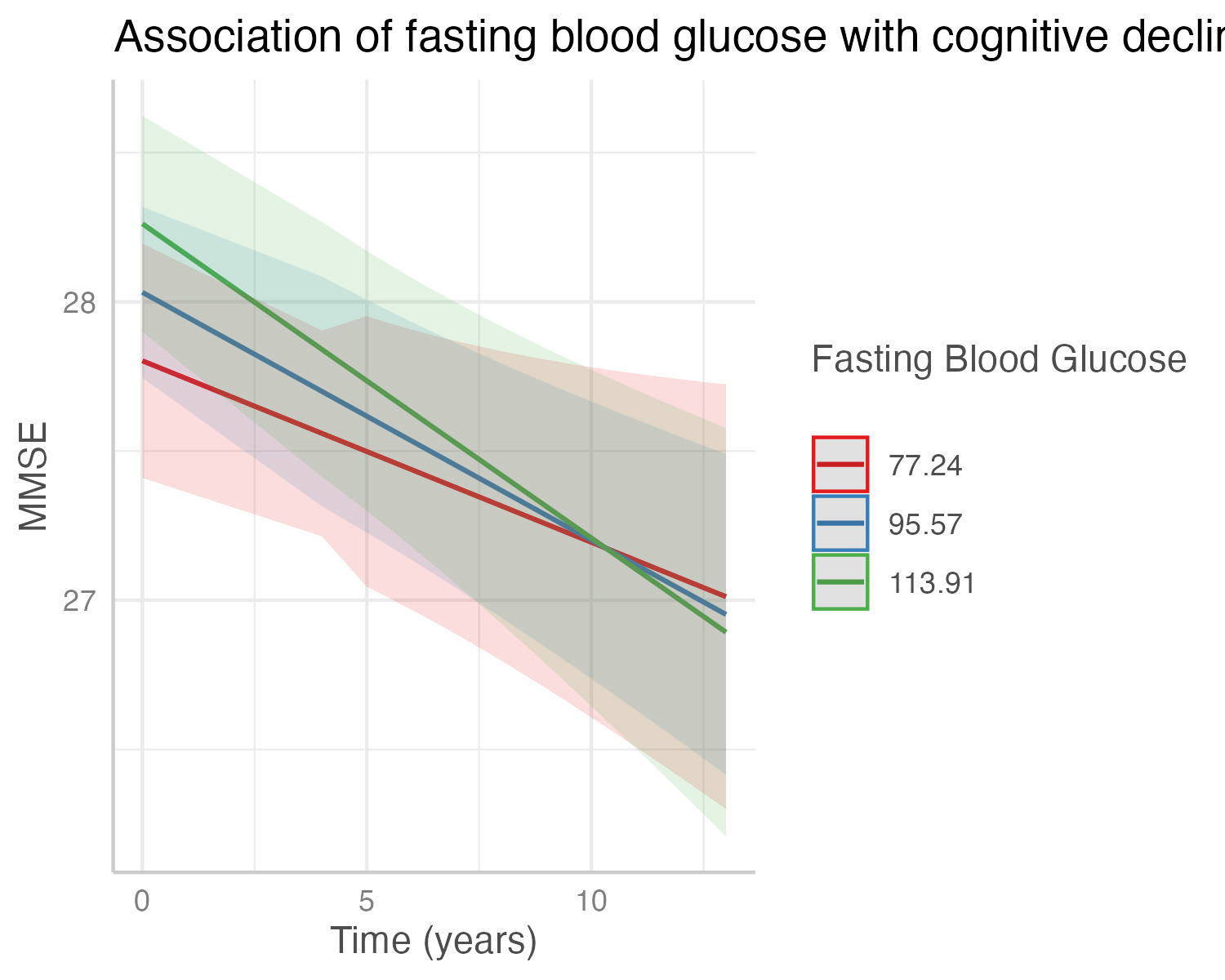

(a)Total Cohort	(b)Mexican American Cohort	(c)non-Hispanic White Cohort

### Supplementary Figure 21. Figure shows the decrease in cognitive state predicted by the interaction of exercise (minutes/week), the measurement used for physical activity LS7 score, with cognitive decline in the (a) total cohort, (b) Mexican American cohort, and (c) non-Hispanic White Cohort. Predicted outcomes and confidence interval bands are shown, with red showing individuals with lowest exercise minutes per week, blue showing intermediate exercise minutes per week, and green highest reported exercise minutes per week.

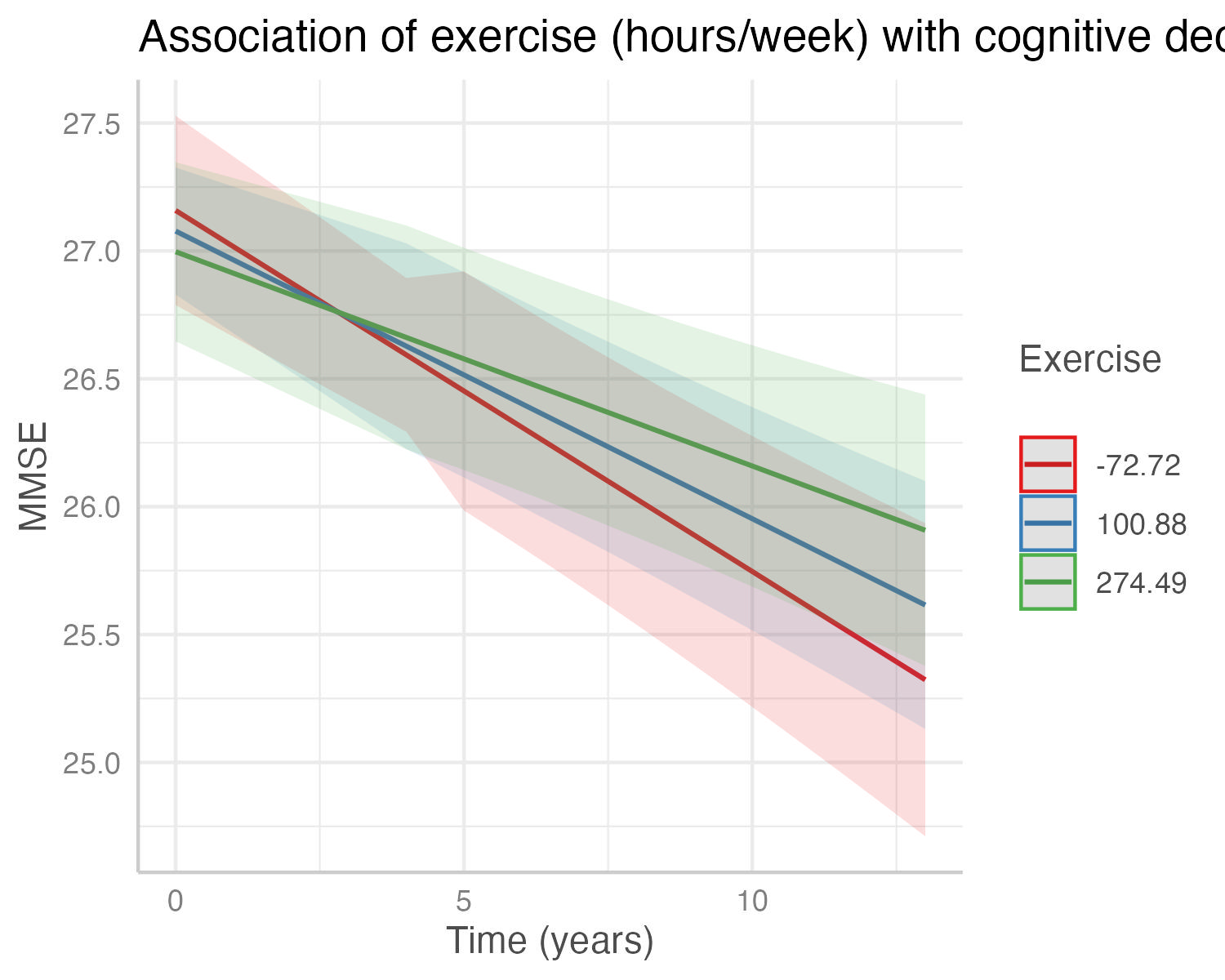

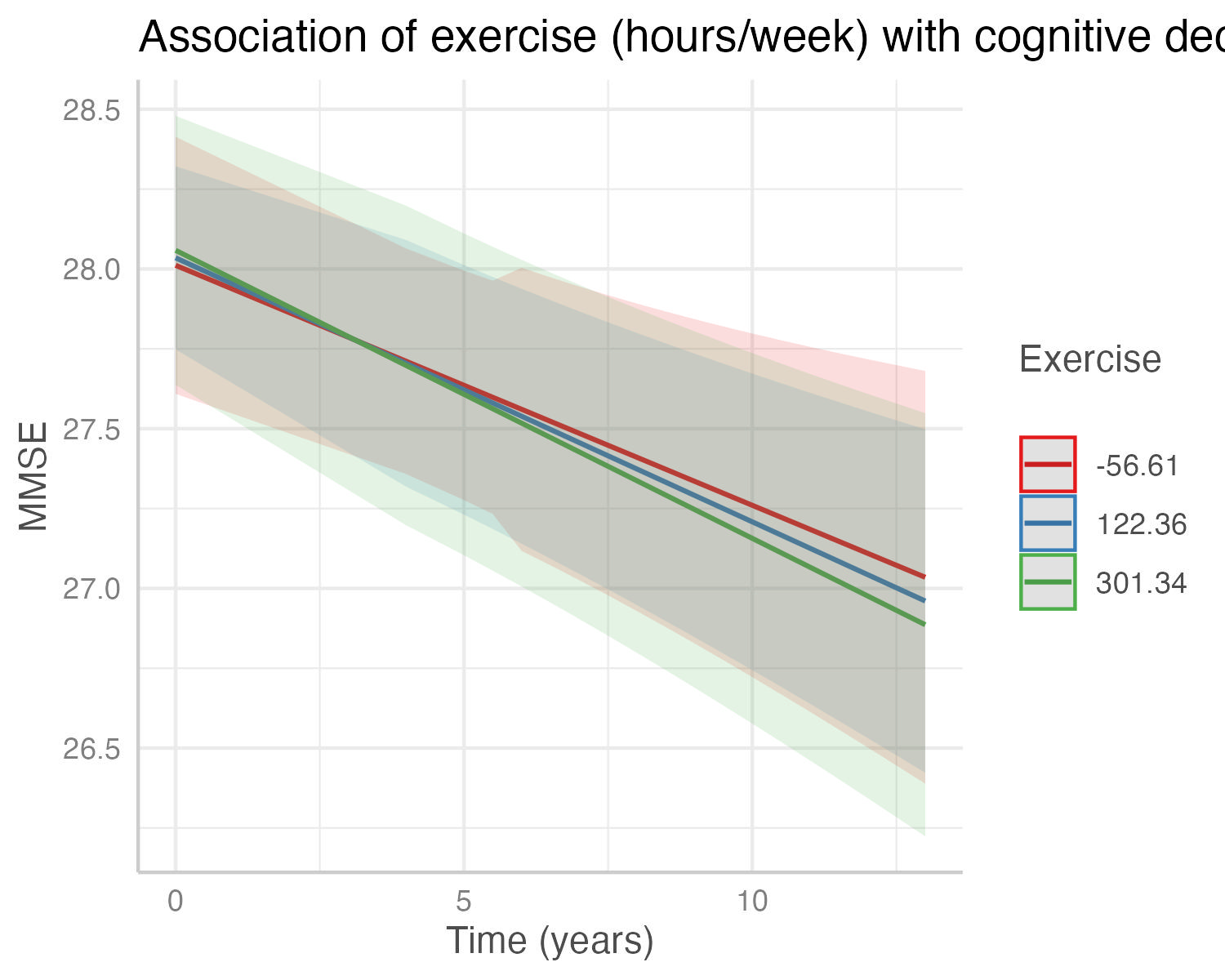

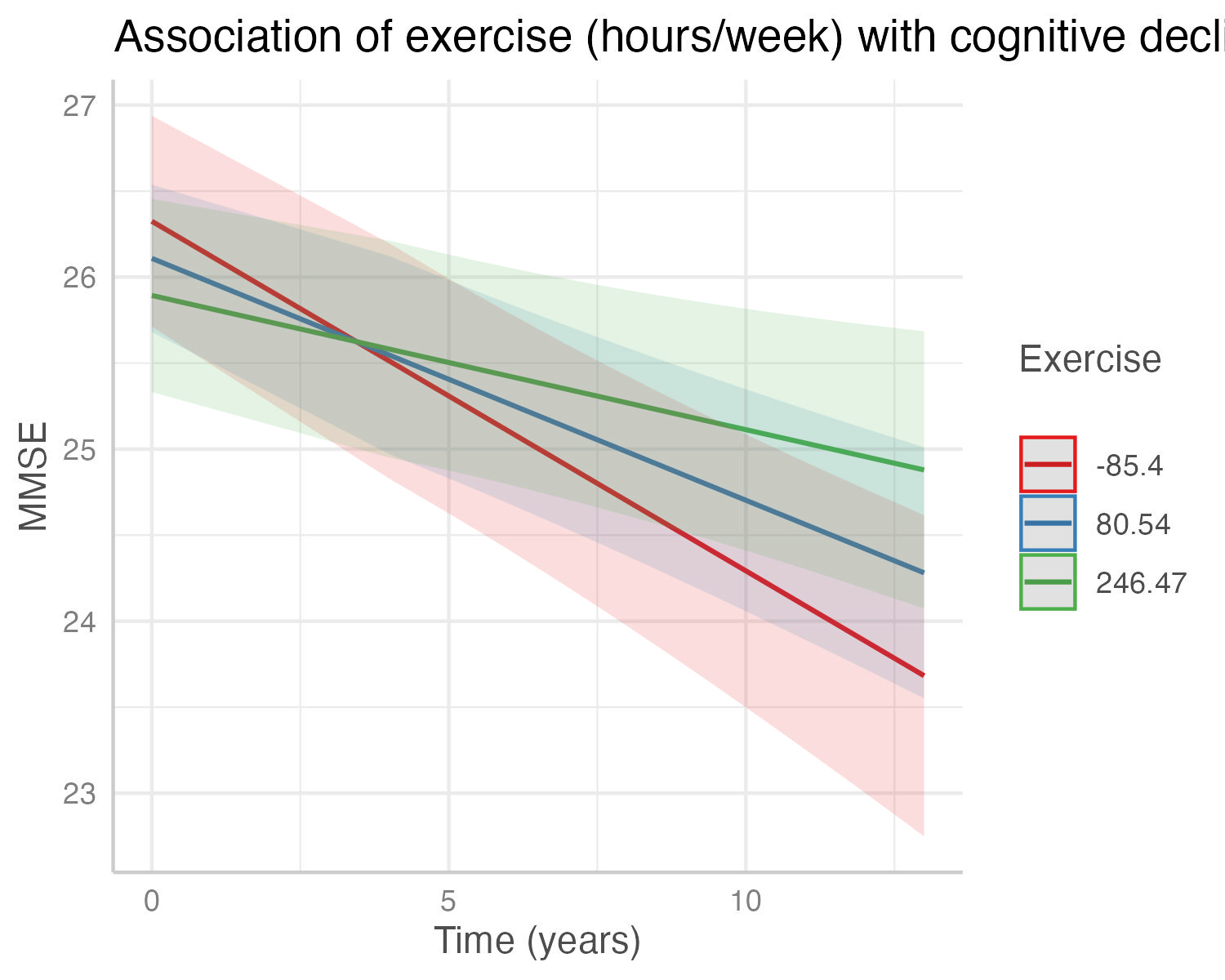

(a) Total Cohort	(b) Mexican American Cohort	(c) non-Hispanic White Cohort
